# Direct Pan-Cancer Multi-Omic Analysis of Radiotherapy Response Reveals Coding and Regulatory Genomic Variation, and Extracellular Adaptive Programs Underlying Radioresistance

**DOI:** 10.64898/2026.09.23.26363823

**Authors:** Preetha Ravi, Shree Chaitranjali Yadla, Venkatesh Kamaraj, Karthik Raman

## Abstract

Radioresistance remains a major barrier to effective radiotherapy, yet its molecular determinants remain incompletely understood. Most radiogenomic studies infer radioresistance using surrogate endpoints rather than from clinically annotated treatment response data. Here, we analysed genomic, transcriptomic, immune and clinical features of 1,445 radiotherapy-treated patients across 15 cancer types represented in The Cancer Genome Atlas with documented radiotherapy outcomes. Cancer-type-specific somatic association analyses identified 132 significant candidate variant associations across six cancer types, including three previously unannotated coding variants in TCF20, PTPN14, and CADM1. Functional annotation showed that radioresistant tumours were enriched for cytoplasmic, protein-associated, and metabolic functions, whereas radiosensitive tumours were enriched for nuclear regulatory programs. Through transcriptomic analysis, we identified 107 differentially expressed genes across 8 cancer types, enriched for extracellular signalling, secretory programs, and tumour–microenvironment interactions. The genomic and transcriptomic analyses revealed a limited gene overlap but converged on cellular adaptation and survival. Together, our findings suggest that radioresistance is characterised by complementary adaptive processes, whereby intracellular protein-regulatory and metabolic processes are coupled with extracellular signalling and tumour–microenvironment interactions. Clinically, radioresistance was associated with significantly poorer overall survival, whereas the radiosensitivity index and inferred tumour immune composition showed limited concordance with observed treatment response. To our knowledge, this study provides the first pan-cancer characterisation of clinically defined radiotherapy response and identifies potential candidate biomarkers for functional validation.

**Graphical abstract:** 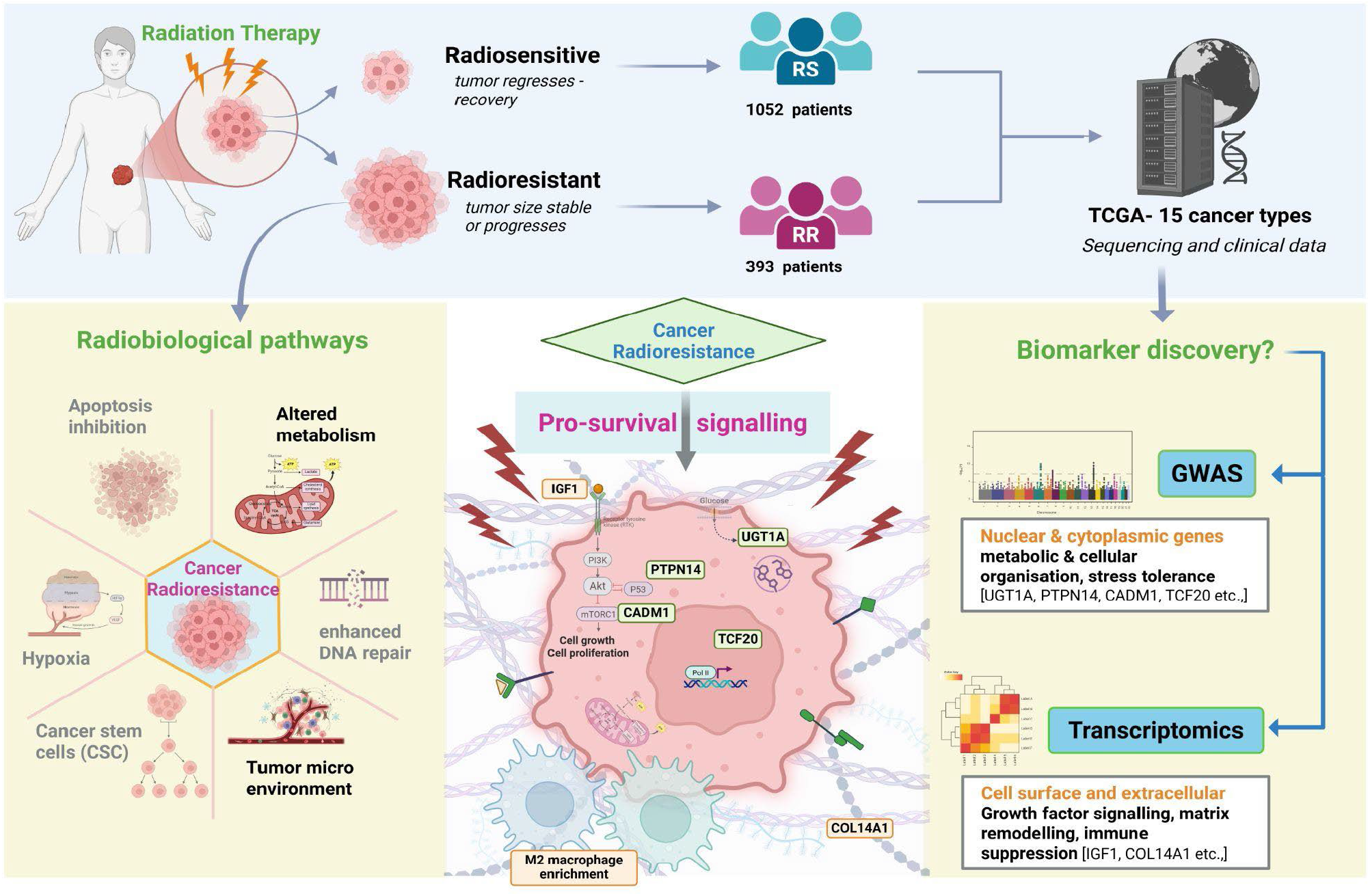

*Created in BioRender. Yadla, S. C. (2026)* https://BioRender.com/6jm1crm

## Introduction

Uncontrolled cell proliferation, a hallmark of cancer, is addressed through various therapeutic modalities such as surgery, chemotherapy, radiation therapy (RT), hormone therapy, and immunotherapy. Among these, RT remains a critical mode of treatment for many solid tumours (1). The administration of ionising radiation causes damage, either directly through double-strand breaks in DNA or indirectly through the generation of reactive oxygen species (ROS) by ionising the cell’s water content, ultimately leading to cancer cell death (2). The DNA damage response (DDR) involves a coordinated network of DNA damage sensors, repair proteins, and pro-survival signalling pathways leading to transcriptional changes, epigenetic alterations, and the activation of key transcription factors such as p53 and NF-κB (3). When the extent of damage exceeds the cell’s repair capacity, the DDR triggers apoptosis, senescence, or mitotic catastrophe, thereby eliminating damaged cancer cells and preventing their proliferation (4) However, resistance to treatment poses a significant challenge to therapeutic success and contributes to cancer recurrence and mortality (5). Radioresistance (RR) refers to the ability of tumour cells to survive and proliferate despite exposure to ionising radiation as a result of enhanced DNA damage response pathways (5).

RR arises from both cell-intrinsic and extrinsic factors. Intrinsically, cancer cells may enhance DNA repair mechanisms through homologous recombination (by using the sister chromatid as a template) and non-homologous end joining (6,7). Dysregulated cell cycle checkpoints in cancer cells permit continued proliferation despite DNA damage (CHK1/CHK2 for cell cycle arrest) (8,9). Tumour cells can suppress apoptosis by overexpressing anti-apoptotic proteins such as BCL-2, allowing them to evade cell death even after significant radiation-induced damage (8,9). Moreover, overexpression of antioxidant proteins, such as superoxide dismutase and glutathione peroxidase, neutralises the ROS generated by radiation, enhancing post-radiation survival (9,10). Overexpression of DNA damage sensors such as Ataxia Telangiectasia Mutated (ATM) and Ataxia Telangiectasia and Rad3-related (ATR) facilitates efficient detection and repair of radiation-induced lesions, thereby promoting tumour cell survival (11). Pro-survival signalling pathways, including PI3K–AKT, ERK/MAPK, and NF-κB, are frequently upregulated, enhancing proliferation, stress tolerance, and DNA repair capacity following irradiation (7). Hypoxia-adaptive responses mediated by HIF-1α and Hippo–YAP signalling regulate cellular proliferation, stemness, and epithelial–mesenchymal plasticity (7).

Externally, the tumour microenvironment (TME) becomes hypoxic due to the rapid growth of the tumour, resulting in increased demand for oxygen, which outpaces the supply from nearby blood vessels. The hypoxic and immunosuppressive conditions of the TME further support cancer stem cell maintenance, activate pro-survival pathways, and reduce ROS production, collectively contributing to RR (12,13). Growth factor–mediated signalling at the tumour–microenvironment interface further supports this phenotype, with pathways such as EGFR, HGF–MET, IGF1/IGF1R, and TGF-β contributing to survival and adaptive stress responses following irradiation (14,15). Metabolic reprogramming also plays a role, as RR cells may increase glycolytic flux while activation of the pentose phosphate pathway promotes NADPH production and antioxidant capacity (16). The presence of cancer stem cells (CSCs) further reinforces resistance, as these cells exhibit heightened DNA repair efficiency, ROS scavenging capacity, and quiescent cell-cycle states, enabling tumour repopulation after treatment (17). Stromal signalling and extracellular matrix remodelling can additionally contribute, as collagen deposition and matricellular proteins modify tissue stiffness and enhance integrin-mediated survival signalling, creating niches that protect tumour cells from radiation-induced damage (18,19).

At a molecular level, cellular responses to RT are mediated by genomic alterations, epigenetic modifications, altered gene expression, protein regulation, and signalling network rewiring. These processes operate across multiple biological scales and interact to shape radiotherapy response. Genomic alterations can generate abnormal transcripts and proteins that may contribute to RR. However, the extent to which these molecular changes influence treatment response remains incompletely understood. Consequently, RR must be examined across multiple layers of biological organisation to develop a comprehensive understanding of its underlying radiobiology.

While prior research has identified several pathways and genes linked to RR and radiosensitivity (RS), these findings are often derived predominantly through transcriptomic profiling or from small-scale studies and cell line models. Predictive tools such as the Radiosensitivity Index (RSI) and Genomic Adjusted Radiation Dose (GARD) have been developed from such data to stratify patients and personalise treatment (20,21). However, these approaches typically infer RR indirectly through surrogate markers like immune infiltration or survival outcomes, rather than using observed clinical response data. Despite advances in high-throughput sequencing, genome-wide investigations into the genetic basis of RR remain limited. Most insights are derived from differential gene expression rather than direct association studies at the DNA variant level. Leveraging large-scale, patient-derived datasets can enable broader, more systematic exploration of both common and cancer-type-specific biomarkers of RR.

In this study, we unravel the genomic and transcriptomic underpinnings of RR using multi-omics data from The Cancer Genome Atlas (TCGA) (22), accessed via the Genomic Data Commons (GDC) (23). The GDC hosts multidimensional datasets from the TCGA initiative, which catalogues genetic mutations responsible for cancer through large-scale genomic analyses across more than 11,000 patient samples spanning 33 different cancer types.

We examine tumours from patients with explicit clinical annotations of RT response, enabling direct comparison of RR and RS phenotypes. While one arm of the study focuses on using the somatic variant data available through TCGA and utilises genome-wide association study methods to identify single-nucleotide polymorphism (SNP)-level associations with RR, the other investigates expression-level differences associated with RR and provides a foundation for future functional validation and translational research in precision radiotherapy. This study adopts an exploratory, hypothesis-generating framework to identify preliminary candidate variants and transcriptomic patterns that may underlie differential RT response. Overall, by identifying genomic and transcriptomic signatures specific to RR and RS, we also provide a foundation for future functional validation and therapeutic translation.

## Methods

### Datasets

The Cancer Genome Atlas (TCGA) cancer cohort includes detailed clinical data on RT, encompassing treatment dose, radiation site, duration, and patient response. However, comprehensive RT information was available for only approximately 13.1% of cases (*n* = 1,445). Patient selection, response classification, and cancer-type inclusion criteria are summarised in Figure 1. Because the availability of clinically annotated radiotherapy response data differed across cancer types, the numbers of RR and RS cases varied between tumour cohorts. For this study, patient samples were categorised based on clinical response to RT: those with ‘complete’ or ‘partial response’ were classified as radiosensitive (RS). In contrast, those with ‘radiographic progressive disease’ or ‘stable disease’ following RT were categorised as radioresistant (RR).

**Figure 1:**
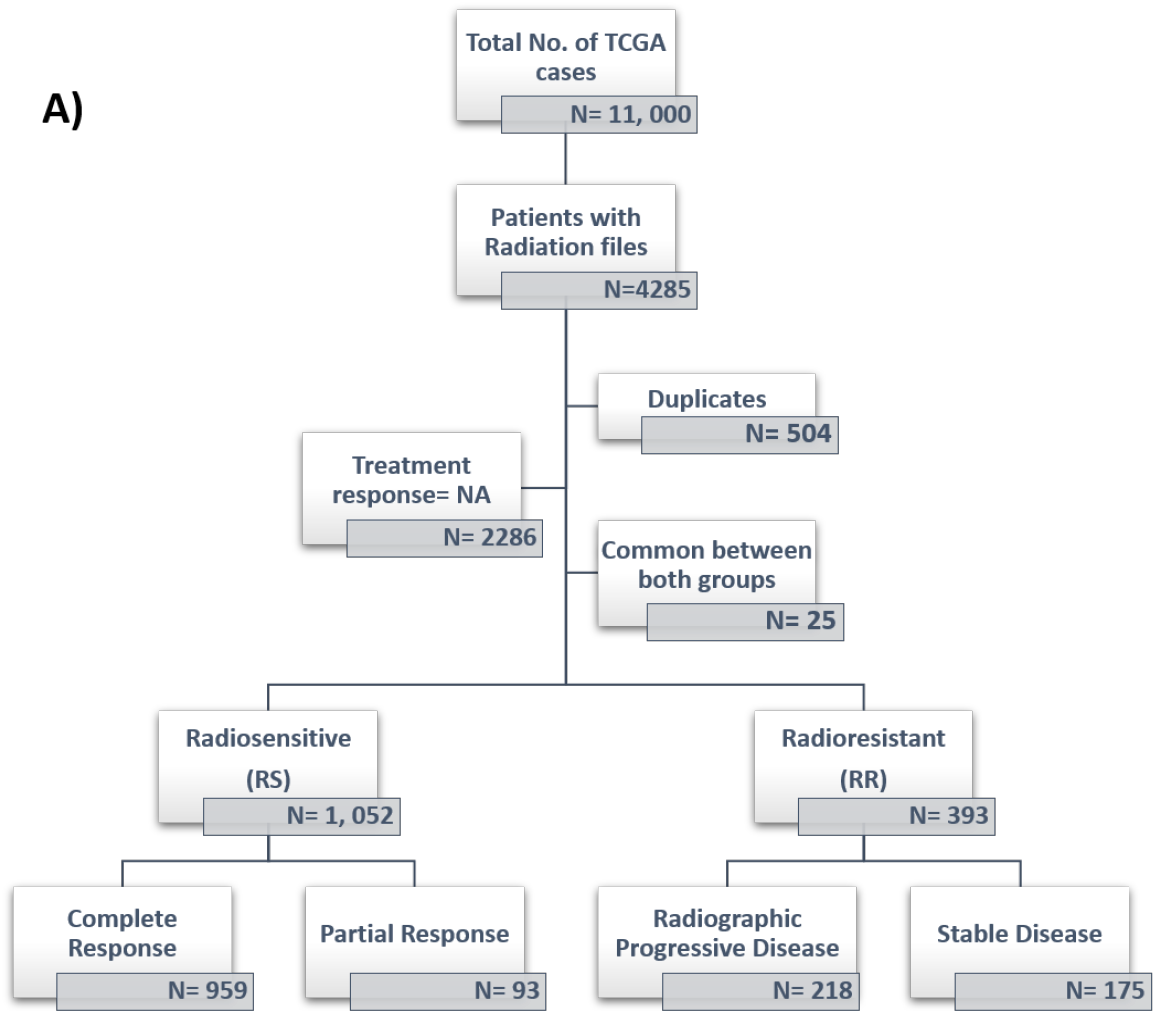
**TCGA cohort selection and RT response classification**. Flowchart displaying the number of cases available in the TCGA consortium, and the number of unique case IDs with available RT response data, categorised as radiosensitive (RS), RR, or [Not Available]. RS includes cases with a complete or partial response to RT, while RR includes cases with stable disease or radiographic progressive disease. The RR and RS patient barcodes are provided in Supplementary *Table 1*.

Cancer types were included in this analysis based on the following criteria: (a) availability of clinical annotations specifying patient response to RT, (b) a minimum of 10 samples each classified as RR and RS, and (c) RT being administered as the primary treatment modality. The case numbers in each cancer type are listed in Table 1.

**Table 1.** Dataset distribution across cancer types. Cancer-wise distribution of RR and RS cases of the top 15 cancer types, arranged in decreasing order of total case numbers are shown. The corresponding patient barcodes, UUIDs, and available RT response information, are provided in Supplementary Table 1.

| Primary site | Project name | Number of cases |  |  |
| --- | --- | --- | --- | --- |
|  |  | RS | RR | Total |
| Thyroid | THCA | 187 | 13 | 200 |
| Breast | BRCA | 175 | 12 | 187 |
| Larynx | HNSC | 154 | 17 | 171 |
| Brain | LGG | 43 | 124 | 167 |
| Cervix | CESC | 87 | 17 | 104 |
| Prostate | PRAD | 51 | 16 | 67 |
| Stomach | STAD | 41 | 15 | 56 |
| Uterus | UCEC | 49 | 7 | 56 |
| Esophagus | ESCA | 35 | 14 | 49 |
| Skin | SKCM | 24 | 23 | 47 |
| Bronchus & Lung | LUAD | 17 | 29 | 46 |
| Connective_Subcutaneous | SARC | 16 | 18 | 34 |
| Pancreas | PAAD | 14 | 14 | 28 |
| Bladder | BLCA | 16 | 12 | 28 |
| Bronchus & Lung | LUSC | 14 | 13 | 27 |

### Genome-wide Association Analysis of Somatic Variants

The overall somatic variant association analysis and downstream functional analysis workflow is summarized in Figure 2. Controlled-access somatic Variant Call Format (VCF) files were obtained from the NCI Genomic Data Commons (GDC) following approval for access through the NIH database of Genotypes and Phenotypes (dbGaP) under dbGaP accession: phs000178.v11.p8. Data were accessed and used in accordance with the applicable data-use terms.

**Figure 2:**
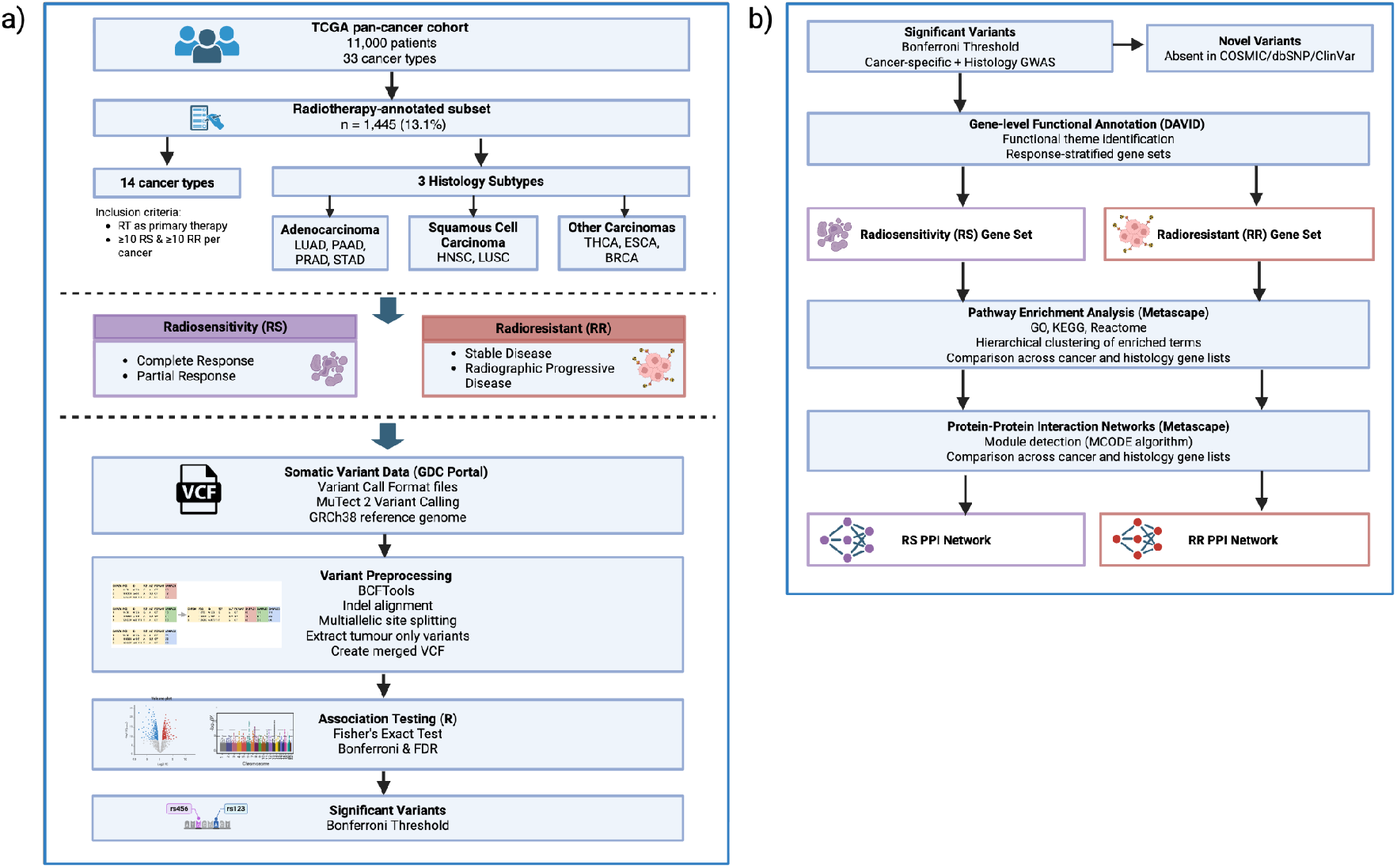
Overview of the somatic variant association and functional analysis workflow. (a) Somatic variant data (Mutect2, GRCh38) from TCGA were restricted to a radiotherapy-annotated subset (n = 1,445) and stratified into RS and RR groups across eligible cancer types. Variants were preprocessed (BCFtools) and tested for association using Fisher’s exact test with Bonferroni and FDR correction to identify significant loci. (b) Significant variants were functionally annotated (DAVID), stratified by association with response, and analysed for pathway enrichment (Metascape) and protein–protein interaction networks (MCODE). Variants absent from COSMIC, dbSNP, and ClinVar were classified as novel.

The VCF files were generated using somatic variant-calling pipelines, including VarScan2 (24), MuSE (25), and Pindel (26). Variant Call Format (VCF) files aligned to the human GRCh38 reference genome and generated using Mutect2 for SNPs and insertions/deletions (indels) were extracted for somatic variant association analysis. Extracted VCF files were preprocessed using custom Bash scripts incorporating BCFtools v1.19 (27) to prepare the data for downstream analysis in PLINK. Tumour-specific genotypes were extracted from paired tumour–normal VCFs, and variants with an allele count of zero were excluded. Individual tumour VCFs were merged within each cancer type and radiotherapy response group, and multiallelic variants were normalised into biallelic records using *bcftools norm -m-*. For each of 14 cancer types, merged VCF files were generated for RR and RS cohorts, containing all somatic variants relevant to that cancer and phenotype.

### Quality Control and Association Testing

Standard genome-wide association study (GWAS) quality-control (QC) procedures were applied to assess the behaviour of somatic variants under filters typically designed for germline data. QC steps included filtering for SNP missingness (--geno 0.2) and sample missingness (--mind 0.2) implemented in PLINK v1.9 (28,29).

Association testing was conducted in R v4.3.3 (30) using Fisher’s exact test (two-sided), selected due to the binary phenotype and limited number of clinically annotated samples. Multiple testing correction was applied using the Bonferroni method. The Benjamini–Hochberg false discovery rate (FDR) correction was additionally evaluated to explore more lenient thresholds. However, FDR-significant variants were considered exploratory and were used to identify a broader pool of candidate variants for hypothesis generation, whereas only Bonferroni-significant variants were retained for primary reporting and downstream functional analyses to prioritise a high-confidence set of candidate loci for biological interpretation and future validation.

### Visualisation

Somatic variant association results were visualised in R v4.3.3 (31) using the qqman package (32) to generate Manhattan plots and the ggplot2 package (33) to generate volcano plots and other custom visualisations. Volcano plots were constructed by plotting log₂-transformed odds ratios (log₂(OR)) against –log₁₀-transformed p-values. To enable visualisation on a finite log₂ scale, odds ratios of 0 and infinity, arising from phenotype-exclusive variant counts, were assigned values of 1/1024 and 1024, respectively, corresponding to log₂(OR) values of −10 and +10.

### Pooled Analysis by Histological Subtypes

To investigate therapy resistance at the histological level, the somatic variant association analysis workflow (Figure 2) was repeated using pooled cohorts. Adenocarcinomas included lung adenocarcinoma (LUAD), pancreatic adenocarcinoma (PAAD), prostate adenocarcinoma (PRAD), and stomach adenocarcinoma (STAD). Squamous cell carcinomas included head and neck squamous cell carcinoma (HNSC) and lung squamous cell carcinoma (LUSC). A third pooled group comprised thyroid carcinoma (THCA), oesophageal carcinoma (ESCA), and breast invasive carcinoma (BRCA). This grouping enabled the identification of shared variants across related cancer cohorts and increased statistical power through larger sample sizes. Significant variants from the pooled analyses were compared with cancer-specific results to identify areas of overlap and divergence.

### Functional Annotation

Significant variants (Bonferroni threshold) were functionally annotated using the Ensembl Variant Effect Predictor (VEP, v115.0) against the GRCh38.p14 reference genome and GENCODE transcript set (34). Variant consequences were assigned according to Sequence Ontology terms (e.g., missense_variant, frameshift_variant, intron_variant). To reduce redundancy, variants annotated to multiple transcripts were collapsed into a single representative entry by prioritising Ensembl canonical annotations with interpretable gene symbols and protein-coding biotypes where available. MANE (Matched Annotation from NCBI and EMBL-EBI) Select annotations, representing transcripts jointly curated by Ensembl and RefSeq, were retained preferentially when multiple canonical transcript annotations were present.

### Variant Prioritisation and Subset Analysis

Following annotation, two complementary variant sets were generated for downstream analysis: all significant variants that passed the Bonferroni correction, and a subset of novel variants. Position- and allele-specific queries were performed against major public variant databases, including dbSNP and ClinVar, to assess novelty. Since novel variants are nested within the larger set of significant variants, subsequent enrichment analyses were performed on the complete set of significant variants to capture overall biological patterns. However, novel variants were functionally annotated individually for potential biological relevance.

Variants were classified according to their predicted functional impact using VEP. High- and moderate-impact variants (e.g., frameshift and in-frame deletions, splice donor and acceptor variants, missense variants, and in-frame insertions) primarily affect protein-coding sequences and were therefore classified as coding variants. In contrast, low- and modifier-impact variants (e.g., upstream, downstream, intronic, synonymous, and untranslated region (UTR) variants) predominantly occur outside protein-coding sequences or are predicted to have minimal effects on protein structure. As these variants are more likely to influence gene regulation than protein sequence, they were classified as non-coding variants throughout this study. Protein-coding genes were prioritised as primary candidates for downstream analyses. Regulatory biotypes, including long non-coding RNAs (lncRNAs), microRNAs (miRNAs), and immunoglobulin/T-cell receptor genes, were retained to capture potential regulatory contributions, whereas pseudogenes and to be experimentally confirmed (TEC) annotations were excluded because of limited functional evidence. Full variant-level annotations and predicted impact classifications are provided in Supplementary Table 2.

### Functional Annotation and Enrichment

Genes corresponding to significant variants were subjected to functional annotation using the Database for Annotation, Visualization and Integrated Discovery (DAVID 2021 Update; Knowledgebase v2025_1) (35,36) using Ensembl gene identifiers (ENSG). Separate enrichment analyses were performed for RR and RS gene sets, each aggregated across cancer types and histological subgroups, to identify response-associated functional themes and annotation clusters. Given the exploratory nature of the analysis, both statistically significant and nominally enriched categories were examined to capture potential biological trends for further validation.

A combined multi-gene list analysis, incorporating both cancer-specific and histology-specific gene sets, was performed in Metascape v3.5 (34) for each radiotherapy response phenotype to enable comparative visualisation of enrichment patterns across tumour contexts. Pathway enrichment analysis was conducted using Gene Ontology, KEGG, Reactome, and other curated pathway resources, with enrichment significance assessed using hypergeometric testing independently for each input gene list. Enriched terms detected across all gene lists were pooled and hierarchically clustered based on similarity in gene membership using the kappa statistic (kappa ≥ 0.3) to reduce redundancy. For each resulting functional cluster, the term with the lowest p-value across all lists was selected as the representative term and visualised in a clustered heatmap, with enrichment strength displayed as −log10(p-value) to facilitate comparison across gene lists.

Protein–protein interaction (PPI) network analysis was subsequently performed for gene lists that yielded sufficient interaction density. PPI networks were constructed using interaction databases such as STRING, BioGRID, InWeb_IM, and OmniPath integrated within Metascape, and densely connected subnetworks were identified using the Molecular Complex Detection (MCODE) algorithm with default parameters. PPI and MCODE analyses were performed for pooled RR and RS gene sets, as well as for individual cancer- or histology-specific gene lists where sufficient interaction structure was observed.

### Differential Gene Expression Analysis

Differential gene expression (DGE) analysis between RR and RS patients was performed separately for each of the 15 cancer types using bulk RNA-sequencing data from The Cancer Genome Atlas (TCGA) (22). Analyses were restricted to primary tumour samples for all cancer types except skin cutaneous melanoma (SKCM), where the majority of both RR and RS samples were metastatic. Raw gene-level count data (STAR-Counts) were normalised using the limma–voom framework, which models the mean–variance relationship of log-transformed counts to enable linear modeling of RNA-seq data (37). Lowly expressed genes were filtered prior to analysis using *filterByExpr* (38). Linear models were fitted with RR versus RS as the primary contrast, and empirical Bayes moderation was applied to stabilize variance estimates (37). Differential expression statistics were reported as log₂ fold changes with Benjamini–Hochberg false discovery rate (FDR) correction. Genes with an absolute log₂ fold change ≥ 1.5 and FDR ≤ 0.05 were considered significantly differentially expressed genes (DEGs). In addition, genes with nominal p-values (p < 0.1) were reported as exploratory candidates for future validation. Principal component analysis (PCA), MA plots, volcano plots, and per-gene expression visualizations were generated using voom-normalised expression values to assess sample separation and robustness of the results.

### PPI network analysis

PPI network analysis was performed for the differentially expressed genes (DEGs) using STRING (STRINGdb) (39). The network comprised 67 protein-coding genes and 11 non-coding RNAs and was constructed using the full STRING network, in which edges represent both functional and physical protein associations. Interactions were filtered using a medium confidence threshold (interaction score ≥ 0.3). Markov Cluster Algorithm (MCL) clustering was applied to identify natural network modules based on stochastic flow. Functional enrichment analysis of the network genes was performed for Gene Ontology (GO) cellular component and molecular function categories (40).

### Tumour Purity and Immune Cell Infiltration Assessment

Tumour purity and immune cell composition were assessed to characterize the TME associated with radiotherapy response. Tumour purity estimates were obtained from precomputed ABSOLUTE purity calls provided by the TCGA Pan-Cancer Atlas (41). Immune cell infiltration was quantified using CIBERSORT LM22 immune cell fraction estimates derived from TCGA RNA-seq data (42). Sample identifiers were harmonized by mapping aliquot-level barcodes to patient-level identifiers, restricting analyses to primary tumour samples (01A), and averaging immune fractions when multiple samples per patient were present. For each cancer type, immune cell fractions and purity values were compared between RR and RS groups using non-parametric Wilcoxon rank-sum tests. Median differences (RR − RS) were calculated for each immune cell type, and multiple testing correction across the 22 immune populations was performed using the Benjamini– Hochberg method. These analyses were used to identify immune and stromal features potentially associated with radiotherapy resistance or sensitivity.

### Radiosensitivity Index (RSI) Calculation

Tumour-intrinsic radiosensitivity was quantified using the Radiosensitivity Index (RSI), calculated according to the original 10-gene model described by Eschrich *et al*. (43). RSI was computed for each sample using voom-normalised gene expression values for the RSI gene set (AR, JUN, STAT1, PRKCB, RELA, ABL1, SUMO1, PAK2, HDAC1, and IRF1). Within each sample, expression values for these genes were rank-ordered, and the resulting ranks were combined using the published linear regression coefficients to generate a continuous RSI score. Higher RSI values indicate a more RR transcriptional phenotype. RSI distributions were compared between RR and RS groups within each cancer type using Wilcoxon rank-sum tests, and effect sizes were summarized using median differences.

### Overall survival analysis

Overall survival (OS) was evaluated in patients with annotated radiotherapy response, with RS defined as complete or partial response and RR defined as radiographic progressive disease or stable disease based on post-treatment clinical outcomes. Pan-cancer survival data were obtained from the TCGA Clinical Data Resource (44), with OS defined as the time from diagnosis to death or last follow-up and expressed in years. Survival distributions between groups were compared using Kaplan–Meier analysis and the log-rank test. Cox proportional hazards regression was used to estimate hazard ratios for radiotherapy response, with additional models stratified by cancer type to account for differences in baseline survival across tumour types. All analyses were performed in R using the *survival* and *survminer* packages (45,46).

## Results

### Cancer-specific genomic associations reveal phenotype-specific signatures and novel candidate loci of radiotherapy response

Among the 14 cancer types analysed, six demonstrated genetic variants significantly associated with either RR or RS, exceeding the Bonferroni multiple testing threshold. These included head and neck squamous cell carcinoma (HNSC), breast invasive carcinoma (BRCA), brain lower-grade glioma (LGG), lung adenocarcinoma (LUAD), pancreatic adenocarcinoma (PAAD), and prostate adenocarcinoma (PRAD). Under Benjamini–Hochberg FDR thresholds, a larger number of significant associations were identified across additional cancer types. While some overlapped with Bonferroni-significant variants, others did not, suggesting potential associations that may be missed under more conservative correction. In total, 132 significant variant associations were identified across these six cancer types (Supplementary Figure 1).

To further characterise the direction of these associations, volcano plots were generated for the six cancer types with significant variant associations. Distinct enrichment patterns were observed, with variants typically enriched in either RR (odds ratio approaching infinity) or RS (odds ratio approaching zero) cohorts (Figure 3). Although variants with finite odds ratios were identified across the broader set of tested somatic variants, the stringent Bonferroni correction mainly retained associations exhibiting complete phenotype separation. Consequently, many significant variants were observed exclusively within either the RR or RS cohort, resulting in zero cells within the corresponding contingency tables. Under these conditions, Fisher’s exact test yields odds ratios of zero or infinity, reflecting complete separation within the analysed cohort rather than a precisely quantifiable effect size. Accordingly, odds ratios were interpreted primarily as indicators of the direction of association rather than the magnitude of effect.

**Figure 3.**
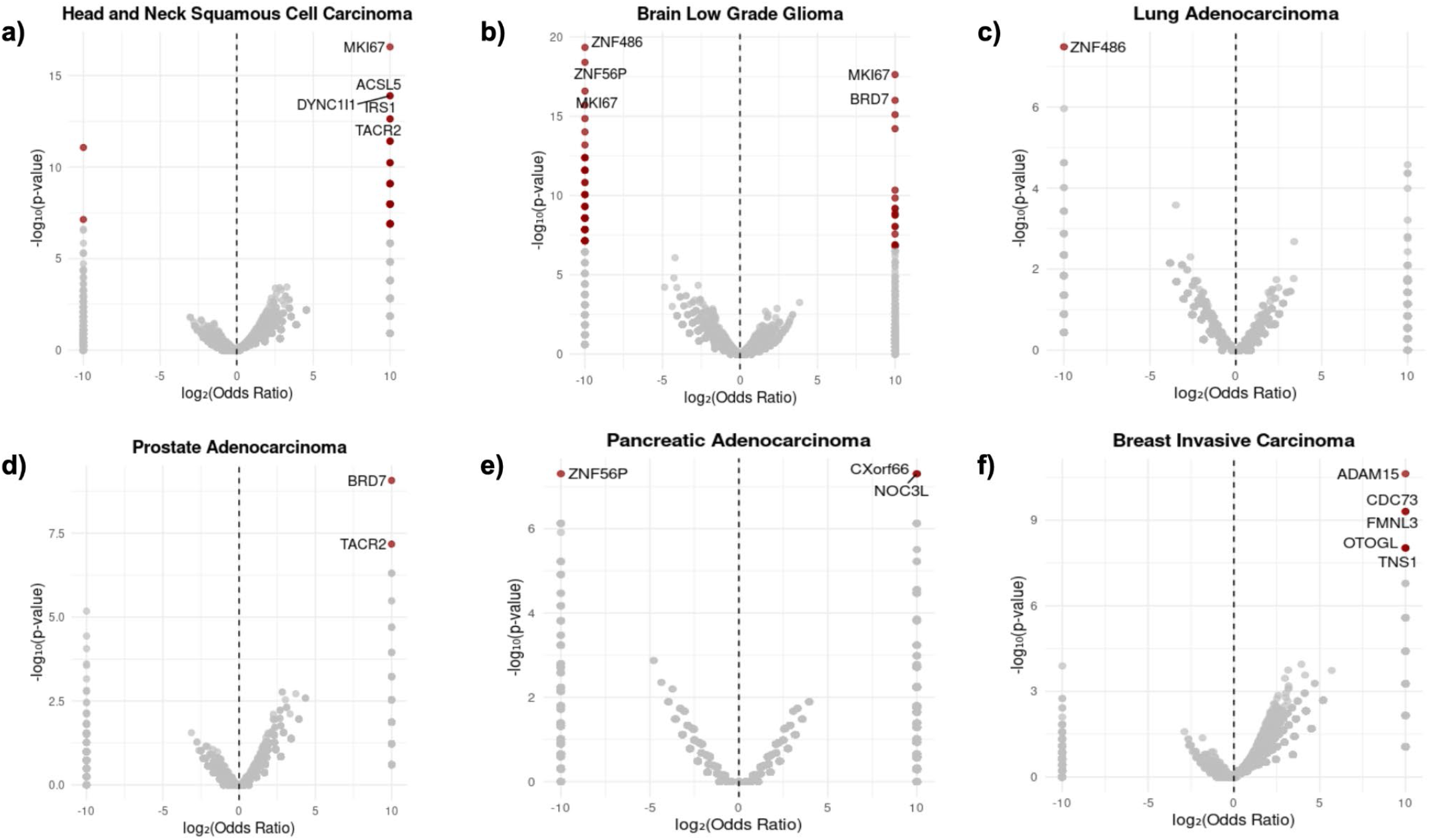
Volcano plots displaying direction of association with radiotherapy response across the six cancer types with significant variant associations. Variants surpassing the Bonferroni-corrected significance threshold are highlighted in dark red. Gene symbols are annotated for the most significant variants in each cancer type (top five for HNSC and LGG). Positive log₂(OR) values indicate enrichment in RR samples, while negative values indicate enrichment in RS samples.

Closer inspection of significant loci further identified several indels occurring within short repeated DNA sequences, where multiple distinct insertion and deletion alleles can arise within the same genomic region. These repeat-associated indels were associated with opposing radiotherapy response phenotypes across different cancer types or histological subgroups and were therefore interpreted primarily at the locus and gene level rather than as completely independent variant associations. Supplementary Table 3 summarises loci shared across RR and RS phenotypes and across multiple cancer types. For example, variants affecting MKI67 were identified in both LGG and HNSC, while multiple indel variants affecting BRD7 were observed across adenocarcinoma and LGG cohorts. Other variants displayed clear phenotype-, cancer type- , or histology subtype-specific enrichment, including variants affecting PRSS3, PRDM12, RBM25, IRS1, TNFSF9, ASTN2, PASD1, KRT10, TCF20, MAGI1, and PTPN14 (Supplementary Table 4).

Among the significant associations identified, a subset of variants was absent from dbSNP, ClinVar, and COSMIC, representing previously unannotated candidate loci. Following consolidation of overlapping variant representations into a non-redundant set, three novel coding variants were identified and are reported in Table 2. These included in-frame deletions affecting TCF20 (chr22:42214566, AAGGAGG→AAGG), PTPN14 (chr1:20857326, TTGTG→TTG), and CADM1 (chr11:115209591, ATGGTGG→ATGG). No matching sequence-resolved indels were identified in dbSNP, ClinVar, or COSMIC for any of these events. The affected genes have established roles in transcriptional regulation (TCF20), growth control and signalling (PTPN14), and cell adhesion and immune-related processes (CADM1), highlighting them as candidate loci associated with radiotherapy response (Table 2).

**Table 2:** Novel variants identified in this study. Moderate-impact variants absent from dbSNP, ClinVar, and COSMIC. Phenotype-specific novel variants were identified in a single radiotherapy response phenotype, whereas the second category comprises distinct repeat-associated indel alleles at the same locus associated with different response phenotypes. All variants were in-frame deletions and are presented as genomic coordinates (chromosome_position) followed by the reference → alternate allele.

| Variant | Gene Symbol | Cancer Type<br>(Phenotype) | Functional Annotation |
| --- | --- | --- | --- |
| **Phenotype-Specific Novel Variants** |  |  |  |
| Chr22_42214566<br>AAGGAGG → AAGG | Transcription Factor 20<br>(TCF20) | Adenocarcinoma<br>(RS) | Positive regulation of transcription by<br>RNA polymerase II |
| Chr1_20857326<br>TTGTG → TTG | Protein Tyrosine<br>Phosphatase Non-receptor<br>type 14 (PTPN14) | LGG (RR) | Roles in lymphangiogenesis, chromatin<br>remodelling, protein dephosphorylation,<br>and regulation of proliferation |
| **Novel Variants at Loci Represented in Both Response Phenotypes** |  |  |  |
| Chr11_115209591<br>ATGGTGG → ATGG | Cell Adhesion Molecule 1<br>(CADM1) | Adenocarcinoma<br>(Both) | Involved in immune response (NK cell<br>activity, cytokine production), apoptosis,<br>and cell adhesion |

### Radiosensitivity is associated with nuclear regulatory programmes, whereas radioresistance is linked to metabolic and cellular adaptation

Functional enrichment analyses revealed distinct biological themes associated with RS and RR. DAVID functional annotation clustering demonstrated that RS-associated genes were significantly enriched for nuclear regulatory processes, including chromatin organisation, transcriptional regulation, and DNA-binding transcription factor activity (enrichment score [ES] 0.98–1.82, best raw p ≤ 0.002; Table 3). In contrast, RR-associated genes showed enrichment for cytoplasmic and protein-regulatory functions, including cytoplasmic localisation, phosphoprotein status, and protein binding, although these associations did not remain significant following multiple-testing correction (ES = 0.30, raw p = 0.20). Collectively, these findings suggest that RS is characterised by gene regulatory programs, whereas RR is associated with metabolic and cellular adaptation functions.

**Table 3:** Functional annotation clusters for genes associated with RR and RS. Clusters are labeled by the primary functional theme of enriched Gene Ontology (GO) or UniProt Keyword (KW) terms. Enrichment Score (ES) reflects the mean –log10(raw p-value) of all terms in the cluster (higher ES = stronger overall enrichment). The best raw p-value represents the most significant individual term in the cluster, while Bonferroni-Šidák and FDR values account for multiple testing. Clusters with high ES and low adjusted p-values indicate strong enrichment.

| Cluster (Functional Theme) | Representative Terms (GO/KW) | Number of Genes | Example Genes | Enrichment Score (Best p-value) |
| --- | --- | --- | --- | --- |
| **Radioresistance** |  |  |  |  |
| Cytoplasmic & Protein-Regulatory | Cytoplasm, Phosphoprotein, Protein binding | 4 | KRT10, ASTN2, IRS1 | 0.30 (0.20) |
| **Radiosensitivity** |  |  |  |  |
| Nuclear Transcriptional Regulation | Transcription regulation (KW-0805), Transcription (KW-0804), Positive regulation of DNA-templated transcription (GO:0045893), Chromatin (GO:0000785), Nucleoplasm (GO:0005654), DNA-binding (KW-0238) | 6 | ZMYND8, MBD3, GZF1 | 1.82 (0.002) |
| Transcription Activators | Positive regulation of DNA-templated transcription (GO:0045893), Activator (KW-0010), Positive regulation of transcription by RNA polymerase II (GO:0045944) | 4 | MBD3, FOXJ2, NCOA6 | 1.60 (0.002) |
| DNA-binding Transcription Factors | RNA polymerase II cis-regulatory region DNA binding (GO:0000978), DNA-binding transcription factor activity, RNA polymerase II-specific (GO:0000981), DNA-binding (KW-0238) | 3 | GZF1, FOXJ2, IRF9 | 0.98 (0.08) |
| Metal/Zinc-binding Proteins | Zinc-binding (KW-0862), Zinc ion binding (GO:0008270), Metal ion binding (GO:0046872), Metal-binding (KW-0479) | 3 | ZMYND8, GZF1, ALPP | 0.61(0.14) |

Pathway-level enrichment analysis using Metascape further supported this distinction, revealing a prominent metabolic and cellular adaptation signature within RR-associated gene sets (Figure 4). Adenocarcinoma-derived RR gene sets exhibited the strongest and most extensive enrichment, with over-representation of pathways related to cellular organisation, metabolic regulation, detoxification, and stress-response processes. In contrast, HNSC demonstrated a more restricted enrichment profile largely confined to immune- and regulatory-related pathways, while LGG, LUAD, PRAD, and PAAD showed limited pathway-level enrichment. Although the broader enrichment observed in adenocarcinoma-derived gene sets may partially reflect increased statistical power associated with larger cohort sizes, the enrichment themes remained biologically coherent across response-, cancer type-, and histology-based analyses.

**Figure 4:**
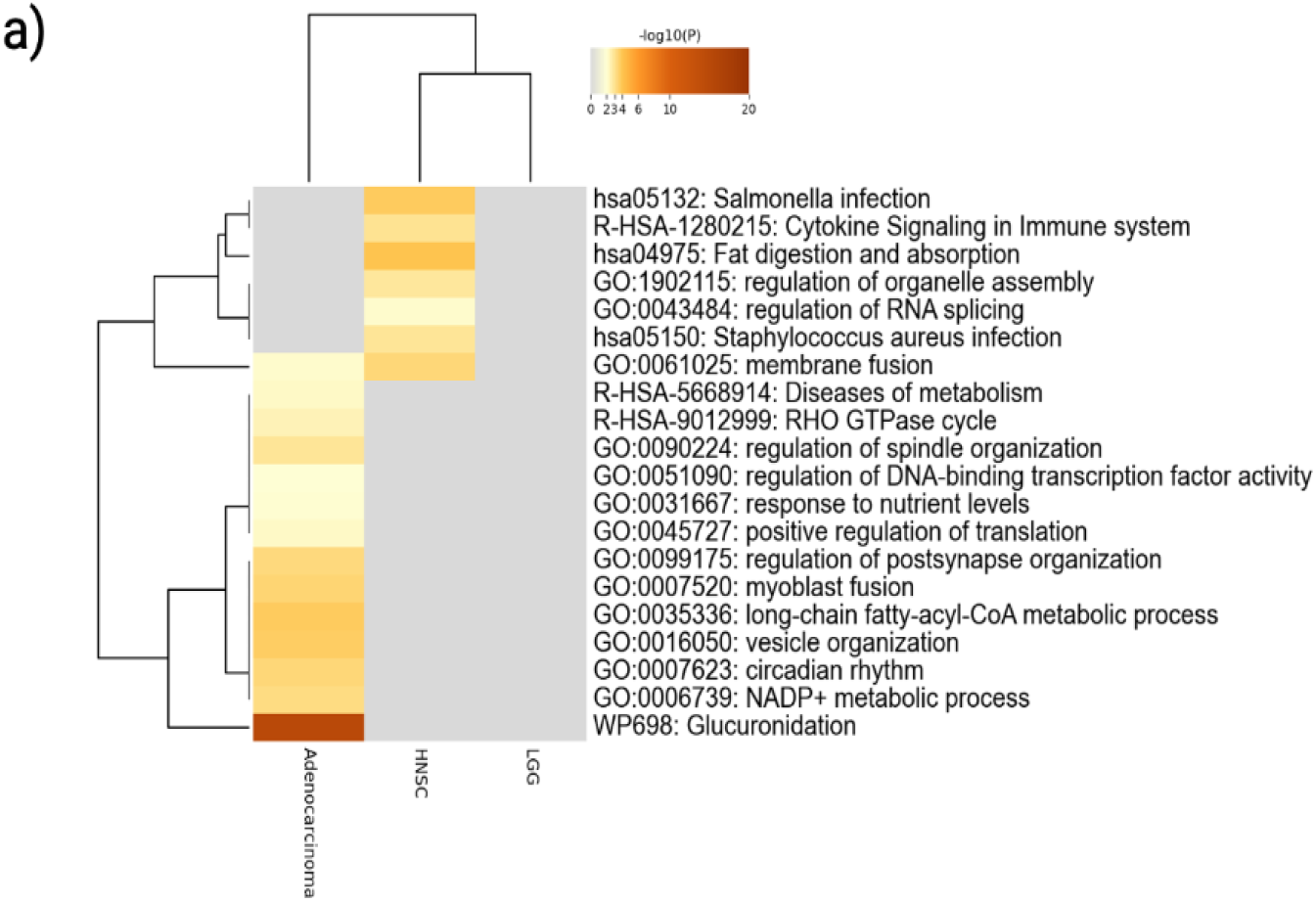
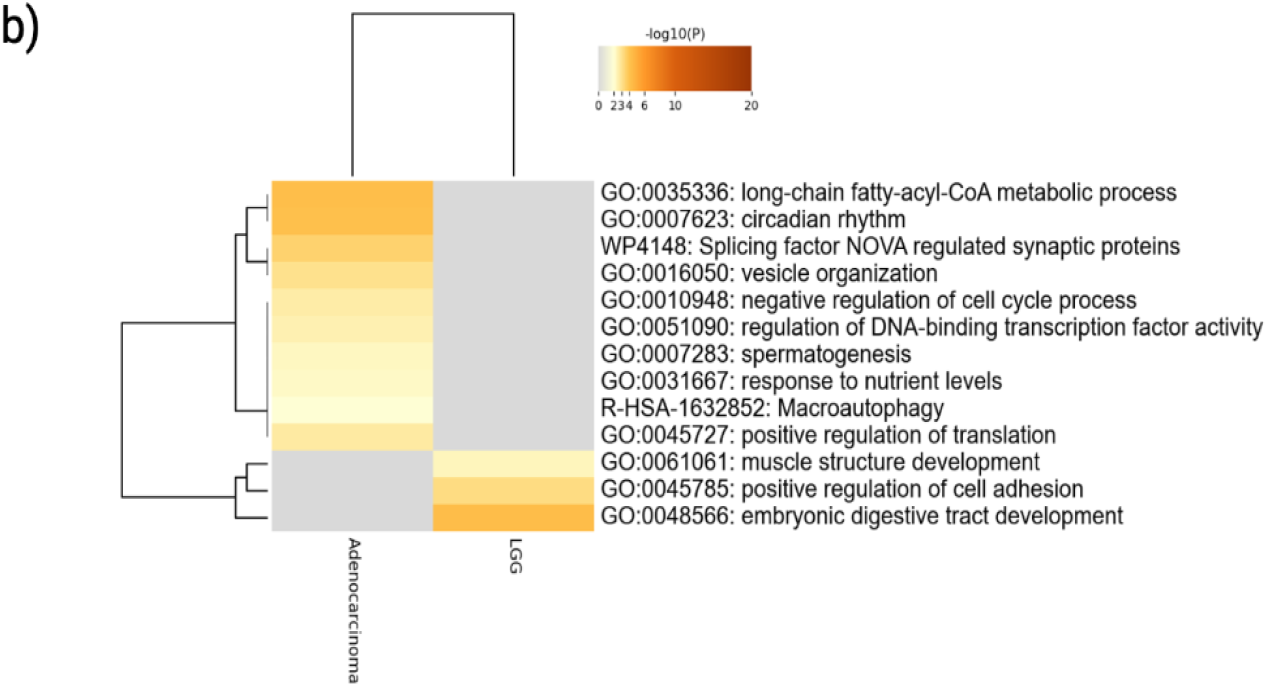
Functional enrichment heatmaps of RR- and RS-associated gene sets. (a) Heatmap showing enriched biological pathways and processes derived from RR-associated gene sets and (b) RS-associated gene sets. Heatmaps were generated using Metascape and displayed significantly enriched terms following multiple-testing correction using the Benjamini–Hochberg false discovery rate (FDR) method. Rows represent enriched functional terms, and columns represent gene lists. Colour intensity reflects enrichment significance, with darker shading indicating stronger statistical enrichment. Only gene lists that yielded statistically significant, non-redundant enrichment and met minimum gene-count thresholds are shown.

PPI analysis provided further evidence for the prominence of metabolic and cellular adaptation programs in RR (Figure 5). Two MCODE-defined interaction modules were identified within the RR network. The dominant module comprised members of the UGT1A gene family together with CYP2S1, forming a densely interconnected enzymatic core enriched for glucuronidation and xenobiotic metabolic processes. A second, smaller module comprised BRD7, MBD3, ZMYND8, HECW2, and PC, representing a heterogeneous network associated with chromatin regulation, protein turnover, and broader cellular regulatory functions. In contrast, RS-associated genes formed only a small interaction module consisting of CXorf66, RSRC1, and NCOA6, genes involved in transcriptional and RNA regulatory processes, with no large coherent interaction networks identified. Together, these analyses distinguish RS by enrichment of gene regulatory processes, whereas RR is characterised by prominent metabolic and cellular adaptation programs.

**Figure 5.**
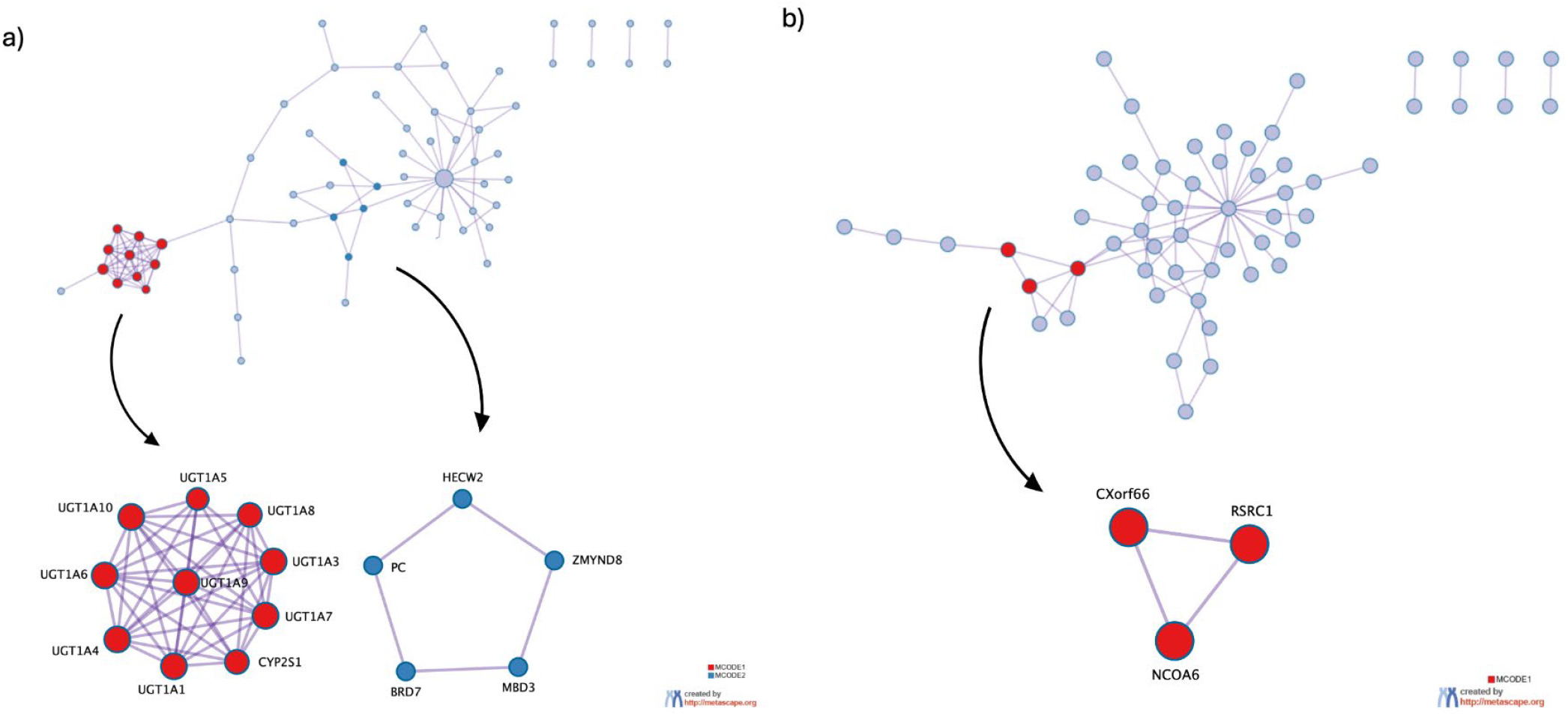
PPI network analysis of radioresistance- and radiosensitivity-associated gene sets. (a) PPI network constructed from adenocarcinoma-associated RRgenes, showing the full interaction network (top) and two densely connected modules identified by MCODE (bottom). (b) PPI network constructed from a merged radiosensitivity-associated gene set across tumour types, shown as the full interaction network (top) with zoomed-in MCODE-identified interaction module (bottom). PPI networks were generated using Metascape, and interaction modules were detected using the MCODE algorithm. Networks are shown only for gene sets that yielded sufficient interaction density to support robust network inference. Node size reflects connectivity, and edges represent known or predicted protein–protein interactions.

### Transcriptomic analyses reveal extracellular signalling and tumour–microenvironment programs associated with radioresistance

Differential expression analysis identified transcriptional differences between radioresistant (RR) and radiosensitive (RS) tumours across eight TCGA cancer types: THCA, BRCA, HNSC, CESC, PRAD, STAD, UCEC, and SARC. Using an adjusted p-value threshold of <0.1, 107 differentially expressed genes (DEGs) were identified, encompassing protein-coding genes, non-coding RNAs, and pseudogenes. Fifteen genes demonstrated stronger statistical support (FDR < 0.05) across four cancer types (STAD, BRCA, UCEC, and CESC; Figure 6). STAD contributed the largest number of DEGs, followed by SARC. Notably, with the exception of H2AC12 and IGFBP6, all identified DEGs were upregulated in RR tumours. Principal component analysis did not reveal clear separation between RR and RS groups, likely reflecting limited cohort sizes (Supplementary Figure 2).

**Figure 6:**
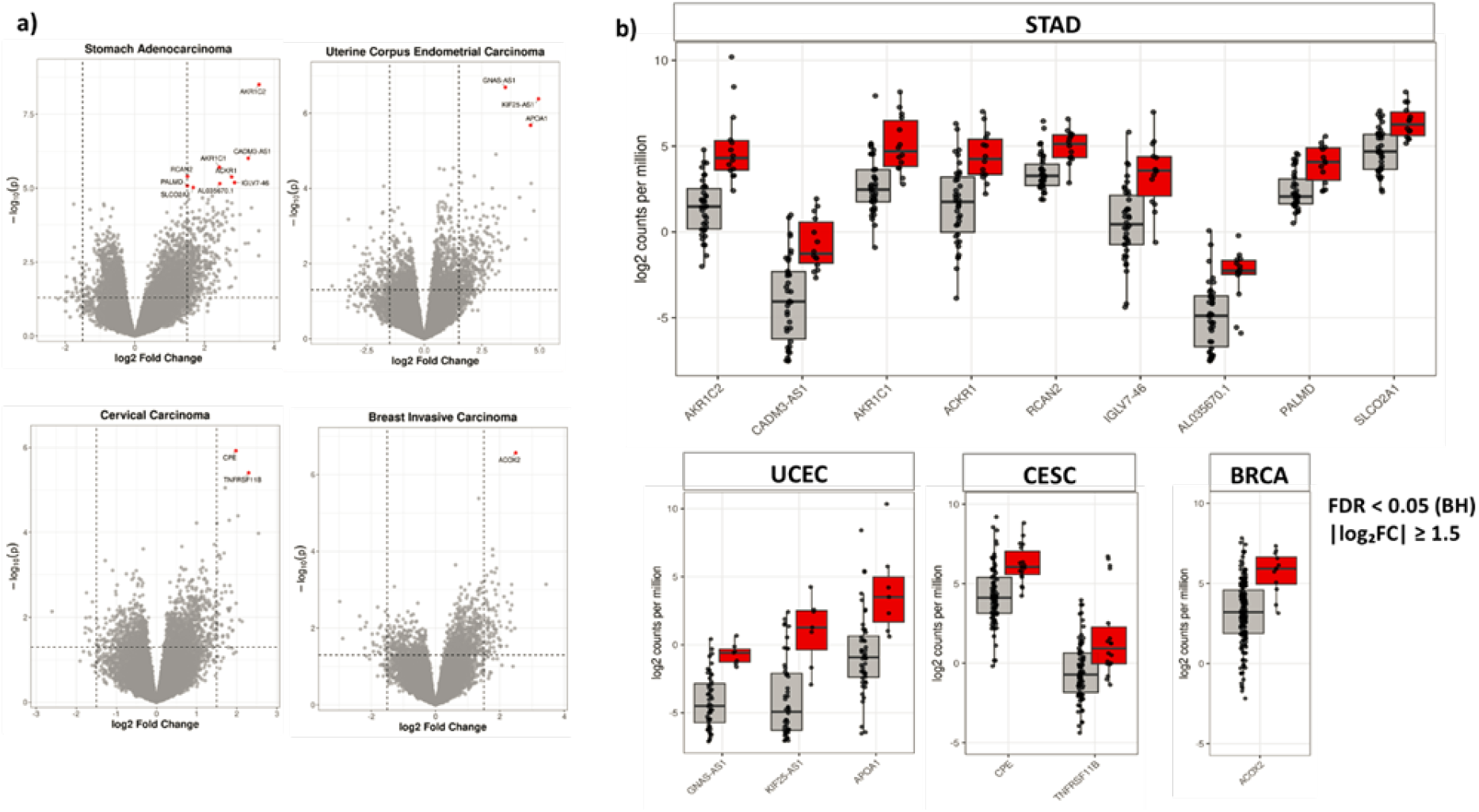
Differential gene expression in radioresistant patients. (a) Volcano plots showing differential gene expression between radioresistant (RR) and radiosensitive (RS) patient groups for cancer types exhibiting significant transcriptional differences (STAD, UCEC, CESC, and BRCA). Each point represents a gene, plotted by log₂ fold change (RR vs RS) on the x-axis and –log₁₀ adjusted p-value on the y-axis. Genes meeting the significance threshold (|log₂FC| ≥ log₂(1.5) and FDR ≤ 0.05) are highlighted in red, while non-significant genes are shown in grey. (b) Box-and-jitter plots showing log₂ counts-per-million (log₂ CPM) expression values for the significant DEGs highlighted in panel (a). Expression levels are shown for radiosensitive (RS; left, grey) and radioresistant (RR; right, red) tumours, illustrating the direction and magnitude of differential expression between the two groups. Statistical testing was performed using empirical Bayes–moderated t-tests within the limma framework. Per-gene log₂ fold changes, raw p-values, and FDR-adjusted values are provided in the Supplementary Table 5.

Gene Ontology enrichment analysis further supported the prominence of extracellular communication programs in RR tumours (Figure 7b, 7c). DEGs were significantly enriched for cellular component terms associated with the cell surface and extracellular environment, including extracellular region, extracellular space, cell surface, and cell periphery. To investigate the functional relationships among these genes, protein–protein interaction analysis was performed on 78 protein-coding DEGs using STRING and Markov Cluster Algorithm (MCL) clustering (Figure 7a). The largest and most interconnected module contained HGF, IGF1, IGFBP6, RSPO1, TNFRSF11B, MGP, and RERG, representing genes involved in extracellular signalling, growth factor regulation, and cell surface-associated processes. A second module comprised extracellular matrix- and complement-associated proteins, including COL14A1, OGN, GDF10, C1QTNF7, and C1QTNF9, implicating matrix remodelling and microenvironment-associated pathways. A third module contained genes involved in neuroactive ligand–receptor and stress-response signalling, including ADRB2, TACR1, PENK, RCAN2, and CPE. Although many DEGs originated from STAD due to the larger number of transcriptional changes detected in this cancer type, the resulting clusters demonstrated strong functional coherence across tumour types. Together, these findings indicate that radioresistance is characterised by coordinated upregulation of extracellular signalling, growth factor networks, and tumour– microenvironment interactions, suggesting that adaptive communication between tumour cells and their surrounding environment represents a common transcriptional feature of the radioresistant phenotype.

**Figure 7:**
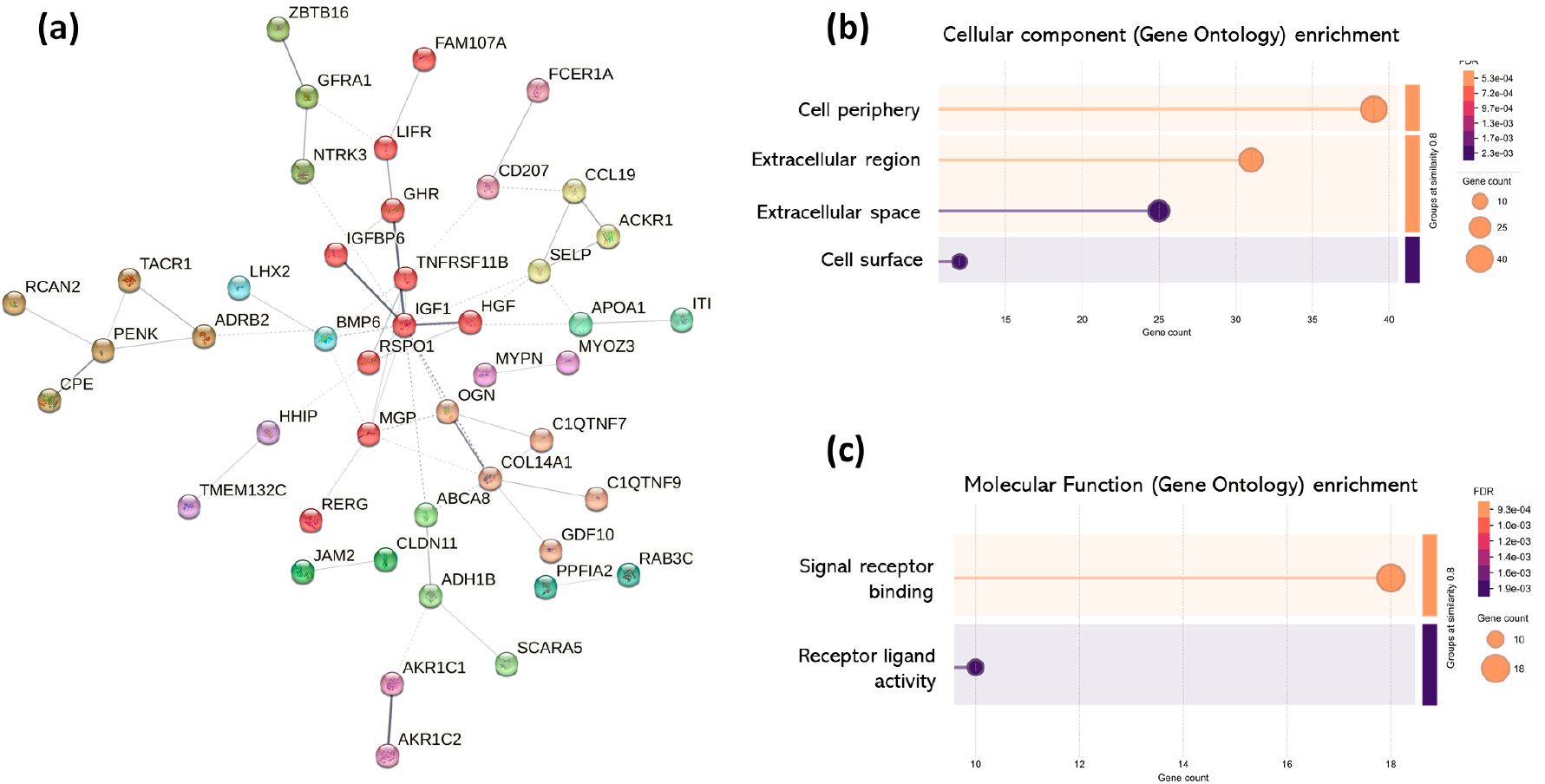
Network clustering and Gene Ontology enrichment of genes overexpressed in radioresistant patients. (a) Protein–protein interaction network of 78 differentially expressed genes (DEGs) annotated using DAVID. Network edges represent STRING interaction confidence, with line thickness proportional to the strength of supporting evidence. Disconnected nodes were excluded from visualization. Markov Cluster Algorithm (MCL) clustering was applied to identify natural network modules based on stochastic flow, with each colour denoting a distinct cluster. Functional enrichment of network clusters based on cellular component (b) and molecular function (c) was assessed using an FDR threshold < 0.05, with enrichment strength and signal values > 0.01.

### Genomic and transcriptomic analyses reveal pathway-level convergence despite limited gene overlap

Although genomic and transcriptomic analyses identified largely distinct gene sets, enrichment analyses indicated greater convergence at the pathway level. Overlap between genes implicated in somatic variant association analysis and the DEGs was predominantly driven by non-coding rather than coding variants (Figure 8). RR non-coding genes shared nine enriched pathways with the DEG set, compared with two shared pathways for RR coding genes, three for RS coding genes, and one for RS non-coding genes. The overlap between RR non-coding genes and DEGs comprised pathways related to hormone and steroid responses and metabolism, whereas RR coding genes shared pathways associated with cell adhesion and cell migration. RS coding genes likewise showed overlap across multiple cell adhesion-related processes, while RS non-coding genes shared only the alcohol metabolic process with the DEG set. Although cell adhesion-related pathways were enriched in both RR and RS coding gene sets, the limited overlap at the individual gene level suggests that distinct molecular alterations within these pathways may contribute to divergent radiotherapy response phenotypes.

**Figure 8:**
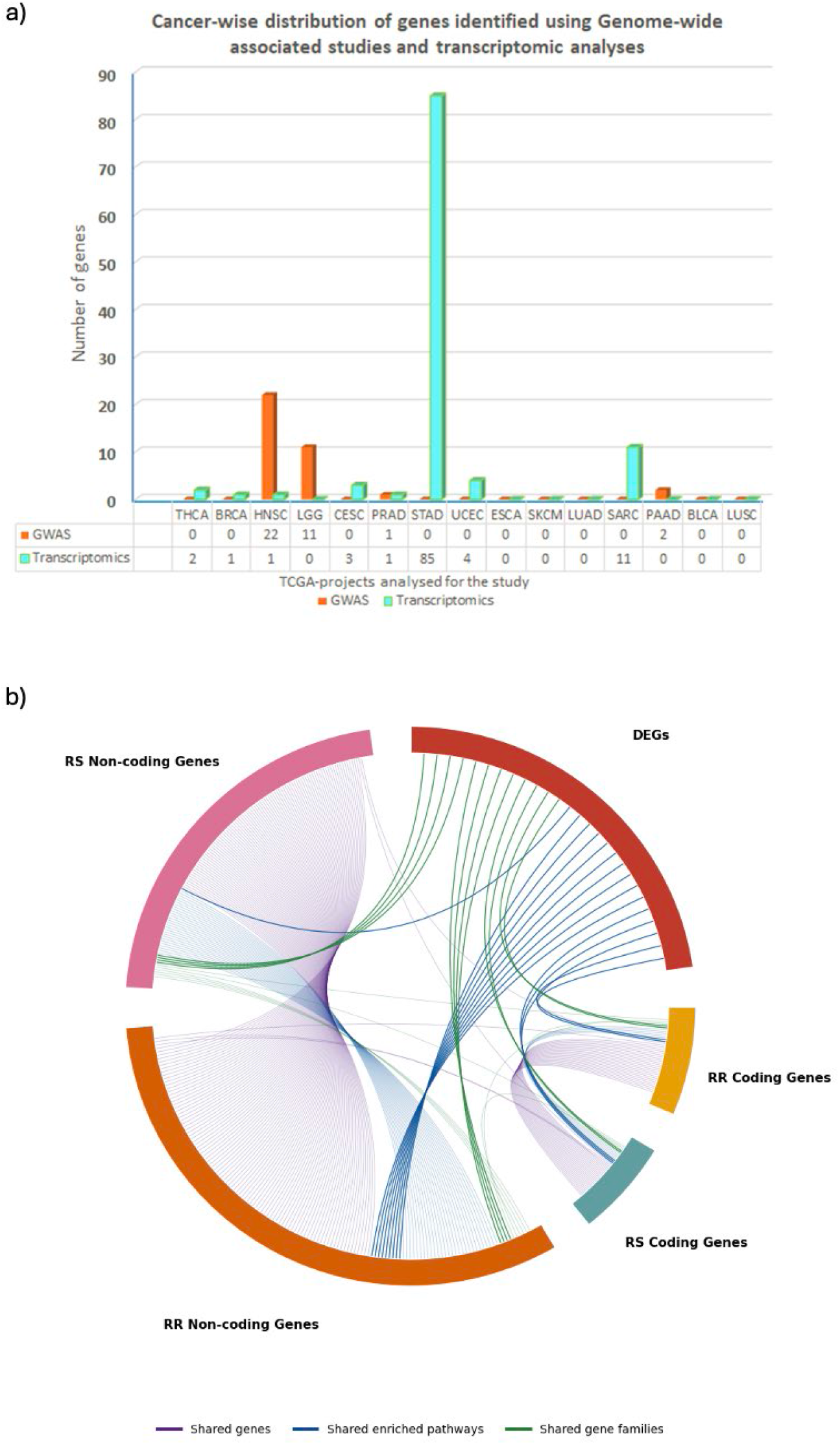
Distribution and overlap of genes between somatic variant association analysis and transcriptomic signatures of radiotherapy response. (a) Bar plot showing the number of candidate genes identified across TCGA cancer types from GWAS (orange) and transcriptomics (cyan). The figure highlights cancer-specific variability in detected candidates, with stomach adenocarcinoma (STAD) contributing the largest number of transcriptomic signals. In contrast, GWAS-derived candidates were predominantly observed in head and neck squamous cell carcinoma (HNSC) and lower-grade glioma (LGG). These patterns reflect differences in cohort size, statistical power, and molecular signal strength across tumour types. Gene count details are provided in Supplementary table 1. (b) Circos plot illustrating overlap between DEGs and genes associated with coding (high- and moderate-impact) and non-coding (low- and modifier-impact) variants identified in RR and RS tumours. Purple lines represent shared genes, blue lines represent shared enriched biological pathways, and green lines represent shared gene families. Darker lines indicate overlap involving the DEG set, whereas lighter lines indicate overlap between genomic gene sets only.

Although no individual genes were shared, related members of several gene families were identified across the transcriptomic and genomic gene sets. Notably, members of the zinc finger (ZNF) family were represented across the DEG, RR coding, RS coding, RR non-coding, and RS non-coding gene sets, highlighting recurrent involvement of this transcription factor family in radiotherapy response. Given the established roles of ZNF proteins in transcriptional regulation, genome stability, chromatin remodelling, and DNA damage repair [47], together with their emerging association with radiotherapy response across multiple cancer types [48–51], this recurrent representation suggests that disruption of ZNF-mediated regulatory mechanisms may constitute a common molecular feature underlying radiotherapy response. Beyond the ZNF family, the DEG and both RR and RS non-coding gene sets shared members of the tumour necrosis factor receptor (TNFRSF), tachykinin receptor (TACR), and voltage-gated potassium channel (KCN) families, indicating broader convergence on receptor- and signalling-related gene families. In contrast, convergence between the DEG and coding gene sets in both response phenotypes was more limited, with overlap observed in the astrotactin (ASTN) family, represented by ASTN1 and ASTN2, and the transcription factor (TCF) family, represented by TCF23 and TCF20. Collectively, these findings suggest that genomic and transcriptomic alterations associated with radiotherapy response converge primarily at the level of biological pathways and functionally related gene families rather than individual genes, with the strongest genomic–transcriptomic concordance observed for non-coding rather than coding alterations.

RR-associated gene sets exhibited greater pathway- and gene-family-level concordance with the transcriptomic data than RS-associated gene sets. While this may indicate stronger genomic– transcriptomic convergence in RR tumours, the larger RR cohort may also have increased the power to detect significant associations and contributed to the greater degree of overlap observed.

### Clinically defined radioresistance is associated with poorer survival but limited concordance with surrogate markers

Differences in tumour immune composition between RR and RS tumours were generally modest across cancer types following adjustment for tumour purity (Figure 9a). Most purity-adjusted effect sizes were very small in magnitude, indicating that radiotherapy response status was not associated with large-scale shifts in immune cell composition. While individual immune populations differed in specific tumour contexts, no consistent pan-cancer immune signature of radioresistance was observed.

**Figure 9:**
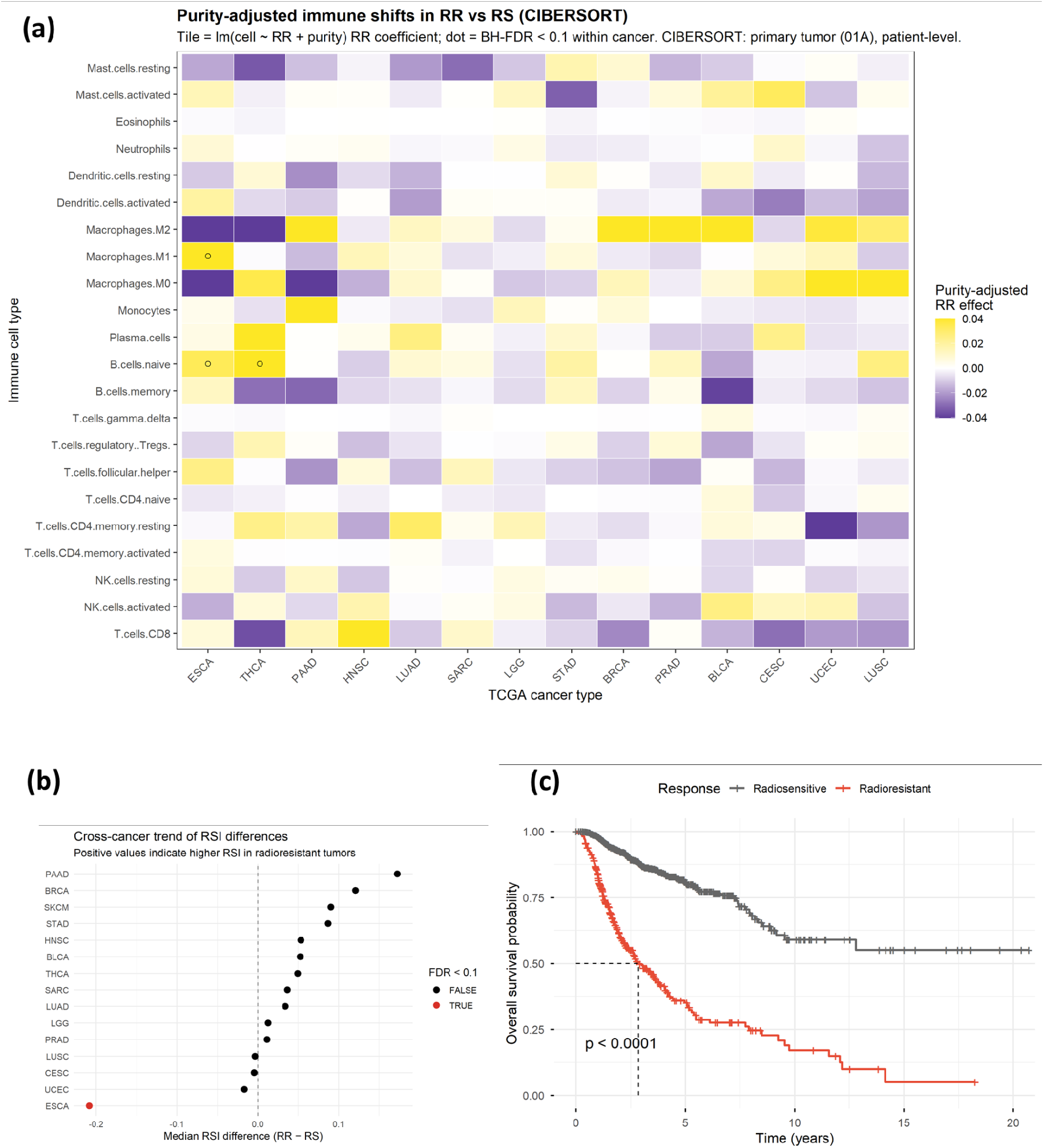
Multilevel characterisation of radioresistance: RSI, immune contexture, and survival. (a) Heatmap showing differences in tumour immune cell composition (estimated using CIBERSORT (LM22 signature)) between radioresistant (RR) and radiosensitive (RS) tumours across TCGA cancer types. Each tile represents the difference in median immune cell fraction between RR and RS tumours within a given cancer type, calculated as median(RR) − median(RS). Yellow indicates enrichment in RR tumours, while purple indicates enrichment in RS tumours. Dots indicate immune cell types with Benjamini–Hochberg FDR < 0.10 within each cancer type. (b) Forest plot showing the median RSI difference between radioresistant (RR) and radiosensitive (RS) tumours across cancer types. Positive values indicate higher RSI in RR tumours. The dashed line denotes no difference (RR − RS = 0), and the red colour dot indicates significance after FDR correction. (C) Kaplan–Meier curves showing overall survival for radioresistant (RR, red) and radiosensitive (RS, grey) patients across the radiotherapy-treated TCGA cohort. RR tumours exhibit significantly worse survival compared with RS tumours (log-rank p < 0.0001). Dashed lines indicate the median survival time for the RR group, while the RS median was not reached within the follow-up period. Tick marks denote censored observations. The statistical summary is provided in supplementary table 6.

Similarly, the RSI showed limited concordance with clinically defined radiotherapy response (Figure 9b). Across most cancer types, median RSI values were comparable between RR and RS tumours or showed only modest, non-significant increases in RR cases. An exception was observed in esophageal carcinoma (ESCA), where RR tumours exhibited significantly lower RSI values than RS tumours, contrary to the expected direction, since higher RSI is typically associated with greater radioresistance. These results suggest that clinical radioresistance in TCGA cases may not be adequately captured by intrinsic radiosensitivity estimates derived from RSI.

In contrast, clinically defined radioresistance was strongly associated with inferior survival outcomes (Figure 9c). Among RS patients, 83.57% were alive at the time of analysis compared with 48.45% of RR patients (Supplementary figure 3a). Median overall survival was approximately three years for RR patients, whereas RS patients demonstrated substantially longer survival, with median survival extending beyond 10 years. No significant associations were observed between radiotherapy response status and demographic or clinical variables. Collectively, these findings demonstrate that clinically defined radioresistance is associated with substantially poorer overall survival despite limited concordance with commonly used surrogate markers, including tumour immune composition and RSI.

## Discussion

Radiotherapy remains a cornerstone of cancer treatment, yet direct associations between molecular features and radiotherapy response remain underexplored, as most radiogenomic and multi-omic studies infer RR or RS from surrogate endpoints such as overall survival, gene-expression signatures, or TME characteristics (44,52–57). Studies using direct measures of radiotherapy failure are comparatively rare and are typically restricted to cell-line models or small, cancer-type-specific patient cohorts, limiting cross-tumour generalisability (58–60). To our knowledge, this is the first pan-cancer study to examine molecular correlates of radiotherapy response using annotated patient treatment outcomes, rather than inferring from surrogate markers. This distinction is important, as the biology underlying radiotherapy failure in patients is not necessarily equivalent to that associated with poor prognosis more broadly.

By leveraging clinically annotated treatment-response data across 15 TCGA cancer types (Table 1), our study provides a pan-cancer assessment of molecular features associated with RR and RS based on observed treatment outcomes. We identified 132 significant somatic variant associations across six cancer types (Supplementary Table 2), 107 differentially expressed genes across eight cancer types, and three unannotated, coding variants in TCF20, PTPN14, and CADM1 (Table 2). Although the genomic and transcriptomic analyses yielded largely distinct candidate genes, both converged on biological processes consistent with adaptive survival in RR tumours. Clinically, RR cases were associated with substantially poorer survival but showed limited concordance with commonly used surrogate markers, including RSI and tumour immune composition, highlighting the value of direct treatment-response annotations for radiogenomic discovery (Figure 9).

In our cohort, the dominant signals were not classic universal DNA damage-response genes alone, but a combination of cell-intrinsic metabolic and translational adaptations at the genomic level and extracellular, secretory, and microenvironment-facing programs at the transcriptomic level. Although this approach was constrained by the limited availability of radiotherapy response annotations, with clinical RT data available for only 13% of TCGA cases, the signals identified were biologically coherent and consistent across multiple analytical layers. The consistent emergence of metabolic, protein-regulatory, and broader cytoplasmic adaptation processes across gene-, pathway-, and network-level analyses suggests that successful tumour survival following radiation exposure may depend on the capacity to coordinate metabolic and post-transcriptional adaptive responses to radiation-induced stress. While these signals spanned pathways involved in detoxification, lipid metabolism, and cellular maintenance, the most prominent interaction network was centred on UGT1A family members and CYP2S1, suggesting that glucuronidation-related processes may represent one component of this broader adaptive phenotype (Figure 5). More broadly, the cytoplasmic bias of the RR genomic signature suggests that post-transcriptional and translational control is an important component of resistance, allowing rapid adaptation when transcription is impaired after irradiation.

RR-associated patterns observed here suggest a model in which cytoplasmic control of protein abundance and activity contributes to tumour survival following radiation exposure (Table 3). This interpretation is consistent with growing evidence that deregulation of translation initiation factors acts as an active driver of tumour cell survival, stress adaptation, metastasis, and angiogenesis rather than as a passive component of the gene expression machinery (61). Cytoplasmic stress-buffering mechanisms can rapidly reshape the proteome to favour DNA repair, apoptosis avoidance, and survival without reliance on de novo transcription. In glioblastoma stem-like cells, radiation-induced changes in protein synthesis are largely uncoupled from transcriptional responses, highlighting translational control as a major determinant of RR (62). Together, these observations support the concept that RR can be sustained through cytoplasmic translational reprogramming and non-canonical modes of translation initiation, consistent with the cytoplasmic enrichment trends observed in our RR gene sets (63). In contrast, RS gene sets exhibited enrichment for nuclear transcriptional regulation at the gene and pathway level but formed only small, weakly connected interaction modules at the network level. The comparatively sparse connectivity of RS networks is biologically expected, as transcriptional regulators typically engage in transient, context-dependent interactions rather than forming stable enzymatic complexes (64,65).

Several candidate genes identified in this study map to pathways that have previously been implicated in radiotherapy response, supporting the interpretation that these findings reflect underlying biological processes rather than stochastic variation. The enrichment of a novel in-frame deletion in TCF20 among RS adenocarcinoma cases is consistent with its established role in transcriptional regulation and chromatin-associated control of gene expression (66). Likewise, the identification of a novel PTPN14 variant among RR brain lower-grade glioma (LGG) cases is biologically consistent with established models of glioma radioresistance involving dysregulation of the Hippo–YAP pathway (67–72) (Table 2). Together, these observations suggest that individual candidate loci identified in this study align with broader radiobiological mechanisms previously linked to treatment response.

Notably, PTPN14 also harboured a distinct variant associated with RS, indicating that its influence on radiotherapy response may be context dependent. Previous studies have similarly demonstrated that the effects of some molecular pathways on radiotherapy response can vary according to the broader biological environment in which they operate. Hypoxia-inducible factor-1 (HIF-1), for example, has been reported to promote either radiosensitisation or radioresistance depending on tumour context and treatment conditions (73). A similar pattern was observed for CADM1, which was implicated in both RR and RS adenocarcinoma samples. Together, these findings suggest that radiotherapy response is shaped not only by the presence of individual genetic alterations but also by the broader molecular and signalling environment in which they occur, highlighting the context-dependent nature of RR and RS phenotypes.

The transcriptomic results added a distinct but complementary layer to the genomic results. Significant DEGs were entirely distinct from loci harbouring significant somatic variants. This indicates that the transcriptome is not simply a downstream readout of the mutational landscape. DEGs also showed no overlap across cancer types, indicating that transcriptional regulation associated with RR are largely tumour-type specific. We also found no association between DEG numbers and sample size, RR/RS cohort balance, or the tumour types with the strongest clinical resistance, negating a simple dataset-composition effect, suggesting RR is driven not only by steady-state transcriptional shifts but also post-transcriptional regulation, lineage-specific wiring, or microenvironment.

Despite this marked gene-level heterogeneity between cancers, the transcriptomic analyses converged on a clear RR-associated programme enriched for extracellular, secretory, and cell-surface functions. This shifts the interpretation of clinical RR beyond cell-intrinsic damage repair alone and toward active remodelling of the TME after irradiation. Such an adaptation may be effective because it allows resistance to be distributed across the tissue rather than borne by individual tumour cells alone, enables rapid and reversible responses during fractionated treatment, and recruits protective inputs such as stromal support, vascular stabilisation, cytokine buffering, and immune modulation that are not directly encoded in tumour DNA. In other words, clinically manifest RR may depend as much on the ability to build a protective ecosystem as on the ability to repair damage within the cell.

The transcriptomic network architecture across cancer types suggests that RR is mediated not only by tumour-intrinsic stress tolerance, but also by active reprogramming of extracellular signalling after irradiation. The HGF/c-MET and IGF centred module is consistent with a ligand-driven survival circuit that sustains viability during the acute post-radiation period (74,75). Through activation of PI3K/AKT and MAPK pathways, these signals can attenuate apoptosis, preserve metabolic function, and support DNA damage recovery, thereby increasing the probability that irradiated cells remain clonogenically competent (76,77). The extracellular matrix and complement-associated module points to a complementary stromal adaptation in which matrix remodelling, altered adhesion signalling, growth factor sequestration, and inflammatory conditioning collectively create a more radioprotective niche (8,78,79). Rather than indicating effective antitumour immunity, this pattern is more consistent with a wound-repair-like microenvironment that buffers tissue damage and facilitates tumour persistence (79,80). The neuroactive ligand-receptor and stress-response module suggests an additional adaptive layer mediated through GPCR and neuroendocrine-like signalling. These pathways are well positioned to transduce radiation-induced stress into rapid changes in calcium flux, cAMP signalling, secretion, vascular responses, and immune modulation (8,79–83). Thus transcriptomic RR is not simply a reflection of intrinsic resistance at the single-cell level, but of a broader ecological response in which tumour cells exploit stromal, paracrine, and stress-signalling networks to maintain survival and restore growth after treatment.

Taken together, the genomic and transcriptomic results suggest that these data capture distinct but complementary components of the resistant phenotype. The genomic layer points to intrinsic stress-handling capacity, including detoxification, lipid handling, translational control, and broader metabolic buffering, whereas the transcriptomic layer reflects the adaptive state deployed in tissue context, characterised by extracellular communication, ligand-dependent survival signalling, stromal and niche remodelling. Clinical treatment failure is therefore likely depends not only on fixed tumour-cell properties, but also on dynamic interactions with the surrounding microenvironment.

These are not redundant findings. Rather, they suggest that resistant tumours survive radiotherapy by combining internal metabolic resilience with active use of the surrounding tissue environment. This integrated framework also explains why there was limited overlap between mutated genes and differentially expressed genes: the former may establish a permissive background for survival, whereas the latter represent the adaptive state through which that survival advantage is realised in vivo. From a translational perspective, this is useful because it prioritises two classes of potential targets: cell-intrinsic metabolic dependencies and extracellular signalling circuits for validation in <u>radioresistant</u> cell lines, organoid systems, and tumour–stroma co-culture models.

Further, comparison with commonly used surrogate measures of RR underscores the value of treatment-defined response and our findings. RR cases were associated with substantially poorer overall survival and shorter median survival, suggesting that radiotherapy failure has clinically meaningful consequences for patient viability and that survival-based measures may provide a useful surrogate in the absence of direct treatment-response annotations, albeit with important limitations. By contrast, RSI and inferred tumour immune composition showed only limited concordance with clinically defined response. This is notable because RSI has been widely used as a surrogate of intrinsic radiosensitivity and to inform radiation dose modelling; however, our patient-level data suggest that it does not fully recapitulate the biology of radiotherapy failure in vivo. More broadly, these findings indicate that patient-level radiotherapy outcome is not adequately captured by fixed expression-based radiosensitivity scores or broad immune deconvolution metrics. One possible explanation is that the dominant transcriptomic signals associated with RR in our cohort were extracellular, stromal, and paracrine, features that are only partially represented by existing surrogate frameworks. However, these approaches may still be informative in specific contexts, but they should not be assumed to reflect the biology of treatment failure without direct clinical validation.

Importantly, the present analysis primarily captures intrinsic determinants of radiotherapy response, reflecting baseline genomic and transcriptomic features present prior to treatment exposure. Acquired radioresistance, by contrast, is a dynamic process that emerges during or following radiation treatment through adaptive molecular and cellular responses (84–86). While such treatment-induced mechanisms cannot be fully resolved using baseline tumour profiles alone, intrinsic molecular architecture is likely to influence the trajectory of subsequent adaptive resistance. These observations highlight the need for longitudinal multi-omic studies that integrate pre-treatment and post-treatment tumour profiling to fully characterise the evolution of radiotherapy resistance.

### Limitations

Several limitations should be considered in interpreting these findings. The number of tumours with radiotherapy response annotations was modest, and the uneven distribution of RR and RS cases across cancer types reduced statistical power and may have contributed to unstable effect-size estimates. Genomic associations were tested using Fisher’s exact test to accommodate sparse variant counts, but this approach did not permit adjustment for clinical covariates such as age and sex. In addition, many significant associations were driven by rare somatic variants observed exclusively in either RR or RS tumours, resulting in complete phenotype separation and extreme odds ratios. These features are inherent challenges in analysing somatic treatment-response phenotypes in cancer-type-stratified cohorts and mean that individual loci should be regarded as candidate signals rather than definitive biomarkers until validated in larger independent datasets.

The transcriptomic analyses are also limited by the use of bulk tumour RNA, which does not allow extracellular and secretory programs to be assigned unambiguously to malignant, stromal, or immune compartments, and by the limited granularity of TCGA treatment records, which precluded detailed modelling of radiotherapy dose, field, fractionation, and concurrent systemic therapy.

Despite these constraints, the recurrence of coherent biological themes across orthogonal analyses argues against these findings reflecting stochastic variation alone. Instead, they support the value of directly analysing clinically annotated radiotherapy outcomes as a biologically distinct complement to surrogate-based radiogenomic approaches. Larger and more comprehensively annotated cohorts will be needed to refine these associations, support multivariable modelling and derive more robust treatment-response signatures. Such datasets will also be essential for prioritising the candidate genes and pathways identified here for functional testing in radioresistant cell-line models and, ultimately, for identifying actionable vulnerabilities that could be exploited to overcome tumour RR.

## Supporting information

Supplementary images

## Data Availability

The TCGA data analyzed in this study are available through the NCI Genomic Data Commons (GDC). Controlled-access somatic VCFs were obtained following authorization through dbGaP and are available to qualified researchers through the appropriate dbGaP/GDC access process.

https://portal.gdc.cancer.gov/

## Acknowledgements

The authors would like to acknowledge Drs. A. Venkitaraman, A. Karumbati and N. Kamariah for useful discussions that led to the conception of this project. The authors would like to acknowledge Drs. M. Ibrahim and A. Ravindran for a critical reading of the manuscript.

