## Supplementary images for "Direct Pan-Cancer Multi-Omic Analysis of Radiotherapy Response Reveals Coding and Regulatory Genomic Variation, and Extracellular Adaptive Programs Underlying Radioresistance"

### Supplementary Section

#### Supplementary Figure 1: Distribution of Variants by Significance Across Cancer Types

*Manhattan plots illustrate the genomic distribution of variants across chromosomes by association p-value. The significance threshold was defined using the Bonferroni multiple-testing correction. Among the 14 cancer types analysed, six exhibited variants surpassing this threshold. Panel (a) shows 49 significant variants from 167 participants in HNSC, (b) 71 variants from 165 participants in LGG, (c) 1 variant from 49 participants in LUAD, (d) 2 variants from 70 participants in PRAD, (e) 3 variants from 28 participants in PAAD, and (f) 6 variants from 197 participants in BRCA. Variants with identical p-values overlap in the plots.*

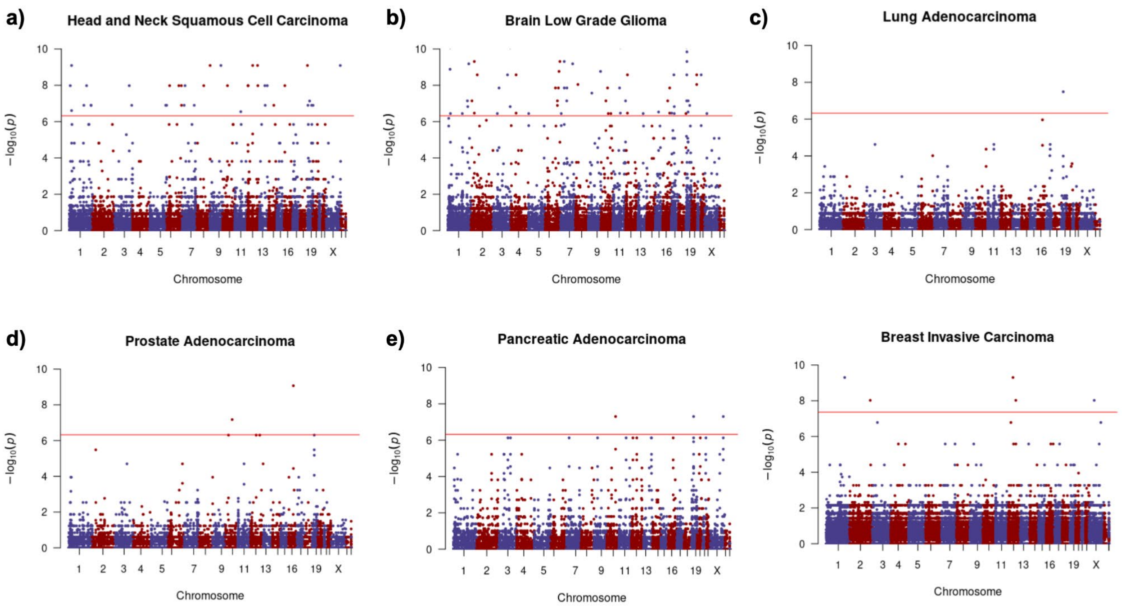


#### Supplementary Figure 2. Principal component analysis of gene expression profiles across cancer types.

*Principal component analysis (PCA) plots showing the distribution of tumor gene expression profiles from radiosensitive (RS) and radioresistant (RR) cohorts across 15 cancer types: THCA, BRCA, HNSC, LGG, CESC, PRAD, STAD, UCEC, ESCA, SKCM, LUAD, SARC, PAAD, BLCA, and LUSC. Each point represents an individual tumor sample. RS samples are shown as red circles and RR samples as black circles.*

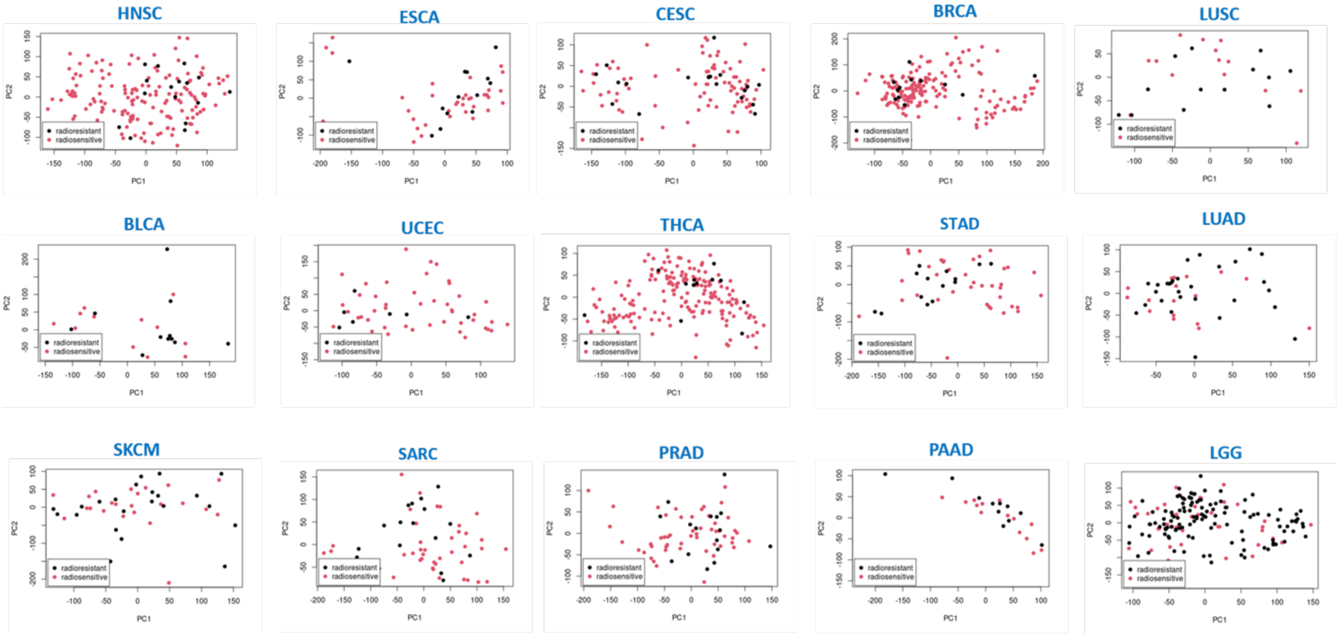


### Supplementary Figure 3. Vital status and overall survival according to RT treatment best response

*a) Bar chart showing the distribution of vital status, expressed as the percentage of patients alive or deceased in the radiosensitive (RS) and radioresistant (RR) cohorts. b) Kaplan–Meier survival curves showing overall survival probabilities according to radiotherapy treatment best response: stable disease, radiographic progressive disease, complete response, and partial response.*

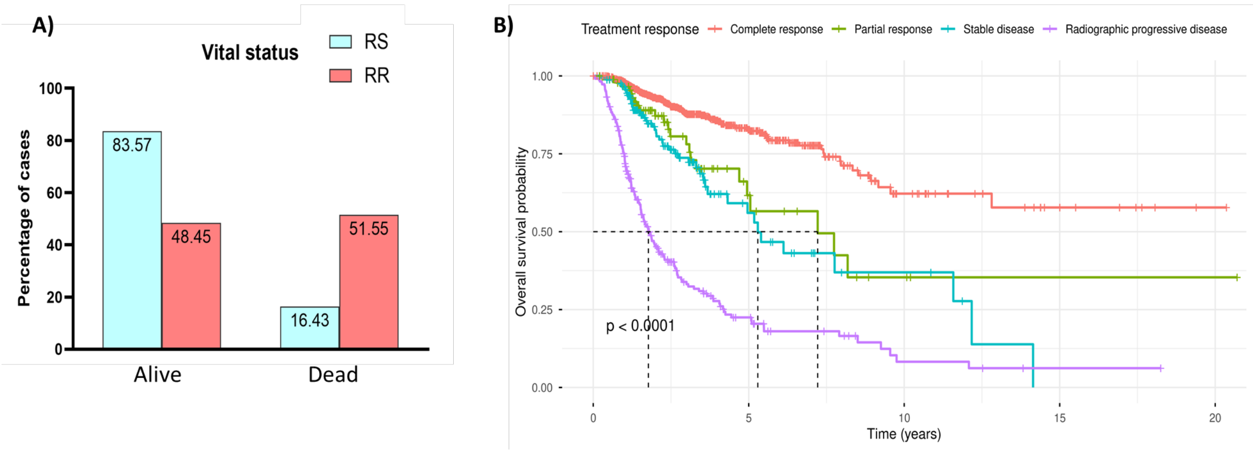

#### TCGA Cancer Types analysed in our study

| | |
|---|---|
| BLCA | Urothelial Bladder Carcinoma |
| BRCA | Breast Cancer |
| CESC | Cervical Squamous Cell Carcinoma and Endocervical Adenocarcinoma |
| ESCA | Esophageal Carcinoma |
| HNSC | Head and Neck Squamous Cell Carcinoma |
| LGG | Brain Low-Grade Glioma |
| LUAD | Lung Adenocarcinoma |
| LUSC | Lung Squamous Cell Carcinoma |
| PAAD | Pancreatic Adenocarcinoma |
| PRAD | Prostate Adenocarcinoma |
| SARC | Sarcoma |
| SKCM | Skin Cutaneous Melanoma |
| STAD | Stomach Adenocarcinoma |
| THCA | Thymus Cancer |
| UCEC | Endometrial Carcinoma |
