## Supplementary table_1 for "Direct Pan-Cancer Multi-Omic Analysis of Radiotherapy Response Reveals Coding and Regulatory Genomic Variation, and Extracellular Adaptive Programs Underlying Radioresistance"

| bcr_patient_uuid of all RR and RS cases used in the study | | |
|---|---|---|
| **Radiosensitive_bcr_patient_uuid** | **Radioresistant_bcr_patient_uuid** | **Common_bcr_patient_uuid** |
| 0153F141-625E-4623-9F8A-296678002C63 | 019c5de9-74fd-48b7-95a3-227532e16c5a | 0394060D-010E-405F-983D-DB525F01F2C3 |
| 0167CF11-74BE-4701-AB9A-4E057D4BB545 | 029DA211-9B52-45AD-B0DC-2322DA783C9C | 084AC674-F4CF-4A4A-A6A2-687C05C272EA |
| 1C095C4A-DF97-402E-902C-FC83BCF8EF88 | 06580CBC-D0D8-4B97-87EF-EBC17CDAE81C | 1BE375FE-5AE5-4E75-A729-69F86A661D9A |
| 1C848577-C3A4-4830-ABFA-549992172396 | 0a017f15-1c6b-45e7-8d55-e0a71df1b2e8 | 1C89DA99-9E89-4726-B196-97EEAB46D504 |
| 1ccd29c1-cf30-4701-9b50-5d34cae2f4b3 | 10981CF9-C8E6-48C0-BC34-F06DB6E1AC28 | 2088AC24-A8B4-49AF-A954-2B78B88A6F66 |
| 1F5B6B07-45F5-4DA4-932A-DF9665F6C538 | 12d31787-e54d-4729-ace8-9141043c92b8 | 2B715EB2-055E-4BC5-BC98-128E8BF6954F |
| 239505CE-F2F1-477E-8AE2-B7065989BA79 | 159B47B2-DC7D-4856-9A00-454E27301E23 | 34AAD752-D78F-440B-ABD2-F9F939C82664 |
| 23C335F6-5749-4443-8AE8-61CCCBB137CB | 1DFB4565-81E1-4A1B-BB59-9B5629D6C447 | 40287CDC-19DD-4370-A6A0-F324960E1148 |
| 24ecbe19-a9ea-4215-a7f1-74300f9ed2a9 | 214a4507-d974-4b3e-8525-7408fccc6a0f | 57a2044d-20cc-4d02-b268-ce570a0fabe3 |
| 26293554-04D3-4FC9-B12E-6A199278ED11 | 27707915-D36E-4DA7-9B39-2FDD9CE0BDC5 | 5FE17DDE-EF75-49A2-90F8-0647077B6CA5 |
| 27286A38-1035-4A22-9EBC-874DF8C49026 | 2C96D3C4-323E-4A2A-9132-40E860B701A5 | 62088274-A45B-4616-B086-B0DDD64895F0 |
| 27D2B645-F1BF-4486-B0B5-273681B58625 | 2F6F86F3-6D31-4A8C-B70D-2759B44BAC6C | 6E22B287-5B45-4F5F-A825-B200AA6943E6 |
| 2A394996-809F-459F-A647-D209921CCBD2 | 3B076D77-6276-4233-867B-6A10D9E17343 | 785803BB-42A8-4AFB-B7B4-58C114894408 |
| 2C736C5C-9D32-4522-ACFD-6620BC291ACE | 3CF77E08-EA34-4091-B242-75C7E35FF699 | 7A211CE6-3162-4F58-BF24-9B68562913D5 |
| 2cd8ea73-cb3f-4b17-9b97-fa12eb03b85b | 45EEFBA8-9F7C-49E9-A3F5-D13CFA264FF6 | 8d5fe6e8-d2cc-481f-a6a1-9cb6e7982cde |
| 2E08A89E-4035-4782-B54F-F98EEF5DB80D | 4a032bad-e726-48f2-8f39-e3acc109cc91 | 909335D0-BB4B-4EA3-869A-93E4D91C2154 |
| 2EA7AD21-3324-4EDA-8C82-FC2CEEA2FC73 | 4A831893-F4CA-4357-A756-B5E954E35DD7 | 90BC0F31-0EC2-4CEB-AD99-6BE8F8B19180 |
| 306C53FB-640D-42AD-889A-ABC5FD3AD778 | 4D67B910-6681-45A4-9DAC-6A12D67287A8 | 9393E4F1-6A1B-4181-B928-72B6F335BEC2 |
| 315b8de8-9842-4738-b23b-1d61722f9583 | 4DA121DA-9193-4AC5-8206-03CB1F7C2C14 | 9583C10B-B21A-4863-98FA-61E735E64EA5 |
| 3203D0AE-5FE9-4FF2-BE0A-1D97F7684819 | 4E227A7A-B885-4E9B-A4C2-AAD9E6C154E5 | 9A8EA017-0152-48B2-ACCB-D040DEB1C6B6 |
| 3210027D-A9C7-4EBB-8497-E3705BED7E40 | 4E7F0FFF-7B17-4DA9-95EA-F68797C427F5 | ADFA6BEB-1AA3-4A2A-AFE7-8D036A80B20D |
| 345a375c-15cc-453a-8054-e68a35178082 | 57CF380D-AB52-467F-B9AE-740B2F84C820 | C07B122E-AC50-4DB2-ADD2-5617A5D0E976 |
| 34D4CC10-B868-4E85-96A5-8680FC729E34 | 602F2512-00B6-44B6-9ED6-F8B010224F8C | D02FFAA9-0403-4A89-BE01-CCE35C80DD69 |
| 35cb7841-9b09-465a-90c5-e3b8a9faad49 | 001944E5-AF34-4061-9C09-BB9EA346F6FD | DEBA32E4-0E68-4711-941B-3B63BD965AFB |
| 362000E4-CF7C-48FC-93CC-8F5994B442EA | 00397CE0-A931-46DD-9407-C1C6A14C39F1 | ECC74D54-D929-4DD0-B9EC-E03163343F9A |
| 02D98B0A-8360-49B9-BBD6-B70C79CBD32D | 0050D8BE-1DB6-4C17-8BEF-3AE2EAAA63CE | |
| 039788ec-1364-40c7-9e9d-13dc305674e9 | 00D588E3-3DB6-4CF3-9DEF-34D941E8CDB0 | |
| 0425CB44-66AF-4E25-AFCB-CACCCC2F1179 | 02661985-D13F-4007-9A42-5E08D1A6CA10 | |
| 051A8605-E674-4B66-A142-11EC36A96454 | 029CE650-5E5A-4100-8596-CD94300E7EF5 | |
| 0691B8C5-6244-407C-8601-FA1BDA01FF2A | 02f5ae33-a563-4ecb-9e33-dfa500a44931 | |
| 06fe7f3e-4edd-4150-981c-93787ea19b39 | 0314B145-2F3A-4A90-A580-375B5F0A63A4 | |
| 09918b81-4696-4cba-a45a-5fe7407c2337 | 035cb359-eac3-484a-9e88-3ddc739440a3 | |
| 0CC9A11F-1AC4-431D-8F94-52A3BC02B572 | 03804F9B-DF7C-462C-8984-8EB3A5ED4999 | |
| 0DB0F02F-94F5-492F-8840-5254792F6879 | 03aca47b-7653-4938-9178-ed7c37eee6d5 | |
| 0E7F79CD-D188-42EA-BF96-4BA9E9B8606B | 03C3AE62-D0AA-412E-BD3C-4577FC9F919C | |
| 0EFAF8F9-177C-4FFB-B03F-F0316ADF5DC1 | 045FE498-6A07-41FF-9956-DC0FB0483E7F | |
| 0F579238-5A11-4ADC-9C80-572CBB2CE58F | 048549CF-D0A5-4743-A0A1-2004F7CC2B08 | |
| 0FA89FBC-307A-48D5-ADCE-192556755C22 | 04fff86a-b6f5-4900-9e37-c1f0b881b160 | |
| 1068AD76-612E-40EE-B2FA-916DA14B7916 | 05026179-B1DA-411E-A286-89727B1AE380 | |
| 108DEE1E-B27D-4056-A167-261725F548C1 | 0557B788-5AED-4919-85EB-A503AD893D75 | |
| 109DF703-2865-4B4C-8E4B-BADFF8EB3F22 | 056386DB-D338-4FC8-8BB1-249BE72FD6CB | |
| 1422BC35-4597-4E76-AA75-8E79613E67D5 | 05A8268A-6B33-429B-8638-8C9BB366EF28 | |
| 1521ac6d-31f7-4240-9d54-4f7600e78e16 | 05fbd98c-28bd-41bc-ae60-16ac0c16c723 | |
| 15b0bfd4-2068-4ac8-aaa8-a2a6797253d8 | 061FAB24-727A-4551-A205-89EEB9F530EA | |
| 16182443-12A1-4B0F-A6AB-33C2942E2991 | 0625DF42-740F-4E52-8F9E-0F5B7DEE251B | |
| 16D3C1C1-CF3A-44B6-B564-8AE5464B243D | 065b26fc-6827-44d5-83d8-1100c63fcda6 | |
| 173174F8-9D95-40B0-9F88-A4BF4CCB521B | 06728B1B-6B69-4467-8E10-A8A4E6328AC8 | |
| 18d0aae7-9694-4252-b8fe-16e62dadab07 | 06EF6E83-5583-45B9-A0BD-EDEA68943083 | |
| 36470b30-a3b3-4d28-851b-918a7ff88011 | 0709DBCC-0828-4066-AA01-96F059E5FA97 | |
| 36F4BE75-CBCF-431B-9199-DBA295A905A1 | 07778870-D400-4495-B786-C63FF33FC29D | |
| 37884d39-64e8-4242-b6bd-4c0a1daa21cd | 077BDC90-1BBD-49EC-93A5-729B1AC1BA8D | |
| 39E147C9-2ACB-4BA8-A991-E43E1719B2D8 | 07A6BBCE-012B-48FE-8C62-E79A1B200EB6 | |
| 3C0BB5E2-AD33-4D91-A31E-2A18F1C94D2D | 07C805C2-FF09-4D5A-95F8-1D7D9F84C33C | |
| 3C1F8B73-BAD7-4649-B08E-C7AFC647EDF3 | 083CC988-19A5-49B4-97C0-4E7BCC5D5C08 | |
| 3CD2D4BD-8256-42FA-8272-DB379D598007 | 0858c8b7-e2eb-4461-b65e-9d476029ad8d | |

| | |
|---|---|
| 3DF1B9DB-D900-47D2-839D-3900829CB2A9 | 085FFCDC-8600-42E2-852A-506465F716A2 |
| 3E637872-F5E5-49D3-BB0D-9C16B8713382 | 0930E97A-72EA-4BD5-B2F0-50C76AF695F3 |
| 3F7B600B-282E-4F35-9FD8-567B3FCA8273 | 09A5E9FD-D816-4F8C-BAA9-0E40BA607B16 |
| 4048c139-1329-4e6a-8d2f-c0aa104a68ee | 0a11e958-7ec0-493c-bd7a-eb17536facd7 |
| 417dea5f-f68e-4dab-940e-43ae8c67e5e6 | 0a45f302-5748-48f3-9dc9-66c01843a68e |
| 42251e02-b687-4bf1-a1b6-e3dd978542e4 | 0AB29DE2-EB9E-4A25-99DE-5F543CFBDB53 |
| 4250AC3C-C8B0-46CE-B9C8-7963BC386041 | 0ABE0B8F-FD93-43E8-B78F-48A7C3F96868 |
| 42BCF1A7-2F3A-4ED8-A41D-CDD67E65B928 | 0ad2a2b3-c1bb-4597-b28e-a6385f93d09c |
| 43b99616-d7b4-404e-8912-4275f8d6dc55 | 0adf59c6-581a-475d-a2f4-40aa40060b5b |
| 45EEA556-E97B-4912-8AA9-7E5C709EB52B | 0aecac64-5982-4d76-8f31-958f6a00951d |
| 46625EBD-5BF3-4FE7-BF3F-19CC5A210555 | 0b1663fc-6a7e-4698-951d-a8c90fdf2411 |
| 47CB93BC-8F6B-4B2A-804C-BB4FC78DF9E1 | 0B6418FC-C775-4BAA-8776-74FFCC492E9A |
| 484B7B36-15DC-43A9-801D-580216A8CFCB | 0bb786ee-07cf-4853-9b49-ec95cd14e282 |
| 494A8495-8D10-4A8F-8664-F68AC339C889 | 0bc5744c-5fa3-45bb-87d0-70a02068b392 |
| 4996E458-DB04-4C6D-9AE6-66316240A8ED | 0BD52D75-5113-4EF1-BC75-509F59EAEF2B |
| 4A00CA75-D23A-4642-BEE8-DB4182517724 | 0C7B4B9C-D497-41FC-A2B9-014BB5E07985 |
| 4A60E87E-6BC1-4AE0-9CCB-4CDC3E99C608 | 0CAFEFEF-911B-46E8-9E6C-19A0E7D555AA |
| 4A95CE25-D06A-499D-B7EE-AFF5EE94D4F1 | 0d0731e2-8d82-4b6d-9258-2896a770a868 |
| 4b7cd595-e7f9-45a8-b736-a8c7c42d9539 | 0d0d83d9-d558-4d38-977b-6f1b2471beda |
| 4c9e6085-41c3-4cfe-ad3d-5f94196a86e0 | 0d497faf-2c1c-4173-a5fe-770cca73323c |
| 4CCCC34B-DAF6-41CA-9BF8-F1614525EAC9 | 0DD8DBC1-C48B-4E7C-B401-57101F724967 |
| 4F0124A6-4A5B-4E42-B9FF-D7DE51D2148C | 0DE19185-3517-4E30-925B-7EB1F5079EC2 |
| 4F4906DC-7EBD-47F1-A8F5-B35D3950E740 | 0E747C7E-3621-4D88-847C-86811655D908 |
| 4f786107-3cf5-4ab3-bba4-f399dee23f0e | 0EC70E40-07DF-461D-8FF9-351240A0D454 |
| 5134c56f-8286-4ec8-8348-237cee7dad5e | 0f25d7fd-fc7c-4563-a8bd-80b5e0c32ea2 |
| 514D12B0-6F60-406E-BDE9-A090E918EBFF | 0f4bb968-6e10-4a86-a03e-37b61d084f31 |
| 51DCBFAC-6F04-4364-877D-0BF1BD566335 | 0f718e17-86c1-4221-a878-d57ae87e744f |
| 53D99A89-6EBD-4637-91DB-7CE43EBC6B8B | 1005ccb2-10ef-4e37-950e-ad72393db03c |
| 53FBE99C-180C-45E6-8585-796028B3D118 | 1007195A-7B6A-4575-B6D1-075D76A997FF |
| 54a9cc9e-3eee-4caa-9f33-4ea64a2999ba | 1009A500-2A37-4AEE-B7C3-049B5CE961A4 |
| 5510ad32-62fd-4adc-964a-ae4e997c2f4b | 10290e91-9784-4f94-8daa-b5d9a8b0efb9 |
| 56D4CA9B-C050-45B4-B636-03FE2251FA1A | 10593E5B-7ACB-498B-BDF7-705E3E2BC443 |
| 5A2B2E1B-B631-40AE-BA41-D8022CDDB550 | 10BDAFBB-831D-48B5-B9A3-AD374FC50235 |
| 5B6C3188-5D33-4473-BDD4-0CB9671E9047 | 119EA204-792B-49EC-8FD4-EB88C4F91042 |
| 5C51FBFE-153C-4CD2-97BD-E444DAA0B20F | 1285EB55-415C-494A-AA58-936F0427CDD0 |
| 5D39E9A2-1073-4DB2-B02E-DB05D59563A7 | 12c27db5-db2f-48dc-a2ca-9557b951f43e |
| 603D2B61-B636-4054-AC95-AB66DABB3B48 | 12DA9527-CF33-46C6-90CF-65478FF15CF9 |
| 6594ACD5-4D2A-4CC5-8CF8-4064E252C5F6 | 12DF24C6-1229-4777-AD90-7DD66259B65F |
| 659668e8-f0d9-4ff2-bbc8-9246f2ef49ab | 134223BD-5C03-4854-A034-EBD2C527833E |
| 6668ACFE-1594-42C0-B839-8A592702675F | 13C97CFB-EEA2-4AE8-9209-C967D9642CEF |
| 6717B82E-889D-45A4-9638-4D0E023399CC | 13E936FF-81F0-49B9-BC2E-2EA43921F450 |
| 6813C53C-B256-4115-AD90-24D75328779E | 14407399-B1F0-46BF-9179-8BF11C39D919 |
| 699D3BA1-94C8-409F-90C3-E7FE2F1CF825 | 149B46A3-75DE-44E1-BA73-BDC4FF379C7F |
| 6BE3885C-219B-44F4-92BC-C055DC843A87 | 14DB56EF-EC5E-4775-A229-5D83E58E027C |
| 6C3C8219-74AB-4E54-81F5-84D4D5884252 | 158FD4C2-68DC-4CA4-8130-D5589C80D825 |
| 6EE03F14-14AC-4CB9-BD23-B69704176301 | 15E43EE8-3080-4895-84FB-A24AD3A68B97 |
| 70C6C3A0-B422-4541-A60F-355356A9A340 | 15EB8D88-ADB1-4F4F-A44D-087049DB60CE |
| 71B347AC-6AC6-475D-8E21-754764180948 | 15FF4B02-BB69-4D24-8A07-B82F428E428D |
| 71ed2681-79bb-4ffb-985c-6d3cf18e9168 | 1636242C-12DA-412E-A362-914A20ADD708 |
| 73D4E806-2C15-4C9E-A978-106FB9494E82 | 1654C42B-664B-407E-8CA7-D4CD908B709A |
| 7451AF3C-ACC0-4D79-8429-6B8BE96911D8 | 16d52afb-a954-4cb3-b168-c1501208f691 |
| 75dd635e-8e0c-4c0b-b21c-20f77770218d | 1757C89F-8795-4C3B-BBAD-1E6F617F2472 |
| 7746BE87-7AD0-48A7-84FB-08A81B220BDB | 17885905-280F-40FA-943E-346ED45403EA |
| 77472095-030D-4679-921D-8E8E217956C5 | 1791E250-70AC-439C-828B-15BA811935CC |
| 778DC00A-0B94-45A8-AE13-9C80F0C1DFD6 | 17a8d208-fcf6-4480-945c-848d4234aafd |
| 781f40c9-c099-4c96-8269-ebe2a449c93d | 17C0CE0F-8227-4119-9B39-EA8DB18F5F4B |
| 7883E417-A793-4614-9528-6C2E9E44379F | 17F63789-2EE3-4CD9-9273-5FA73662DB39 |
| 78C0D30C-0905-49DE-BE7F-AFC1337F59B7 | 18334157-AA38-4859-A786-99A920A1FBE3 |

| | |
|---|---|
| 79823022-7FEB-49DE-8DE7-607721B595D2 | 183dd089-e932-4be2-b252-0e8572e7da4e |
| 7A918706-C697-4CEF-9661-346C5A612FF8 | 189E1F27-7738-413A-A4D4-97D41D592A13 |
| 7ABACA8F-962E-4F9D-B63F-26C131BF5961 | 18E7B73C-4520-42CD-AC9B-23BAC1C8ECC6 |
| 7bcf6d42-eb8f-4aa2-825b-38671d4f9e3a | 19015644-9C17-4AE7-A656-0FE8623C04AE |
| 7EE2F8D5-6C31-4F5F-A779-B43979E66D0B | 1944E155-B765-48C3-9602-6787C105E551 |
| 818AD88A-131E-49DF-85D8-55BBA0E9EC02 | 197D01A5-321C-461E-9B45-B359288B959A |
| 81b7cbc1-c037-426f-95b0-d729a30697da | 1a1f2197-e303-46e4-8871-53f2eb2e599a |
| 823F571A-3CFD-465D-95FD-187A398CCF6F | 1A3D5555-7013-4F06-878A-98944F075B63 |
| 82476d2d-e403-4f6b-8dd6-cc84e3329478 | 1A4164D9-4484-452D-9382-8CBF896A870E |
| 83059A0F-4DE9-454C-8E51-9265AB5F3558 | 1A679332-30A3-4495-A2E5-39D299E14333 |
| 83A0F9EF-4BF0-4EFE-8713-F8B6AB4F5773 | 1ACDA00D-04FB-4D71-99A4-996F92601827 |
| 8504fd86-a70a-4cba-9ec8-25c9e60ca549 | 1ADDCD66-79AB-4976-9679-37F0F31BA1DF |
| 8655FA51-D784-4F8D-9BC8-F1DA7303D22F | 1ae087ac-63f0-4cf9-b698-f5c13e1f4681 |
| 865C624A-D52D-4C53-9904-40014CB8CCE7 | 1BACEBF3-609E-4BB7-BB93-EB217B3A1B5C |
| 87FABA0E-28E7-4A7E-A7E1-12582EC68FAC | 1BC03593-5AB6-4082-B157-7EA456845E28 |
| 897f85ec-7e14-426a-a532-eed01ccc90f1 | 1BCB1132-A9FC-4F03-BBED-ABAF80C7A24A |
| 8A94FF28-A74D-4FE2-B9C0-06C01F0A3FB6 | 1C801A6B-8407-46DA-B6CC-245F05418B9E |
| 8B10F00F-FDE6-423D-BC36-027931359A5A | 1CA54115-93A3-4C0E-ABF0-915B4BBFF10C |
| 8B9E81DA-EA22-4F6A-BDB1-281B0EC8A451 | 1CB1D2CA-DFA6-4C42-9DA9-C711DC5AAB5D |
| 8bb3ed57-6e48-4c90-bcf2-baf9a843398f | 1CC30842-4A5B-4F07-B36F-5B84CEAA5662 |
| 8D30C7BE-73B6-4C4C-80C5-31152B2F7CB0 | 1D38D356-D126-4476-94D0-26616B9375B1 |
| 8E44FB97-C649-4056-80AC-257CEF61D226 | 1E308B12-0590-4DAE-94D0-A539FCF25DF7 |
| 910655DA-2C12-4572-96B7-DED616E69B4B | 1E3BC429-50EB-4087-B39A-6D92F03412F7 |
| 917961E2-78AD-4182-9CDB-5EFB09808111 | 1EA575F1-F731-408B-A629-F5F4ABAB569E |
| 92654ad8-156c-4dd1-862e-dc339240d721 | 1EBD5834-56B8-498A-8F34-C0031FCFD856 |
| 93a265b7-6c23-4e5f-b797-c117793744bf | 1f0a5312-0590-4230-9da9-fd0cfe693586 |
| 93CBDA0F-8454-4127-A6DA-A19DDA345DEB | 1F13065A-40F7-455D-B8EB-F9A128722EAC |
| 95EDFABE-0B35-4CDD-AED4-E2234D806F35 | 1F9B1B71-8753-4CD5-A585-708BEA8FF1FA |
| 95fa64b0-91e1-4427-b81a-53c47a053563 | 1fe937f7-b4e5-4cf6-be9b-215a012533c6 |
| 962FF090-16C0-438B-8213-86B2BD306E4C | 20094E3E-CEE3-41DD-8553-1FC393117786 |
| 96D9249E-501C-49E0-9559-01DA978A9CA6 | 200F77A5-803C-4891-980B-5C3B08C91254 |
| 975E17C2-B93C-4D45-8A85-786F365E4A93 | 20280122-A66A-4C83-8F25-0B9D97558DC2 |
| 97f6c8d3-09a0-4128-8f10-41e736648a5f | 20ad4ac0-6a87-461f-83a5-fded51bf821a |
| 994347ba-3045-4ade-926f-06420fbd906c | 21254CB0-68F3-4ECC-8D9A-73FB0E994C5B |
| 99706AD3-5CF4-45F8-815E-51E78F2E9F79 | 2132F775-5D09-4B7F-B739-883B4AEFB264 |
| 9a50e7e4-831d-489f-87d2-979e987561cc | 214A09BB-CE52-4055-B036-065E8CB7AC7D |
| 9B161C41-4F76-4ED1-B60F-2B093BC70EDB | 21A29827-7896-436D-AB52-E7EDFC91E7D0 |
| 9b4eeecb-6aff-435b-a244-ec362af92b7f | 220D75AE-C94B-473A-A988-AC40C15FA219 |
| 9BF40EEB-9A87-44A2-A782-197AB954C937 | 22233A31-023F-412F-9B42-73B58C51DEE8 |
| 9D5C5303-550F-454F-8FD4-8ABCDD311215 | 227DAC7D-6BC3-4D8B-AC9C-EA4CAB9A602C |
| 9DD97821-7AFD-4982-8B1D-F14F845B79F7 | 22AE99B4-52CF-4D20-A068-7457E16919B6 |
| a02932f3-5606-42a5-aee4-e5b9f5aa1aaa | 22b35927-1c3f-4f98-ba42-deafc56b677a |
| A1066AEA-3122-4887-83D0-132EF84C06E1 | 233cb00f-b470-47fb-ad93-2a1597655ed6 |
| A1196D5F-8C7D-4D9D-9E46-C5C22A9C2FEF | 23451AB5-A32B-4EEC-9B8B-FD282282A0D4 |
| A1B99260-8643-4CEF-95A0-3853D64762B7 | 23592BC3-17FF-44F3-9B17-C8F21D0AF6B4 |
| a2fd0991-cfbd-4162-8ca9-2ed096f54a14 | 23CC4CD3-87DC-4342-9072-BE3695E393FA |
| A39AC229-60D2-42C2-9721-7E0894C0AE38 | 240EF385-DECC-48F7-A798-A81B4AB0410E |
| a3f6bc1c-19ab-4eeb-a9ac-3d2fac850bde | 24D7D4D8-F5C1-4D71-80E0-72A429FC9DBD |
| A6885883-F1A2-48DD-8D72-0038319069A1 | 2501ee46-8d38-448b-8765-e9c9706cbbe8 |
| A6975EB0-CFA6-4B7A-BBE4-72BBDD3D3B6F | 25184E51-1464-438A-80FF-CFC9A443CD36 |
| A6A07BC7-AB1B-429D-81ED-D7A890C88E51 | 2560FB5B-5F49-4D38-8F61-1AB911AED3EB |
| A80B0FAB-6189-497F-997F-77234B69179C | 256976A4-41AB-445E-AE26-C594BA1FA324 |
| A8F5C479-8685-4E2D-BB60-63F1CC651083 | 2610485E-F2B2-45F7-A578-99D6983D5F0E |
| AA6142AF-BAD7-493C-98A0-FD070DE39073 | 261C3D74-706E-4751-BD15-8F3C1A402FF0 |
| AAC385C1-42F9-4D9E-B9BF-94CA13EBC5AE | 26573441-eedb-4364-966c-e7f803deef19 |
| AB706EA0-D642-4F58-BA8D-F5D930B91286 | 26CB1330-CE48-4006-8155-F6D52D3AB9B6 |
| AB7EB3EC-BD87-4E49-9010-B51B17148F7C | 274225CF-A0F4-4635-848B-819F479001DE |
| ab8d7f84-dd71-42c7-9471-4db967a9c89c | 274496D6-0A9B-45CA-876F-B91F7277963C |

| | |
|---|---|
| B5B31686-995B-433A-AFAB-6EEE027817BA | 2779fa01-ac93-4e80-a997-3385f72172c3 |
| B5FA3AC1-0691-4B6A-B6E4-765311B419ED | 277b02e9-ded5-4980-845d-af53690000ac |
| B64A5F5A-5913-432E-9D08-0FF67A8C6B64 | 27953774-E97B-48E7-B966-1EC1E9C830B4 |
| b76d1de7-6447-474d-bf67-77e0bb2da73a | 27B529B8-0A7E-4A1A-9020-D33EE956C920 |
| B7AC2F06-012A-436E-AFDE-7E4D653C84D9 | 27ba717a-5cd8-440a-9345-4ae086f6efca |
| B865DEC4-F051-4FBE-9405-F832FF2010D7 | 286C69B0-75BA-48D6-A26B-26077F101BBB |
| ba3911e6-2208-4a04-8891-b5cba9fda6bf | 287059FB-4E69-4D6D-99D5-91EC92D35BDE |
| BCC17E9A-6FE3-44FF-963C-D2D850A431F1 | 28C50A5C-E0EA-4F93-B34C-B0A651264539 |
| bce3ce45-4fb3-4d8e-9ec7-d24427c2ba4d | 28D2FD8E-ABE9-4EA3-87E4-4F2B61A33C90 |
| BE73FDE2-D843-426B-9306-A02D3F2A754B | 2919AFA2-3FEE-43BD-AE68-E70718E73608 |
| BF0F1735-F94D-4B82-8F91-2FD2BAC80E32 | 2923e404-38f2-437a-b57e-23401fbe0273 |
| BF193C4F-FBAC-41BC-B1AF-19B6CE856F0F | 2983F08C-3B32-4FB3-9D7E-5B34F9C2F741 |
| BF3074E6-8780-490E-A8C3-4237C19CA877 | 29AFF186-C321-4FF9-B81B-105E27E620FF |
| c0e7fbe7-b9cb-4e65-941b-cd7d581c54ec | 29C4319C-3EAD-42B1-810D-418DA8D515C4 |
| c31b3adf-3fc6-4a72-83b0-bbc0f295b5e7 | 29df128d-ace2-40cf-88ee-3f6f2170f159 |
| C4060CA5-33E0-4C83-B91C-742D75059BDC | 2A398AF1-AEA5-4AAE-AB00-A2DC152F965A |
| C4F522C9-BFF5-4899-8D27-C105DAB795A9 | 2AE1706B-C0F7-40E3-AFE5-CB75D1584B96 |
| C512BA69-7B90-4334-92EB-48AB91AA05F2 | 2b22db1d-54a1-4b9e-a86e-a174cf51d95c |
| CAC8EABA-9E40-4048-B8CC-87574F048D91 | 2B794EAE-814C-4B5C-B276-BE9096B94F94 |
| CB695FDD-381E-4C00-836E-0BA27E32176B | 2C600A58-98ED-4572-9B6F-EDBA2719790C |
| CF29547A-305B-4E6E-8342-36AAC0850065 | 2C792B77-1BF1-42F7-BD94-73C6474C6C7D |
| CF332DAA-F335-4AF1-81DE-CBFDAFB0F764 | 2C878227-77A1-4988-A46C-885DB9E400F7 |
| d10a4887-373b-4726-a61d-126f950e4a06 | 2CF8DE9F-B305-4D98-BFD4-33377BF6138F |
| D3FF2AEB-46BE-4F54-8A8F-178EB921FBB6 | 2D166DC0-8890-4B00-AB73-9E2DBB6C530D |
| d7ab7ec0-3de7-4ffd-a5ac-f75579355b2a | 2D468C02-45F7-4B4B-920A-557AE6C91A43 |
| D9FD2724-7DB0-4AF3-AC14-217BDFA5203F | 2D633CED-AE85-462C-90E7-C1B109E084CF |
| DBC29B1B-42B8-4249-A1D1-3BA117EFEA43 | 2DCDF9A1-9437-4515-88C1-1C17D6B6418C |
| DCCD3854-8865-4452-9F1C-97283509B553 | 2e73e016-61ff-4f23-9ca9-fd48d9050d82 |
| DD056692-4E2A-4E45-A4D0-357AF9754F0E | 2ECCAABE-0783-4596-B674-D59846373A1D |
| dd46c83e-6551-485b-b935-f3beed891244 | 2F6A0E87-1E6C-41F3-93E0-3E505FA654B0 |
| DF27B584-FA45-4A8F-BD12-A018CC63DCA1 | 2fbafcc5-896c-4e50-8956-a61540fc89be |
| df5ab6cc-6f68-4b6b-95e2-954c6b57ba9c | 30274B4F-13D5-40AC-8929-A986860F38DB |
| e10568fe-0436-43f2-9f0f-48f9903868c4 | 30F623A3-DAE7-41DA-AB10-8786D094209E |
| E3953994-0299-4E04-9423-6153B142FB8E | 30FA311B-05A2-43E4-9EDC-02B327898132 |
| E4469259-2504-4987-88A4-02CAE33F46F2 | 3101785D-1CC2-4BD9-9FCE-02205D0548C6 |
| e488f613-f155-4e11-87d4-8c789ea81691 | 31397970-59D8-4EE8-83CF-901CD8BA17B3 |
| e6ec5a68-7555-4f26-bd7e-9cdb4c5f7004 | 31427699-E27A-4035-8925-6D8D6900D097 |
| E9F82800-647A-4D61-BD1F-2C17C4CCAFBA | 315CBBE8-62C8-488B-92C5-E2046321EFB7 |
| EA3A524B-6EE3-4904-8924-29FCAA20985C | 316625b8-a217-4829-90d1-dfd9cb7e4bc3 |
| EB9A558C-39EF-42C0-8DFC-64D01B729426 | 31e58fac-3ac3-4eeb-82d6-3f2b5cb8a60d |
| ec8268c7-943d-451a-9036-f0196078b6c5 | 32128246-1258-43d1-b1e5-30ae63822c32 |
| ef3ce0e5-d2e8-4616-8b1a-a78c591cf235 | 3223FEC6-1D35-419A-8EE9-7C5885B8BE1E |
| f1aa75c3-13f3-4bd9-9e51-923c2f62c2f7 | 3265F9C1-7242-4C7D-B4AD-B6519EE94F84 |
| F1F80DD2-891D-46DD-A3E0-F007628CB515 | 32AAC152-322B-4D7F-A78B-448FECDF17BF |
| F519BE07-434E-4EB2-ACF9-318565B83D50 | 33C0199A-BE0C-46D1-BB1E-889BFAEDF20B |
| f625e522-226b-450f-af94-dd2f5adb605e | 3424120F-248F-451A-948C-E320C19D1B19 |
| F6A81D81-7DF4-4273-8A86-E0CD3B8B1DF5 | 3428AA0A-127D-492A-BF4A-EE82563D3701 |
| f7287eb4-c77d-4c67-a768-87fcccd668f5 | 3434b91a-c05f-460f-a078-7b1bb6e7085d |
| F788F950-7953-47A6-B833-F9E5DAA28FD3 | 351239DB-52D3-4AB5-BD4F-8FEAA6701BA1 |
| f7a2b87f-c265-4cc5-a792-428756e31d8b | 35277C30-010C-4D3F-9DFE-754BF72799D7 |
| F94AE8F2-D555-4C2A-97B0-DBE7D642C566 | 35BD694D-1DD2-466F-AB27-03320614B40E |
| FB54458D-C373-46C2-841E-82663E13EFAA | 360d1d6d-cf3e-4c78-b36f-ef5e3160aa6b |
| FD15FF57-F121-40EA-A3B9-B66CBF34C1E0 | 36449392-de76-4ee9-bc09-627a07e18727 |
| FE57B639-DB7A-460F-ADFE-552F1E034E46 | 368689A0-F782-4826-9F08-CF0E0055162E |
| 0073A136-D5F4-4FD6-88F9-711768F2ABC6 | 36ABEAF9-CC0F-43DD-9C55-B9347D826529 |
| 01A13ABA-74A4-4895-A5AD-E5119925C202 | 3713865F-B6D5-44F4-809B-8B17B696ACE3 |
| 029b33b1-7ea9-4b77-a615-7eb0e3ebd5e7 | 37D13493-975C-432C-BD21-65F383FC66C9 |
| 032096A6-1923-44B9-A1CB-181BED041C8E | 387DB1DF-EBAA-41C2-B036-F46AD61E313A |

| | |
|---|---|
| 03ab1e9f-4918-404d-b178-3a4fd929c5e8 | 38a8b734-9acc-42f9-b5b7-e51b0dfc6504 |
| 048056CD-39DD-4134-A58B-70BEBB3DD2FF | 38f4b9f3-6709-40ff-9a81-5fa2111b1ce6 |
| 04E6E843-7F31-43DA-B111-280CE2BD1949 | 39303E7C-5BAF-479D-A112-D8F03A18C621 |
| 0536F465-1650-4411-823C-C660BC7F3ADC | 398D5C2E-F877-4BBF-A10C-486B8523C9CE |
| 0639B753-4099-4CA4-ADB9-763069A614EF | 398FB71B-CA83-44E7-BF0D-B1CA464B0283 |
| 0B283F95-DBD4-4C36-9201-2198A90EE0D6 | 39A5AEBE-ACAD-4141-AA5A-66228C4049F8 |
| 0BDE34DD-DAE9-4533-98EF-F5A61A11E7C4 | 39acb803-f282-422e-ac56-a3a50693d203 |
| 0CA72EBD-FF33-45B8-A97C-3F1435603D71 | 39D90499-8549-4DA5-B391-C421F18FACF1 |
| 0F8A76A1-4C3E-42E1-9DF3-95AA3AE8E676 | 39E24241-CB1F-445E-B167-82EE44BFC14E |
| 1043BC90-72C9-48C9-AE24-F617D0485E78 | 39E68907-3FFB-4CA1-A955-DB5FA2F0DEB1 |
| 10887E42-444D-49FC-AEE5-DD0E9FC4EA54 | 39FA91BD-DFFD-4B2C-AF7D-7FD914730B08 |
| 10fbe481-f111-4f88-ae45-4d67cb265567 | 3A0775A0-61AE-43F2-B5F6-629A44355C2F |
| 1133B8A9-6B11-4511-B70A-F200E3B8B5DB | 3A2FE6DA-5F3C-4AD8-B735-23ADFFE3FD6B |
| 11dffe71-b516-41e8-9987-e0ff5d2356d1 | 3A7C35E0-9ED1-4098-8585-52F5991B2534 |
| 12F90EFB-C45B-4372-8197-2DFD9CA247EA | 3A867DF4-D548-4408-A90B-441DED53B7B6 |
| 1610D467-E83F-490F-A971-7B099F3B46F8 | 3AC62513-B5F4-4063-AABD-DAD08A1F56FB |
| 1701F4AD-58C5-40D8-90C4-99E3ACAC0104 | 3AF5B391-E72F-463D-A086-A86C6C30A51A |
| 1BFF1E07-4923-43D5-B2D3-CB46B6568EED | 3B68B6FF-7EFB-4D53-8BD8-C76CD68AB7C0 |
| 1DB4962A-3CF7-4F36-BD60-46A1B63B6C20 | 3BD2FCEE-5B16-49F3-A717-F1E106BBC12F |
| 210726BD-DB19-48AE-9A3D-E6AC44480E85 | 3bf81a9f-abb4-4495-a514-fd3865e13a03 |
| 24E5D323-F3D4-4ADD-9A29-79E77CED357C | 3C2A6E30-A507-49F6-8B1F-36EB3AA41E60 |
| 284680B2-F961-402D-8740-E5F9E4FB4A98 | 3c612e12-6de8-44fa-a095-805c45474821 |
| 29E95C77-F55B-42E1-B81E-EAB4BB3CF803 | 3CA67CD0-81DA-4F68-A9FA-F1A8C013C28D |
| 30ABA886-5152-4E25-B36F-6CE7009D419E | 3D190F07-2F79-4EE6-9F63-0B563DC05C74 |
| 30BF5895-0671-4FB1-A96D-857847FCF7B0 | 3D2FF6C0-8C03-440D-8EC1-231862BD6713 |
| 30C7445E-D0C4-4A0F-96A4-27E4259EE10B | 3D644EAF-82FC-4BC7-93B7-3E600E8C6511 |
| 33615113-791B-4286-A23A-AD3D1F8D4B09 | 3D9828AE-EB65-4D0A-8D7A-CC09E98B5E99 |
| 36cf458b-3c8e-48e7-92ab-aea1c698e417 | 3DE1A86B-AAEB-4F0D-9F32-1FDA924198D7 |
| 38ECA720-AD68-43A7-8A1C-F79F838D5D03 | 3e290dc6-aba3-4135-b844-3e8c4cc8a25f |
| 395798C5-3136-4A30-8557-A67A3ABDECAF | 3E41CF2F-3DE9-44C1-A819-BCE693BAF67D |
| 396FF766-A383-4F42-BCE3-F2F32B7F1151 | 3E71FA1C-855C-475A-9DD0-A314A3ECD48D |
| 399c936e-ba68-4040-98d4-917a87dad603 | 3E9FDA23-676D-4E78-8948-0C9DF3D10064 |
| 3A161C85-2544-4283-A10D-E332C9DD1692 | 3ecb0147-276d-486c-9af0-ce437c24ba83 |
| 3AC41CC6-5DFB-40A1-BD13-5DC7CA2BB6F3 | 3EDB62C1-25AD-4DEB-A005-E881A887D06A |
| 3B19B5D1-13AF-4669-A082-1BD546048185 | 3F0A06D1-4C45-46B8-A06D-036C8998342C |
| 3da2e90e-6450-4286-8c74-08dbecbfd788 | 3FB3D96E-85EF-4071-A220-2382F8D9DE82 |
| 3FEA3F02-2DB3-44E0-984E-C61130D92C9B | 3FC062BC-1B0B-4C8F-B79C-770BCAA1EC6E |
| 40587B69-0F62-4A2A-88CB-1BD711377F19 | 3fe8f8e4-5396-4362-82f5-366e2fb69db2 |
| 41685C5A-A548-483A-8A20-305AD8D61771 | 3FF9B46B-325F-4655-AB40-953154753DAE |
| 41E4DC71-7767-44C2-9D38-110DCD37B451 | 402fd96c-76d3-4ac2-9f64-77bd34be0adb |
| 41FBB9F1-4A13-48AB-8649-0608C89CDD9F | 408cb583-6dc3-4698-8bd2-e284042bd5ef |
| 42156C6A-416E-4832-9C95-CEE6084B0910 | 40CB6F49-3D68-45DC-9BFA-B4D724DE7DFE |
| 427F4D23-2B76-4A2A-A498-0C3473F57169 | 424DE829-9EFD-4689-AC5D-24485A5D5ECF |
| 442DF188-B678-45BB-BD9D-7071ED8AAA5A | 42D342D9-F8A5-4940-9B6B-5773D66055F9 |
| 451BAD4F-6D72-4526-9978-38E090FC479F | 430AD51D-F68C-40B2-BFA2-9DA68370B7FA |
| 457EC7F6-8612-4641-AAEF-8A7F2C44C30C | 432FBB6C-331F-4D2D-834D-F6378D88107A |
| 4BD8EBC1-6F6E-44A2-8544-106901A7BAE5 | 433a32d0-93d1-4897-8e36-beb1ec1d998e |
| 500B86B2-3AC3-4745-AF84-3DD28F38DAEB | 43B3BF47-7313-4DFB-952A-EC0B9D48E16C |
| 505E9812-D13E-42D3-AEE2-55A85D6F4551 | 43CC3C60-F0A6-4B44-BD7B-8EC4DA71991F |
| 5469c0e8-0c58-4870-b897-8fa0704599a7 | 440421AD-5F1F-4345-86A2-FC25CB7CD453 |
| 55BAFAAD-323B-4B92-813E-52D18D70FC3E | 44465A32-8EE9-4575-94B8-0200BA7C5BD4 |
| 56353D9C-9578-4D62-9DCD-92D800EFDEA0 | 4480D290-5E8A-4289-8E3C-DE087E0DE412 |
| 58650D08-7685-41DE-9AE0-21665F8F449F | 4482DA24-BD16-4826-A9EA-1078BCB12411 |
| 58c82c76-9198-4c95-80f8-c043d1cea5eb | 44B5453A-3009-43B9-BC34-71A11A6D5E63 |
| 58de25d5-8a32-4412-866b-acac2e321bfa | 44d99f11-ef5b-45d7-b7df-b5c5893d878f |
| 5E2539E0-2088-43DF-AD2A-F854D8BB6D30 | 451E4916-80EE-45F8-AD70-84340868A5A9 |
| 5EEF573E-6A44-4DC7-8AA7-B136863CE910 | 459028fd-0cd8-4e6d-a668-71fd60bd0a37 |
| 5F6ED48B-3B5A-4D20-8FBF-C9AA3FEEA186 | 45aacbb4-0fd8-431c-a57b-279ea57a19fc |

| | |
|---|---|
| 610603D0-43FB-4A4E-B368-D143E6C61802 | 45C3975C-D333-4237-93F7-E12DC2451C38 |
| 669605F6-39A1-4F8C-81DC-39430FF5F892 | 4648225e-5b68-488a-969a-e8d812190c6f |
| 67E9ABC1-4B6F-4054-BDC4-29906C55C682 | 46801BE1-F035-421F-AAE4-81BAF3BB8D40 |
| 6a21c948-cd85-4150-8c01-83017d7dc1ed | 468061C2-0ACB-4F67-A357-99CCBD81A8B7 |
| 6A8241A8-D9EE-414B-89FD-4C3B9F181CC7 | 46E064CC-933B-444B-9E15-80FADBC53487 |
| 6D17C327-435A-4CBA-9626-3B5B7B7375BB | 46E51EB2-0C5E-457B-AF1A-8BAC1B8A8BEA |
| 6E9F74A4-2F41-4B2F-B0E1-39C4D779BD40 | 4729410A-7240-4ED0-B478-78CC6C7A4810 |
| 6EA380AD-D73D-41B2-B448-2D224DEEC01B | 4770EC81-9174-4D2A-AF54-70C00696FB0B |
| 6fab9c30-354b-4f99-9b60-6823c12b9b0a | 47F18EEE-3E56-4388-AAC3-9AC141BCB414 |
| 70C5CE85-4022-46C0-8BD0-F5E9009DDF41 | 48ABE0EF-DFD5-4E36-A319-C4EC3165CBA1 |
| 722C172F-46E6-47A1-82E5-3207278DF89B | 48D81B1E-CD75-4B55-A0B2-C331BE2A88AE |
| 75DBC8FB-4DB8-4764-824C-ECCF3A223884 | 497cc506-03fe-4ffa-860b-5a12adb1bc7d |
| 7638E294-2DCF-4C03-B238-24E40491C2AD | 49A2FD48-744D-4D88-B9B4-8C778D4F48FD |
| 765588AA-075F-477F-9C18-099D018D3A1B | 49A3C7C6-81B0-4B14-8E13-03F6901171F8 |
| 76CB16F9-37BE-450E-B60A-5F0D0F1EC2BC | 49EB77F3-F7D8-4E98-9F91-547543028877 |
| 78A212BC-A740-46BF-A10D-0B9809A5BB3F | 4A31C620-5C6E-44AE-B7F0-0553C4B7EB33 |
| 7cce924f-12a1-4895-9866-5d4b1869dde6 | 4ADC20D7-5AF1-4126-9073-C7D7AE465D92 |
| 820AEA32-8F1C-478B-AB56-8171425CD76B | 4b0d295c-e185-4b52-9752-178e5bc1d47d |
| 83a6ed20-b3cf-48b9-8ed3-a080cd4951fa | 4B185183-7E94-4AAB-B19D-6A2E48B85A5D |
| 84A5D5FC-7350-4846-A1BC-8BFADBCFF8DC | 4B3C152F-A9EA-48CC-9189-F09078A92C48 |
| 85C11B74-9E50-4DA1-8C0B-D5677CC801B1 | 4B50AEA4-4AD1-4BF6-9CF1-984C28A99C84 |
| 861F94CE-45FF-4588-ADDB-E1EA48878C2E | 4C197D76-DAF6-474D-B561-DED42905F52A |
| 86aa5790-9afd-4653-926a-625516df1f56 | 4C313E5B-4DC3-408E-8E92-22427F74743E |
| 872A12B4-8CBD-49D2-AC98-D846587F5794 | 4c42dc4e-66b7-40bf-9fbe-b92543248198 |
| 8996DC64-8C5E-4E07-8ED8-2EB11C03626B | 4C9FB311-0860-4A04-A2F6-79558AC1D98F |
| 8B20DD03-46DA-4D6B-95B4-F329FDC486CF | 4D1AEC67-C345-4721-8EE8-356B0DFF009B |
| 8FC9CC74-F388-49F0-B957-DEBB62638634 | 4D215C5D-6813-44A6-BA6E-03C24DA903B0 |
| 9084A558-11EA-448C-9F9D-0F90A94A29AF | 4D30606F-3AB6-4EF0-9991-8897CDB9960B |
| 931B549F-B9F2-4E8D-83ED-FF663671883C | 4DAEEFC3-0E2E-4537-8E13-AF9717B3CE29 |
| 94B95C07-5F70-4A1B-B40F-DC13F6A160D5 | 4E4E6280-713C-41BF-BF3A-E93C2FA44D59 |
| 977118C8-B446-4696-AA6E-9F1A1055E241 | 4EC2BB20-2190-4C37-8AAA-4086FEF1FBB6 |
| 97831d28-ab41-4c18-bfc2-c4c6bc757d13 | 4F321D41-3255-46A4-AE0D-1CF39699E624 |
| 97FFBE14-9070-4A5A-8760-225A88834B40 | 4F340C8C-5912-4257-B57D-C096DA254C26 |
| 98784BBE-E286-48DF-838D-1A88040658BF | 4F4E8834-A716-42E6-BD51-60D80A619202 |
| 98E82E98-341A-4BCA-92C0-23C65C1BF94F | 4f608829-ffc4-4527-886e-6bc764ab29f5 |
| 9922DE10-9280-44C0-B86F-6D1CA7E55BF2 | 4FC8E011-4433-4537-B03F-457A3A70240F |
| 99ac4f2e-2e63-4f1e-af0f-8f23a2caf999 | 50072717-861C-4B60-B595-D8EA23BB1C67 |
| 9F469EDB-6B12-4EAB-BD6F-9C10034D4CB7 | 50239d1d-b3a4-4240-ab83-b24ba67b1299 |
| a0339182-2fd6-459d-9703-422474222e76 | 503E6C30-1B62-4F85-BCB4-E34C009894AB |
| A1192DF3-7667-4D1E-A1A0-79F6CC827DCF | 50612721-2E12-4DC3-8E1E-87624CF9333B |
| A343FBE3-147A-4656-9E90-5408298CA12C | 50EC4170-29EA-4EE9-8CE7-67EC78D1DC76 |
| A418FE65-DFDD-4C84-B27C-B4567B50F457 | 511B5F28-709B-4E43-9EFD-B2F425BBA962 |
| A477B55D-A591-482F-A554-D8EF7298C23D | 515b1400-e896-4f2a-81bd-70895441a0bd |
| A56C2083-33D5-4C98-B85F-3B68A3D80ADD | 51ABE033-5FB4-49B5-89EF-C44D0BB1AB00 |
| A5BE4E42-CB6C-4E67-8547-45DBD058BB95 | 51FECA45-114E-4817-99D6-1C9842CD679A |
| A6E5F869-1182-4F81-B4BB-CF55261C8F3F | 523E24A2-51B9-4658-BE2F-42E5FCCEBB17 |
| a73c0842-39c5-4c2c-aa6a-f31341a4f36e | 52BC133B-8A10-4C9B-ADC3-22910A191E6B |
| a7911bdc-13f1-434a-bfa3-f8ba55d15846 | 53EAC147-7EB8-4C54-A982-5E058D6749F8 |
| A857D684-7195-44F9-BA21-B88020BD9633 | 53EB4C4E-0719-476F-9B2B-279D438D7710 |
| a873475c-b2c7-4c84-bd89-5b9ee6060ce1 | 5436FAFB-BC16-4450-AC8B-568E1E5EE5FE |
| AE4F0714-3B74-46C0-9C1B-F028C7CF2E62 | 5452FFD8-0408-4A78-B2FD-F1E0B0D05276 |
| AE61A71E-53A1-4884-A4A6-729744D1C309 | 545C9F09-DF14-4302-B131-519D141438D1 |
| B106D71C-7809-4EFA-A969-EE1F93A44720 | 54782BD7-79BE-4E3C-BF99-844E7BB0E5D7 |
| B297C4DC-EC58-4392-908A-586E81D57EF0 | 54B295D4-6315-4416-BDE7-A221859F965C |
| B2B57E93-2722-4546-9A0B-20155B86EDEF | 54E45B43-363A-47A3-B43B-5E41FF589B47 |
| B30F5B13-8CC9-48A0-93EA-3D49CE8A7D0D | 55DEAB98-29C3-45A1-BC13-BCE3F13AD481 |
| B56BDBDB-43AF-4A03-A072-54DD22D7550C | 55E03FFC-F006-4EC8-8E5D-A738FF091F8A |
| B838A4DB-EF0A-4D88-A220-BF57E31E0B3C | 562A562B-CA03-44AC-B282-B51B457D8DAE |

| | |
|---|---|
| b8df1577-2a75-4eb7-ab27-f2dd76160730 | 56403463-1727-4C23-A709-DE5F168C4073 |
| b9a3b647-61b3-4d98-90ca-e307ab50984a | 566792ae-f853-4a47-856d-f02cdcfcb18a |
| BA65B478-65D0-4C9A-A2C2-D32B29A1A9C4 | 567e4d90-5c5a-44f0-a462-90c831f3c096 |
| BB4A30D5-4EBA-45DB-B05A-BD843044D508 | 56FDF145-26C2-44D2-BF0E-67D8C8999A6E |
| bc3b4816-df1d-44ed-a8c6-f32c8e011a1f | 570F9947-1CE5-49C7-9619-49439DF6C7A9 |
| BC6FBB3C-870D-40B5-983B-7C88705E020D | 5779F7A7-05D7-4972-BD4F-AACB32420510 |
| BD5374C9-7F36-457F-B66B-7CC7FF64224E | 5798D741-C198-4C14-BCC2-E3B17CAF1BE3 |
| C2F9DFF7-751B-4704-8E6E-F4F800297030 | 58BDCACE-0047-456E-A225-68953EDAFE5E |
| c371e2b4-209f-40f3-a30c-51df8270b5de | 58E66976-4507-4552-AC53-83A49A142DDE |
| C51CCFF8-2502-40E1-B130-FE655C119169 | 58FF4366-20A3-40A2-A87A-551E17CDB5D3 |
| C56FA247-AD97-4DCE-A2D3-9BDFC5DC7B1B | 59A7D695-CB0C-47B8-BA9D-CA52CA2CFA7D |
| C6CEB220-282E-4EEF-8D36-4CE0A625CF7E | 59DA377A-12A6-454A-BE01-886EDA894179 |
| c81d4681-c02f-4b33-abe0-b6ad7d61194d | 59E47734-96DE-4328-ACA7-E9423DDA9C52 |
| CB1E9916-C03F-405A-A797-15823DF09C8E | 5A2D3BFA-4BDD-417D-9796-E3699760114C |
| CC8614C4-D535-4AD9-82F0-F4DC7AD11D0F | 5A53EC47-E63F-44A2-8078-87659544FB04 |
| CCA23650-1B06-4977-91E8-F35E73E70C48 | 5AC57AEE-4BE1-4A29-A53F-343F5A3D2E86 |
| ce3585d3-7388-4d05-84c8-64e0883003b3 | 5ACA16CA-4516-4E27-8249-6914029A7EBF |
| CFB61FD8-0BCA-40B4-878C-CC914A289792 | 5AEAC31A-176A-4F93-A376-A93A670821BB |
| D209CFFC-FE21-4AE7-B610-E5668EA84A3B | 5b7e557a-6aee-4077-b3cd-c96179e643ae |
| D344DAE0-ADF4-4697-B703-7690C13D8331 | 5b9d7565-a933-449d-8ffa-ec5f82db2e77 |
| d39df936-4bc6-404d-9e1d-f6416cc5a34c | 5c087d33-0bd3-407d-8350-fabba6aec597 |
| D3A6ACCC-5C45-4E70-BAF5-2188AF53E0DB | 5C118E15-1E48-42A6-8D29-179E6E71A26C |
| D3D8987D-0A54-45F8-B9C9-17EC89FA864E | 5C68E3E4-58F7-4699-989D-80D2E4996794 |
| d766c3af-f0b3-4791-a754-d6546d370f8f | 5CD79093-1571-4F71-8136-0D84CCABDCAC |
| D8628D35-C4C6-443F-9DA2-8517A157F137 | 5CEA1082-2AD5-4233-A6E3-067566780471 |
| dcd45077-f068-490b-bdcc-4d4a62285116 | 5d1d00c6-fcae-479e-ae1e-de76efd41d98 |
| DCE168B1-D2A9-4409-8437-AB382F6D7D31 | 5D5DEFB3-8572-467D-B383-7358F802B398 |
| DCE21359-004F-4E8B-89E4-9E1A30FAFFDF | 5d688049-f271-40ef-813a-d37240ffe21c |
| DD163547-43AD-4FE4-873E-83C4C6BE4465 | 5de6fd84-ed02-4c27-9947-db57f05faf6d |
| debe24c5-e4df-4e80-82f6-8f40395aa9e6 | 5E972E96-7271-439A-905C-6E308C1EF29E |
| DF22BC30-96E2-42DE-9A39-2954DB49841D | 5F263B2C-3385-4D07-99C2-D7B613C4A470 |
| E1758D75-B2A0-4386-BD72-6B2421D4DA6B | 5f2f2e7a-8aec-4011-b261-0c5bc68da840 |
| E349D8AA-BB82-4CE8-B14C-61216289F25B | 5F853A88-811D-4EB4-914E-D90862C5D2A2 |
| e3ed0c19-6acc-4be1-aae2-73fe73cc9f24 | 5fce954c-11b9-41f4-9471-4b5bdc0efa7f |
| e4d9b696-3311-44e7-9191-fde56c080a00 | 5FD37868-4762-4109-9DCF-6FDBAB5B645D |
| e669a4b7-a494-490a-b78c-e664c59d0697 | 600D5316-B795-4AB2-86FC-52835059386F |
| EA31C705-4CF8-48F5-8D13-E43563213C91 | 60117B70-36C0-41BB-A9FC-85DB8FA10102 |
| EA8DBC7A-54C6-469C-865E-F49D00B0223D | 601a4146-8093-4f20-9a0a-a97916c9e7f6 |
| EF12BFB7-4147-407F-84E9-586DBB10D583 | 60CFA29F-4770-4EE4-AA37-787F53155662 |
| F005FA99-7E4E-4308-B11C-83B25683B1FD | 61FF31A4-4FDC-476C-B456-2A2FED4BAE6B |
| f3c73dc9-6280-4619-a629-b575fdbdb0ca | 620e0648-ec20-4a12-a6cb-5546fe829c77 |
| F4AEBD64-A356-4C05-B847-6DD28CF26339 | 626b4ff0-2188-4a14-aac8-6703c624917f |
| F79420CA-B0AE-417C-95F7-D28C35760202 | 62ca707c-0657-4b7c-863c-99f4ec475972 |
| F842F756-CE78-4C99-BCF8-3BCE18A4D9EF | 630C683D-D1A2-4BEC-AA20-38EF59227D97 |
| fb3884df-3680-4c5b-8092-981007aeba03 | 6385194A-D75B-4BD3-9E70-9AA36250D5B9 |
| FB6437F9-ADE8-4C33-A0D0-CEACB2BF784D | 6397D7BC-9CB1-4A84-81AD-76954E0A8CEE |
| fc8828ae-484d-4fc8-a168-f8bc58ad65b9 | 63D646E2-60AA-4BD5-B2ED-6254C9C51BE4 |
| FEBE2CE5-737B-43B8-BC70-4194FE3ED5FB | 63DCFA32-C69E-45AD-BEA3-AE672ACB11C6 |
| | 63E8C48E-F01B-4A7F-87E2-30607A2C8F6C |
| | 63F4AA26-DEC0-4309-8B75-6446A7241527 |
| | 63FA8434-4A6F-4C32-9C5D-89DCC913900B |
| | 64131B0B-9F36-44BE-8BEC-92C71EC36343 |
| | 6429C443-8AC3-407F-BB9C-66420B904BBF |
| | 6435EA21-7214-40C5-9CCB-A1F22685A728 |
| | 6454FF0C-26F8-4CC2-B77C-7E5ABDC20ACD |
| | 646910EE-5F5D-40AC-A21A-75790F89430A |
| | 6491D025-C061-4180-BFD4-D7C6E6E55F66 |
| | 6524A6D3-A9F0-4A46-AC0B-7BEB0CD1B915 |

| | |
|---|---|
| | 65ba24fb-07df-47ce-91c4-677684b64cf4 |
| | 65CAC997-4D39-4501-85EC-4FCB328A8EB5 |
| | 66229DD6-C771-4455-A90C-C66FB40CF7FE |
| | 666D8B0C-593F-4EC3-B16A-2DADBEAD8C32 |
| | 66763a0c-6cda-4832-a0cc-e7b496d78eaa |
| | 66AF140D-DEAB-440F-BB58-B8C3DA2C3CA5 |
| | 66af34ea-f471-4092-aff6-9ac9a5b75b11 |
| | 674E010C-7AD4-4DC2-86B0-284CD4560633 |
| | 67560840-70FB-45FD-8572-648F74811AEC |
| | 677C9D22-E518-43AB-9668-A606C35A6EEB |
| | 67C5F371-3FA9-47C5-8B15-C2DD9ACC8519 |
| | 67C73260-A242-4BBA-87C5-D2302556DFF7 |
| | 67E92755-396E-4D9A-B737-979EADC743F9 |
| | 67eae8b3-3bd2-4e87-af78-ffe8221f318d |
| | 67f2ac9b-b9fb-4ffd-ae60-6ff1114e9290 |
| | 6817BB3E-1F8A-457C-9AB0-31CCDA8B6868 |
| | 6868BA1D-B612-454C-8146-74AF2F573D76 |
| | 68F612B7-33B4-4CA4-B012-439BFD598888 |
| | 6949C03E-067A-49A5-B48D-E9380A1B0B27 |
| | 69BABA9F-19C2-4947-95DC-0DD3412929BA |
| | 69FC24FC-CFE2-487A-935F-1A954B30B709 |
| | 69FC74BC-E961-40E2-9169-D7B51658EB10 |
| | 6A0490EA-D9C6-41CF-BEC3-3257E98CC6ED |
| | 6A82A217-F7A2-4697-98A6-6C6553E8FAC1 |
| | 6ABC861A-376D-446B-ACEE-FB0F03A82C09 |
| | 6ad260ad-769a-4f3c-bd07-927adf6b2338 |
| | 6AF6D256-A753-46D2-B611-E4094FF265AA |
| | 6B529FB3-0B1B-4D3B-9309-BFCB063DBB71 |
| | 6B85D76D-6098-47EA-9823-586238526CF1 |
| | 6ba76174-620e-4851-857a-d0adbfd6f3e6 |
| | 6BCF8B41-7ABC-4B6F-899A-1F67B4ADA765 |
| | 6c2a0f37-49cd-4fd0-b126-58b35f9121df |
| | 6cdc0d53-f813-4101-81e5-9bee68270536 |
| | 6D01421E-D533-4967-8422-97FBC0FB0895 |
| | 6E126B73-D3E8-4641-A128-306F3B313E40 |
| | 6E1C9AC8-EEAD-4DB9-8F30-EF3591DAF78B |
| | 6E6B7742-A562-490B-BB72-04A5653852E4 |
| | 6EC931E1-7A6F-47A8-8B30-F37271A50881 |
| | 6ecccbf9-bfda-4f5e-9618-3bffaebf4660 |
| | 6F02648B-0C48-46EA-874A-407F951F0409 |
| | 6F0FD68F-ED6A-4C9E-BE02-C97BA4950F76 |
| | 6F18F96E-C0C9-4CC4-9FB2-CBA8949D5402 |
| | 6F4C8D30-47FB-47DF-9EB7-4E5881E3711E |
| | 6F77FC81-1BF4-47C8-B1D8-78BB17CB13F8 |
| | 6FA2A667-9C36-4526-8A58-1975E863A806 |
| | 6FF12A54-10DA-4941-BFEA-7B66E19B4BE9 |
| | 6FF5C8BD-9929-43EE-B53C-3C7C0B23F97B |
| | 70081320-540F-41D2-8687-EE6D011F8EB0 |
| | 705F9841-988A-488A-8CDD-3E00BC731E76 |
| | 7067B7DA-FB1B-4827-8FE6-9433ED672D3B |
| | 707312FE-1D92-413E-A891-1EA3F496C343 |
| | 70c2c861-01de-47bc-abfd-b7d14d68e1a1 |
| | 70F1569E-F8F5-4F1D-BF18-D0586B29077D |
| | 714B4003-959A-47D5-8912-A250B90D7CD4 |
| | 719082cc-1ebe-4a51-a659-85a59db1d77d |
| | 719FA3F6-0194-423D-8A96-3709E8180565 |
| | 71EC4C84-4AEF-4C1F-AB07-DC264C5C0602 |
| | 726C6892-0DCE-4869-A7A7-CC44C26FE843 |

| | |
|---|---|
| | 72e486b8-a866-4916-b2e4-8b4bb5dcd92d |
| | 73AA428A-4288-45F8-8A53-CEC173DCC61F |
| | 73B7D4BC-6B56-4AE8-BE7F-074745563BEE |
| | 744DF1F5-A7D5-4675-92CE-6E4713F0D541 |
| | 74543fa4-ce73-46e4-9c59-224e8242b4a2 |
| | 747083ff-0703-431b-aad4-f2adff739516 |
| | 75113445-d2d6-44a0-866c-c9175e6d214b |
| | 753D5D49-3B40-4737-9D4C-B1EC29CC2692 |
| | 7555E736-A9E2-4B96-A633-FB0F7224D62C |
| | 75783DBE-E18C-4535-B9EB-2AFFD320E098 |
| | 75A43851-8FA0-4259-B843-88DC6FBCE270 |
| | 75BC22D8-738D-45F8-B6C0-C664F5FEF47C |
| | 765AD5A5-134B-4680-A6D4-1DD113DC47CC |
| | 76618CF5-F6E0-46EF-A1C0-ACC0AD9FDCCB |
| | 76ace6aa-0505-442c-978a-f889cf2d44bb |
| | 76B0106C-A4D2-4B29-BEF4-FE03A25B5D21 |
| | 76BD9BB9-9A8B-4A90-BB7C-EAFE76472EE4 |
| | 76C4D298-6F17-4F11-A706-CD844A0E0914 |
| | 76F98002-E8DF-4DFA-926E-8BB2D5114966 |
| | 77396AE6-3838-47BF-AAAE-F9A0F401EC7F |
| | 778A8F53-53F6-4A03-8CE1-B0928539A444 |
| | 77BB90F0-E923-4E28-BB88-4D6B420E0B5C |
| | 77ED36AD-222F-4693-8668-0ECF9073D6E8 |
| | 785DDEA1-399D-47D8-B2B0-39E95EAA24B5 |
| | 7861a5f2-8910-4bcb-9c34-f79c5acd6e21 |
| | 7861DCA2-BADC-4575-A388-621D3614A8E4 |
| | 78d14f86-896a-4f98-9274-b2e8e387add9 |
| | 78e1da41-127c-4e9c-aaaa-77a0d94c31d0 |
| | 78fcc813-d0a8-44aa-b004-62810dca9d12 |
| | 790ff5db-b7f3-4946-aaed-305f66b1dd6a |
| | 79126E88-9CF8-45FF-BF91-A98DA3B304D7 |
| | 791c5768-f0f5-4ab6-86eb-998e5c4b49e3 |
| | 791D3824-890F-43F2-B545-C7266506D716 |
| | 7A32847F-69BB-4592-8FEB-CFD5D355ADA9 |
| | 7a589441-11ef-4158-87e7-3951d86bc2aa |
| | 7A8D2E13-315E-4512-85CE-B018D11C3BD3 |
| | 7A99A808-5AC7-49B3-87F6-106663EB323F |
| | 7AA71C68-4205-4745-838B-0D00BD49A501 |
| | 7acf33e7-c386-45fd-8c9a-469600378687 |
| | 7aedb46a-48ec-4e0e-8baa-c9ea4c4a7cf1 |
| | 7AEF4DCF-0A4B-4848-BEAB-377394456BEE |
| | 7B228C3B-E604-41A1-9B76-8DD3D5A59FFE |
| | 7b605fb6-d077-401e-b870-b87ddf505f82 |
| | 7B82E2D2-47F9-41A1-9ECA-40904C92039C |
| | 7B8C04D7-824D-493D-8F2C-9ED87A907192 |
| | 7C4AB1CA-4BDD-4EF7-9A42-F6C0C7136D26 |
| | 7CABCC66-F159-42DC-BD79-953BFFD93B8D |
| | 7CB42865-AF55-487D-B604-FAACE62BBA51 |
| | 7CD5ACE2-D340-487E-BEB2-1B576F6D751D |
| | 7CEC93DA-42C9-495D-8110-9FB43763041B |
| | 7D22FF96-9249-46B3-9CFA-E40DC4ECEBD6 |
| | 7deedeba-f05b-453c-9df1-eeb20b0ec72e |
| | 7E17E6FD-ED83-471F-91E3-5934885C8E87 |
| | 7E41BABA-7EF6-4030-B595-D95CD96336B5 |
| | 7e4e5633-6f83-4b46-b6eb-64b3f2866ba3 |
| | 7E9C8D3C-790C-46AD-8F31-B557B7C26F69 |
| | 7EB5D055-E1C8-4B93-9205-A49A6E79DFD4 |
| | 7EEA2B6E-771F-44C0-9350-38F45C8DBE87 |

| | |
|---|---|
| | 7EF22138-AC76-4846-B3BB-DDF1EFB96531 |
| | 7EFC26B2-EFF9-4F1E-9F4B-F26AF072C84A |
| | 7f0bbba1-9bc3-45db-bdda-eb3319bb15d2 |
| | 7F9FAD75-AC3E-4580-B949-80EED07A3230 |
| | 7FB87427-9B19-4378-86D5-F4F775F97805 |
| | 7fee9d6f-c0ea-43c2-89c2-66d965a077a8 |
| | 800D3A17-269F-4D97-94FF-8585F20038D8 |
| | 800F3039-97E0-441E-8292-4CCF1BB4797A |
| | 8027ADC9-4918-45F0-A7DA-5767DA0B999E |
| | 80420C04-709B-4AB2-A88C-1E22B3A09B9C |
| | 80593c77-4530-413b-bb05-adca5d43bb82 |
| | 8062CA27-542F-4A00-A047-C88B9B26F34F |
| | 80D51546-C046-4AF4-B3B3-B6BF5B1EAB02 |
| | 811F3FEB-A5A9-48B1-ACDA-FBB23FCBCF16 |
| | 81a7f6a2-e44c-43d6-b687-01158eaf44ee |
| | 81B70C58-4A12-448C-A594-2ADE44F6A0AE |
| | 81FDC97F-8BAD-4286-9D36-9A887C6B59A1 |
| | 829b41b1-693b-4f7f-a5c2-68c14b079aae |
| | 82C47F60-B863-45F9-82DC-72D89753F4B1 |
| | 82E85AC9-8E3E-4C2E-AE0B-A32CEEB8115D |
| | 83488A9C-6B28-4A48-850A-2A2FA47CE25E |
| | 83762308-4DBA-4870-B90B-64331EBC9BAA |
| | 83B97927-A1BE-441A-97A1-4363E1F2D530 |
| | 843C8F16-5E59-40D4-9ECC-8A75375156F6 |
| | 8488BB97-BF11-45CC-B1BD-52969598448D |
| | 848a5838-8a06-45a9-a3d6-e4ed47b824ab |
| | 84D80BCF-DA24-4B93-BFE3-3D2D53B8B07D |
| | 85AE3B0F-3A54-4A23-BE25-9A4DEEBBC011 |
| | 85D1AFDF-6164-49E1-B599-71512C5ACD99 |
| | 85DEF63A-39AB-4C83-AB8C-02C6F702F05D |
| | 8631969d-4ac6-4fe1-9db3-d47db604494a |
| | 8631BB5E-6D08-4C20-A85E-CB84F5DAC290 |
| | 86657A3C-10CF-48B7-9FDA-F2DF5E2AFAD5 |
| | 86731af9-2d33-4521-9cf9-5f6152ca15d7 |
| | 86D5865E-6A1C-467C-97C9-47460B77F5A4 |
| | 872244E2-D1ED-4A1D-9162-365F45E7F1A0 |
| | 8751429B-4A11-451E-B978-DC9E9DB0EB36 |
| | 875333ab-9048-462d-aaa2-693ad127e3cc |
| | 87858089-1BAD-4E5D-80ED-04F846DA9CA4 |
| | 878f975b-94fd-4d69-b7e7-1ed3ac2ee438 |
| | 8839F236-BC04-416F-84D4-50F67FB338DC |
| | 88C0AF76-B1BA-478F-A42B-8A50C0568D5E |
| | 89414e79-7f89-42d9-8228-101b5bf2f31f |
| | 8950691C-600F-4A90-ADF6-42A8F129AA3E |
| | 896E37FA-3A6A-481B-B790-5EE62BBDC9FA |
| | 896F5DCC-D076-4E1F-AE7C-52D21109DA7B |
| | 8980FB06-E38D-459F-A777-52048D492DEF |
| | 89a8da07-4487-47c6-8d3c-442a87e4f632 |
| | 89AAB5C2-9761-43BB-9053-EED42EE08969 |
| | 89C128B9-1C6B-4C04-A4AF-7066772E783C |
| | 89C72F0E-E724-4826-BB9C-D7524D1B1258 |
| | 89EA0E21-C5AF-42B4-9F64-5BA4874AB76E |
| | 89F4A6A5-BF9B-4A0E-BACF-8CD715C2CA93 |
| | 8A67ED86-156F-4B45-B460-000FE33DA26B |
| | 8A8E05AA-F13A-4142-A335-7AAFBEE7B664 |
| | 8ad1c98b-5395-477e-87a8-19dff3a59c26 |
| | 8B7C42CD-FC2A-4023-843E-A411821F94FF |
| | 8BA2CF7B-E474-4ABA-8C6D-9FD36F5C6759 |

| | |
|---|---|
| | 8C113BC1-101B-405F-9EB1-8C9E8EBBE185 |
| | 8C279BC5-117E-440B-B118-AF01FC6C11A2 |
| | 8C4F5492-3567-4E12-8607-2CD65FAB9BAA |
| | 8c5aef43-341e-4a37-84eb-fd730eb957c7 |
| | 8CC1E08D-75B6-48DA-90DF-E6AF23CB4C35 |
| | 8cf8b620-7ab6-4b6e-84bc-ff5a83f381fa |
| | 8d20352a-6180-40d0-adc0-c87fef854dfa |
| | 8DD09D74-0BE8-4C58-BF44-DE4A2466D044 |
| | 8E54232D-D8BB-4612-8B0E-4FEB076A60F9 |
| | 8E7792B3-2B6C-47AC-ACBE-3DA7462A8321 |
| | 8E9C3621-BC43-40D0-BB2D-0C8B7ADE37C2 |
| | 8ED09A8A-3284-4A1B-B369-E25D2FAB0F61 |
| | 8ED15B85-5BB3-4317-A93D-67EE376A8244 |
| | 8F10E006-531A-49AC-B51B-DF3FDCB20929 |
| | 8F1B904E-39A7-4870-B0F4-EFA1D43E50A8 |
| | 8F9AD301-6699-4078-A80D-886E40894AF7 |
| | 8FAAD47D-3686-44AB-A0E3-842F64179229 |
| | 8fc1f1be-d2d5-4b3a-9973-f4d964018beb |
| | 90507BAA-B716-4BCF-A18E-9D8AE97D809A |
| | 905C998D-4219-46EF-9906-8E92D7416794 |
| | 90750006-FB25-4CCD-90DF-9F300CD26A5B |
| | 907b64d6-a26a-4092-a247-ac9af444d1c7 |
| | 9084b672-8e65-4b7f-a256-32fd35aa8ab5 |
| | 9111759F-A0B7-452D-A0DE-838FE010245E |
| | 914DF75D-25F5-4B2F-96C7-95861D7DFA30 |
| | 9180E195-DAE9-4AF9-90CF-71778BC9AEFC |
| | 91B09291-5944-4691-A0D8-7D1CCC649B9B |
| | 91C94A0D-F088-40E6-A4B1-D2A72AD35077 |
| | 91D44093-D8D9-442C-92ED-E659C45B4E0A |
| | 91F67F4E-B8B1-4455-83A6-4D96D04E8CA9 |
| | 926176C2-0A8F-414A-A7B1-91734BDCE6AE |
| | 929ACC67-B8BF-4F65-9110-380E44AB66B7 |
| | 931A50FF-3149-4C4A-9456-45ED3A9DD457 |
| | 9348C35A-58BA-4DB5-BC4A-E9FC8E07CDF3 |
| | 939ABCA1-8BD0-4271-9A9B-666764EEB3B6 |
| | 94443DF0-7F16-4178-9E66-BEE82F6CA33A |
| | 953359DA-E535-48E6-97DC-1B4CCAD6A671 |
| | 95858EA9-F71B-48D4-AA15-7DFDF21CA9E7 |
| | 95873E61-AFDB-496C-9F77-3F9BEB008CDA |
| | 959E1F13-360E-4E12-ADD5-BF9A376990D2 |
| | 96B95A2C-FB4F-4CBA-89E7-A7F0E90DB146 |
| | 96b9c7db-1be1-4b60-b47c-26b654c3d64c |
| | 97384352-0E80-42C2-8A8B-7BDBDA2B58EE |
| | 97677FD7-8523-48C5-A8CE-614FD4010551 |
| | 976D067B-9D7F-4920-A04D-E831EDC6C2C2 |
| | 97C3C5DC-5540-4713-805E-4D98FD8C03AD |
| | 97CA0157-0096-4B0C-A3E7-BAB59078D2D6 |
| | 97CF3268-9756-4659-9E98-F200F96A91F4 |
| | 981066EC-ECA6-4621-8979-E935279AA1CA |
| | 989E9202-C08E-4BBF-85B3-1E278A475CB7 |
| | 98A08015-9040-4E0B-BCFA-2C6DB7D0F888 |
| | 98E709E7-E195-4B37-9537-F6081AFFB609 |
| | 9924E309-5C47-42FE-B937-547F2B3D6842 |
| | 9944f46f-9fa8-4e2c-a806-776bec7f8803 |
| | 998C8517-89C1-43B2-84E4-B60905787BF6 |
| | 99e32c59-aa73-43fa-88c9-399ddadb2c72 |
| | 9AA61AB5-F675-43DD-8A99-077164B41F30 |
| | 9B6ACC25-447D-4144-B4C0-EF52B8850D96 |

| | |
|---|---|
| | 9B781072-66D8-49F2-80F7-7B020746B6E9 |
| | 9BA72A77-AEF0-4788-8072-60C28902E55D |
| | 9BCE44A2-DCEE-4D1A-A522-C2BCB9A8E025 |
| | 9C516D28-BEF2-4334-BDDA-97950BD95AC0 |
| | 9C58E3B2-1DEE-4375-8239-CE69A725E3DE |
| | 9C6C44B3-A2CA-456E-A8D1-D375309C1751 |
| | 9C9B53E5-CDBE-4510-A839-4A3B5039681B |
| | 9ce7ce09-4409-4fa3-9f78-b2b24716f860 |
| | 9da462b0-93c2-4305-89f6-7199a30399a7 |
| | 9DBA5A81-3587-4F5A-8D19-E0B0AED0A677 |
| | 9dd8b7cc-d094-474b-9573-cbff4ab233c8 |
| | 9DDC014D-9A8B-407B-845F-452A73F08E5E |
| | 9DFDDC49-1E66-4E26-9D71-40D748CCA0A6 |
| | 9E2E29E8-D94A-499F-A98A-5B53B2C38F3B |
| | 9E5B6BEE-0E10-4559-8D0E-2DC9677CC90C |
| | 9E79059F-3453-4AA6-9849-55A267C711A4 |
| | 9EBDA267-A475-408A-A512-CD3761DE83B4 |
| | 9ed5c6fe-24d2-4ee6-88ba-7319c9cfa05e |
| | 9EE944EF-4293-4228-B750-CC34AB388EB3 |
| | 9EFD27C7-9893-49FD-886E-E85057DD5D2E |
| | 9F15BF13-11D2-400F-B6F9-C8BE0A57C763 |
| | 9F31DEB9-B9F4-439B-AEA4-F87C71406C79 |
| | 9f4574ec-a58a-4410-8735-c5e5bc4ca7e0 |
| | 9f80649b-a249-4fb5-97ad-059a30e13fea |
| | 9F92080F-F6E0-42DB-A02B-B3069664F6D0 |
| | 9FCBEF9E-2CCA-4033-B574-DAAB2A42198B |
| | A09D0BE0-FD41-4B30-B488-9A9F2ABEF8E7 |
| | A0BE056D-792D-40A0-97C6-FBD4976820B3 |
| | A0D35A25-3500-46F3-874F-1A0820200473 |
| | A1206473-CF9D-4BCA-97C9-67F35817B806 |
| | A1A233EA-5DDA-4EEB-BD92-6667E8B3EF57 |
| | a20fa2c4-a502-47e3-bfaa-2ec4cce3a859 |
| | a2b6246d-2b80-49c5-968b-694b4e8171dc |
| | A2D42990-747C-4618-88DD-E2DECE8E566F |
| | A2E2FAC3-CCA5-4E33-AA6C-1D9D3AC3AEEC |
| | A31554AF-D77A-4862-8AED-4581FB2BD877 |
| | a32cb96a-78bf-456e-a6ab-2d47b2c67ad4 |
| | A3B49C90-FA9F-4189-A442-64EC1AD97A01 |
| | A3BE4E6F-C845-4D64-8111-D63FDF8CC8F1 |
| | A3F98E70-7050-46CF-A679-CA7A83F4442F |
| | a40bb4bc-9ee6-4a8b-8a33-3a4ed78f69a7 |
| | A412B374-4D25-491F-B76F-DDAA989ACF59 |
| | A45833E1-7CC9-4273-B408-620F74679A12 |
| | A45D296E-EFC0-479E-B2F6-BAD834668CDF |
| | A4A217C1-66E8-4CC5-805A-1172CE5DA721 |
| | A4E2E507-7DDF-43DF-B959-FE451971AF1E |
| | A563109D-0990-4BDC-8BD7-6766D1EADBDC |
| | A5885835-9473-455A-BAC0-6FA43FA8A866 |
| | A633D316-59A4-45AE-8BF8-135CFBD27656 |
| | A6502F17-6BEE-4D5A-8520-06364593D39C |
| | A6635058-4498-463C-9BBC-1009DDA3308C |
| | A7030D2B-2CC0-46BE-9721-DC210954BEE0 |
| | A71B425B-CCD3-42F0-8163-3721CB1794EF |
| | A76774FE-7298-4D68-A2B2-C1BAD93F0C31 |
| | A82D0A57-4383-473D-B334-D13B278404B1 |
| | A88D6F62-88DA-40C4-BC16-7FE58F038B8C |
| | A8A33BA9-0976-4E19-B0E9-FA5BA4D4EAA8 |
| | a8a391ba-e595-4a91-bd55-02b312e55427 |

| | |
|---|---|
| | A91C3F09-5F48-4B54-B669-C768F9FE9682 |
| | a91f7a8c-e0fe-403a-9ef0-f7679b2761cc |
| | A947A945-4721-45CC-BC45-13B8EA41C10E |
| | A9C10A1D-EF0B-4A4E-8433-522A9E04796E |
| | aabc43bb-485a-49bc-b48a-d4e4ecd46d33 |
| | AAD9A83A-AB62-47B0-990C-527CCDB58D63 |
| | AB68CD1B-5075-4B30-A829-D6D0F95871CB |
| | ABBCC1D8-AB74-4459-BF28-CC627BEF440E |
| | abd6647b-d2eb-404d-8283-443e92addf55 |
| | AC2EEB9D-B5EB-465C-8271-C89BDB1BA136 |
| | AC359B60-1977-4AA9-99D3-8D17D7D66E11 |
| | AC9C3D6A-2DF7-472A-B9D7-0A5583463750 |
| | acb3b9db-16e9-40f5-87a9-9df9d4ceee54 |
| | ACBA8D76-999A-4F6D-B3A4-8D7DD0F2BB52 |
| | ACF5CBF3-09CA-4F4D-A44E-1792613190FC |
| | ACFBA23A-06EB-4FF1-BE60-0452A13A4978 |
| | ad1c5951-ca93-4102-889b-e28e2bcfea76 |
| | AD29744C-A09D-463A-84B5-63CFEB7FF786 |
| | AD80A042-C4B4-445C-9678-1C2614422684 |
| | ADB7C5C8-4AFD-40DC-89F1-571FBD88E4F9 |
| | ADC60720-AC20-4E73-9C37-3AD3119C07D6 |
| | AE10B73D-E7F4-4334-8452-167B60D71CB5 |
| | AE90972D-5BC2-4B53-B5FF-1B8C31F39342 |
| | AECD6689-F203-4519-AFAB-3F46EE9D3E2C |
| | AF0ACAA9-A61C-4E57-BC0D-453AD5BF193A |
| | AF363CEC-BEC9-4DE3-8CC2-E98B0EED2858 |
| | AF546062-0960-41F0-9ED1-6ACECBAD99F3 |
| | AFA7D9A2-3998-4182-BF63-54895ED2DE37 |
| | AFB00CD4-E5DA-4251-86FB-532C3F65324C |
| | AFB5442E-0249-475B-8F82-CAE4E0268AD1 |
| | AFC984DC-588C-4277-88AA-5D82DD8BD40C |
| | B05AEFC4-2D61-44B6-9451-E660D10D8A79 |
| | B0669EEC-6A1A-4618-AE23-AE0F9E9C266C |
| | B0700958-5F90-4546-B35F-635CD506889B |
| | B0B293C1-BD7E-4BA8-9F44-47398488EF2C |
| | B0CB81AD-3C20-4D56-AB7D-F64C0CAEE1CE |
| | B0DF5500-D608-416E-82F4-9A99206D24C8 |
| | B10A9314-FD40-459E-97EA-AFFC636DCEEA |
| | B12DEB95-6AA1-49CB-A3C5-A711D42D6FB9 |
| | B16469AF-A721-43E9-A4D7-C39B94D1139F |
| | B1B3983D-37D2-4BEF-BD17-708E3E600146 |
| | B1F27FA2-133C-42C0-B69E-815AF67E2A7E |
| | B304302A-F2B9-4CD7-AB14-C21EDF7778EA |
| | B305B716-5AE5-421B-B3C9-0824D37B08A8 |
| | B35FBE19-0E6D-427E-B9BE-EE3527B5FCA6 |
| | b36b7567-30d3-4d0c-84c4-b203986e0fdb |
| | B3720405-CE3F-49C7-A5D9-BBFFD06F6AEF |
| | B37ABDC8-B9E3-4F1A-B16D-2E59442DB335 |
| | B39C64BB-122D-44B3-9672-D0DC8468CA93 |
| | B39D2880-6964-4C5B-B5A9-AB64E5CB31E8 |
| | B39F3099-62B0-41C1-85A2-61989CEE0B60 |
| | B3DBF9AD-9602-4E44-BEAC-1CFA5F8BBA5D |
| | B3E248B5-5D40-495C-BD10-90ECF0ED6A95 |
| | B41FAB9F-AF83-4EA2-89FB-A5999B168412 |
| | b43a4f92-7a13-4558-a97e-7b3feb2d70c9 |
| | b43b8aed-d166-4f00-8144-24ad3fe314ed |
| | B46D88E9-8477-4900-8DA9-5E7DBDED0653 |
| | B47FB978-2DFB-4ABC-A5AF-6E14DD684182 |

| | |
|---|---|
| | B4A60A32-2C1B-44FB-92F4-B30F985DC424 |
| | B555B7A4-6324-4D3E-9EED-B140AF6C768A |
| | B597023C-FF84-44CC-BFCC-8BEE64CA4CFF |
| | B61E39F5-F0DD-485E-8E22-D4FDEA751F55 |
| | b6633c36-7ce4-4b69-9bf6-30b64d46c66f |
| | B6750B08-C59D-49B9-8C08-A19E17CB366F |
| | b6abbdcc-f826-499d-b6f1-f6e2c83f9d8b |
| | B6D1490A-AE85-4E99-A053-1614B498A081 |
| | B782C157-6B33-4247-83DF-A82DC5E5D432 |
| | B79558C5-1DB2-4B1E-A7B8-EFBD19AA855E |
| | B7BAFE7F-295C-4598-9CC8-49D785BD2218 |
| | B7C8CA4C-8D9C-4C5B-99EE-2FCEBE4F70E6 |
| | B7F2958F-256E-4C30-9C61-11E76DDB36BA |
| | B815FAE9-C595-4C76-ACC6-6D38B9AF9578 |
| | B82E08E3-AE79-404A-A501-0D501DF84B8B |
| | B835289F-6174-4BF9-AEDF-9EBB9EF4EB62 |
| | B84B58C7-95B8-4162-8E61-414F8FE422C6 |
| | B8A44FDF-9CB9-4123-9AB0-4BC198921FEE |
| | B8A8D2FC-421B-4393-8D5A-946CF72CB496 |
| | B8AEFC48-4A6E-4254-A57F-5F688399B582 |
| | B8CB385A-4E5F-4932-944D-04D6EA1BBA5A |
| | B8DD67AE-5808-45FC-9A1F-47F18A2F827D |
| | b8fbfe87-e92c-4d54-a620-17b2dcde6d53 |
| | B9E6A966-DA7C-46F3-825F-8E19C06518C3 |
| | ba39bfbc-5cac-4f6c-b55c-4eee3466ee93 |
| | BA62B5EB-E169-4B41-B949-E004D66E98A1 |
| | BA7B9B84-C227-4AD0-9E71-88EE3309162C |
| | BA80DB4E-D899-4DA4-AE49-5263D98E1530 |
| | BAB3CE0B-D318-4791-AA17-D815EA23B62D |
| | BAC49A26-1556-45B8-8729-A05DF52623D8 |
| | BC282A61-9E8B-4330-9177-FC515293F051 |
| | BD75D8EE-916B-4ABC-BF33-BCE6A4217076 |
| | BDD08EC8-8892-4273-8F21-6FDE0F23C52D |
| | BE6531B2-D1F3-44AB-9C02-1CEAE51EF2BB |
| | BE8FA67D-C2D3-4FFF-82FE-A46BA1FA8DF0 |
| | bfac45a6-fa01-4edb-9807-6898ea82e80e |
| | bfd49783-1767-469b-9d79-7822301c5efc |
| | C02699B5-580A-4D55-971E-F4692B2CCE57 |
| | C047E741-2B29-45DF-A577-351BDDC50EB8 |
| | C0B7B798-3383-4A45-A455-9ECA5810739E |
| | C0BB1051-09B2-46F3-A22B-FE313B1A6A4E |
| | C14468EB-E725-4029-96CC-75BB9EBD9DEF |
| | C14A0460-30B0-453B-AB81-DA03AD4AD02E |
| | C16EED96-8DA8-4D01-B87D-660F46172777 |
| | C179D6B1-6182-48B8-A376-3133E0DEC278 |
| | c30eb9bd-d705-4464-9014-e0aa7cda53c7 |
| | C45E02E5-7E3C-4F36-8B8D-F54617E8A436 |
| | C4F61715-EE58-4799-A5F4-0F5E27C4093D |
| | C53167D4-14DA-40CC-B30A-35EB3967B8DF |
| | C53C4BFE-D9D2-4882-A46B-FA487D95B8BA |
| | C57835BE-77AB-4CF5-9B23-62C11A7B2EAF |
| | C5A16472-AC62-422B-A799-26D17AEAA38E |
| | C5D08BE7-B94C-423A-AEA8-BBC635787C63 |
| | C5F8C89C-EF69-42F4-9B37-5B552FCCDF4B |
| | C60E89A7-A8BA-458E-8F10-785C3F562FD6 |
| | C72CB184-462D-4009-9CDB-848782FF8A76 |
| | C739FD61-22B2-412D-BCF3-89BDA45A2C0F |
| | C77976B6-35EA-4623-A92A-6D9440A978E6 |

| | |
|---|---|
| | C7912CA7-11E1-4CB8-902A-9D6CB23FD344 |
| | C8388AC0-975D-4F1E-8324-8CABDC75738D |
| | C83FB5E6-8AF3-47E4-AE48-CEE92DFF9DB7 |
| | C849127A-2F15-4FAB-8815-DB09B017F48C |
| | C85F340E-584B-4F3B-B6A5-540491FC8AD2 |
| | C871EA6B-FBD2-4E6F-AB29-F43F6E3DDCBB |
| | C8744498-5D0E-4090-A6F1-C84AA6BB2BCB |
| | C8F39325-5382-447C-9291-A6915FC978B8 |
| | C93C47F4-88F3-405A-85F4-BA9450B74522 |
| | C9A5B051-0503-4AAE-8BC6-270AF3033326 |
| | C9F519B6-73F1-4737-83CB-F5EF208592CD |
| | CA076CFD-C628-4AF2-837C-37617429ADB9 |
| | CA20249F-B7EA-4FD9-9ECB-34F74755AE35 |
| | CA242B4C-B879-4E13-A1E7-84C984067189 |
| | CA65A38E-8B49-4B50-908A-478B95B95C02 |
| | cb0a2055-2168-4454-8755-47dd613db36f |
| | CB31B3CE-8595-4D75-9436-BB59E95C3A19 |
| | CB4394EC-1991-460D-B823-B74E005E3DA7 |
| | CB78EFB3-BC29-4DE7-9128-FDC4F9539B48 |
| | CBAB7A72-11F7-4DFE-80F8-1B1EF1CF0055 |
| | CC14603B-E32C-43F3-8218-B6ED7EE2EC18 |
| | cc348a26-ee11-47a4-8b51-de922967e175 |
| | CCA74404-CD91-4A69-AE09-BFB5D9142D3D |
| | CCD672DF-44D4-421F-8727-0A05E513B6C8 |
| | CCF7073E-B224-497A-A476-C67AFF5226BD |
| | CCF8809A-CDFD-46F7-B607-001B8B91C1A4 |
| | CD0C33C3-4427-471D-979E-79DC43F3A018 |
| | CD1656D8-55A5-4248-B81D-FED9D5992CBE |
| | CEF2EFB2-93CC-4BDF-A343-9965E7E0432C |
| | CF2A1B98-5693-4C04-9167-F85E69A88A4A |
| | CF87812F-6DB6-4592-9842-C62B3E4FF03F |
| | cf9db1af-17f0-490e-8139-142bd704763a |
| | CFCF057A-00FE-4603-A720-586712B75803 |
| | CFDC8F4D-A25B-4C63-9832-58444581A505 |
| | D0680568-BBB3-4561-9E4B-08FE2A4832CB |
| | D0CBE668-5C95-4261-A1E8-CEF55093A0F7 |
| | D0D989CB-A50F-4ED9-8478-AE300EBC0970 |
| | D112136D-34FA-426E-B432-4D1E4286ADF8 |
| | D13AC936-87A9-4164-BDAB-02CCF3908CFE |
| | D13FB44B-291B-4EA4-920C-142DAA8D1989 |
| | D22DE643-5F93-42D0-9199-1F55405241D6 |
| | d296cdb5-a0f6-40cc-8ce7-73e4ccd176f3 |
| | D2CC4D31-38FA-4232-895E-767260F607E9 |
| | D2DF20D4-6704-41E6-B33D-616225BD737C |
| | D383008A-A1BF-4689-B243-3C464C79B818 |
| | d39d1a49-fcc0-4f2c-9338-df904e7f6a09 |
| | D3B77DA8-BE1D-4955-B1C7-55B7769785E8 |
| | D3D3DBA9-139E-4F57-AC70-B741553D1687 |
| | D3D9CCDD-4789-43B0-8E73-B2C42CB7D83D |
| | D3E75FB4-FB7C-4EBF-9C75-2BE9FFD9F52D |
| | D3F786A6-5D72-4D6E-85FD-0C6ADE9F6E95 |
| | D4355111-3ABB-40F0-BDB7-B9EAA39DC771 |
| | D487536F-0521-4D42-9150-6A806AE00241 |
| | D48E9F25-F083-4F88-A92D-C12D9F765BA6 |
| | d4a1ecdc-399c-4f82-aa7a-68a0c82ded3f |
| | D542AA45-86CF-462E-ABBE-76A3354E707E |
| | D5932DD7-B777-427F-8B55-3EC369FD76A3 |
| | d5e14ef5-1b78-4498-a2e2-77070c83eb7f |

| | |
|---|---|
| | D5F2B85A-94A9-4168-BBE8-149EC71342B0 |
| | d622bcab-5b5a-45a4-8c02-7bcfdbbb781c |
| | d67cd793-2931-429a-9084-2f3c4c8be7ad |
| | D684399E-46C4-4BAD-92E9-3DE17C9F9A5F |
| | D6974DB6-2BD3-4082-B48E-9C60F7FE3A1F |
| | D69E29CA-24BE-4F35-A581-F82E624B8E8B |
| | D6B1516A-8271-4071-8C56-D4538DE63892 |
| | D6F7AFC0-1558-43AD-ACB1-2B5311ED2264 |
| | D78012B6-24F9-48F5-9409-9845D0BE2B3C |
| | D7D793F6-EAEA-4DD3-8237-C2C02B042340 |
| | D7D9204B-B85E-4260-8A54-C39E7847E982 |
| | D81594D9-DD6A-45C3-8BAD-61960611D78D |
| | d83b6b2d-932b-4240-b99d-5506f18af892 |
| | D84312E8-999D-49AD-B0AA-E48E87EFC462 |
| | D8D53DF1-42D6-4D8B-AC4C-6F3993AC247D |
| | D8D54C81-7EF5-4B0F-B35B-28FBF8C5198E |
| | d8d7a6c2-6427-4f47-968c-6c3affba4617 |
| | D947828F-D62F-426B-9710-3C93A7AEF05B |
| | D9C2CA21-F1A6-4CF4-BE28-6C2492DD38E5 |
| | D9DC3B59-613D-469E-8B4F-6C5A557EB26A |
| | D9F4A2C1-AC6D-48C6-95FE-FFBC2FA08268 |
| | DA13446A-C4DC-4D02-80C8-410273925AC4 |
| | DA40E6D3-A2A9-4517-AAA2-AEF6C3892EBF |
| | da9c10ea-c13f-4d88-8b94-a0cbc106d036 |
| | DAA9292E-8012-4AA2-8221-AD65FB1C7B78 |
| | DADCF6D5-03EB-4090-BCC3-B9E9DC3C7E08 |
| | DAF211F2-941E-4244-8CF5-D31492269443 |
| | DAFB9844-1A5C-4AF8-B382-59C751942BB8 |
| | DB67A9CD-65BA-4D6F-A799-031B6B3DF599 |
| | DC4062D7-1C81-4B19-AABD-8307A0DF5029 |
| | DC659941-8D12-4095-8D8A-1769878C363C |
| | dcfd0b1d-5707-44a6-b225-5e8f7cea5ca1 |
| | DD305533-E48C-4FED-9283-5082703C23FF |
| | DD43FFD9-DA18-4AD3-ABE7-6F443C76A717 |
| | DD478399-D2EC-4AE4-A82A-B0258D24D088 |
| | DE509AA1-6F20-480F-BF9D-D6BC04A7F72C |
| | DE5ABE44-6D9D-4797-A57A-5ADFD71383F0 |
| | DE9315B3-B3DF-45F0-ADC2-69AA6B8FF717 |
| | DEB57BD8-545F-477A-9A86-6BC6DB257270 |
| | DF15901C-2508-42C9-BF06-026EFD89E51F |
| | dfefb76a-ec6b-4cd2-9d45-2e1e4befc7ea |
| | DFFEF9E8-5B69-43D9-A95B-0744DAA38D1D |
| | E0174496-D774-4417-81D1-CB16771760AD |
| | E03C99F2-93CB-457F-8A77-D600F292E313 |
| | E069DCAD-2FD1-464E-A5E9-FE36F05A0446 |
| | E08FF4F7-62FD-4240-9E8B-C3AD7A9B2C44 |
| | E0A5D99B-B58B-4301-B2DC-A767D90059B1 |
| | E0AB95EF-5C96-4D00-8950-04E26E3B4672 |
| | E0DF1FEF-C650-4088-A974-26F712ED4F64 |
| | E0DFC37A-4A2E-4353-9673-241B100B3467 |
| | E15D92A1-6925-4B58-A494-D6F665F67B83 |
| | E1786E78-AC46-46FD-8CE0-8F7456E46082 |
| | E1FF7694-64E1-47ED-B0D8-34ACFB4FFEF1 |
| | E2272DE4-02B8-4A94-B8EE-C7454996A2DA |
| | e23cd75a-06ad-4da0-b19b-b812b0f6c1b9 |
| | E27BC39B-54F1-47B5-AC8F-3245DCE690F4 |
| | E30F1994-3622-43D0-BEC2-6D7E2D9881D2 |
| | E3121DDE-EEE2-4975-A135-BDB89434A7E2 |

| | |
|---|---|
| | E35B2928-8784-45D0-A71A-A2DF161542FA |
| | E48A8FA7-4E2B-44A0-95F5-E2925C60BAD3 |
| | E4BCCEA0-0104-40C4-9E3C-2AD7262D918E |
| | E5086EAB-C920-4E3E-A965-90F3F495363A |
| | E5D64754-15E8-494E-B7F7-25CF2819FE4C |
| | e5e4aa9f-6015-4d62-a088-69920d7f2500 |
| | E6183A43-24B9-4FE1-95C3-4CFF67E6C1ED |
| | e61fb24b-613a-4b6d-bb2b-00ccedbd3dbc |
| | E62ADAC4-803A-4D94-9AE0-7B1A49041799 |
| | E656CF38-2F5D-4306-90D9-ECB504D86351 |
| | e6b72c24-1607-43b9-8b8a-7bf83eea5895 |
| | E6F7BF6C-92B1-4795-94FC-B189D0C7877F |
| | E7174299-3CA7-4D51-9048-3EF3EFDE4E5C |
| | E77C3F70-08D1-40D3-B038-F09A519423CC |
| | E783E518-C1E5-4EAC-8EAF-E2D65CCD9692 |
| | e7a00d67-2c26-4d1f-bd17-35f659e88bc1 |
| | E7A72F7E-77B1-4D95-8F37-39A6CFFC35DC |
| | e7bae6e7-f779-449d-92b9-66996041e00d |
| | E7C7783C-1D0F-4F42-A4CF-518825E91D91 |
| | e89a9deb-6321-4e61-b412-b4993d4277dd |
| | E89D7900-915D-4936-B1B2-CAC3670EA2F4 |
| | E89F7631-FDAC-4BE9-9997-70E400EFDC04 |
| | e8ac39e6-377a-412c-b18d-f6c4fc654ec6 |
| | E8EA576B-EB16-44F7-B241-DAEFC1375388 |
| | E9002F4E-BA0B-4738-9368-DD9614A5DA8A |
| | E90BFE17-2E5B-47ED-B986-B3D4181F2BF6 |
| | E94DFD42-4FE4-48E9-AA62-422AE071413F |
| | e98b69bc-766a-485f-b5e8-9166b8d457b3 |
| | E9949320-AD37-4542-98C1-B72DF60B1668 |
| | E9A12DF9-024E-4CBC-B6BF-E4E87485FA90 |
| | E9A42A85-9513-41B2-A71D-6D62F70B00E4 |
| | ea93555b-daa5-439a-a13a-796917365454 |
| | EA953CE5-7082-41FF-8E38-760FE4100414 |
| | EB0EB159-732E-43FF-A402-7F2D9F67DAA8 |
| | EB642568-30D4-4805-A295-BDA314CCAAAF |
| | ebd1be2c-35ca-45c0-9aba-9baf432114c1 |
| | EBFB8C1B-471F-4ACB-A1FE-287E02CADBA7 |
| | EC0AB947-9341-4FFF-BDA4-FDFB9434D508 |
| | EC461EAF-F5CB-4225-A9A0-59D8E6C17EA6 |
| | EC4C4296-6AB4-420C-80A4-E7A34B9BF4A2 |
| | EC4CC744-04DF-4BA9-9159-E4940C702281 |
| | ED17685C-B706-41E4-8A76-11CB0E6700CD |
| | ed879063-e603-4151-bf5e-22a4ee210281 |
| | EDA9496E-BE80-4A13-BF06-89F0CC9E937F |
| | EDB90376-0EF9-47DD-BE6F-A6E8A0380A9C |
| | EDBF86E5-3F55-4DDF-A97F-3E9DC607E53D |
| | EDE2E8A7-465E-4725-880D-519899ABA0C0 |
| | EE08EDA6-033E-403B-A26A-747B118AE897 |
| | EE2C9743-EAA1-4F0D-8D35-E96686AC3913 |
| | EE6083AA-9453-44FF-8CC5-79B71FEA289D |
| | EEA7D707-AEB9-4D00-AB5E-C1EA608517E9 |
| | EEAA0FA3-408C-4E0F-A18A-095514173FF1 |
| | EF306507-F426-499E-8A1F-3401A522FB6B |
| | EF3DD880-7BE4-46A8-93C2-7CBA1A9CD24D |
| | EF573329-9659-4106-B391-9A33E1C732C4 |
| | EF8BEC6F-F594-4076-9F38-157165633EF1 |
| | EFBDEDAA-4B02-40AC-B93C-9670C018CF7F |
| | efe8b105-b170-4679-8b97-c1142fefe069 |

| | |
|---|---|
| | EFF14FED-8A21-41C5-8ED3-75346A06E44E |
| | EFFB5839-24A2-4B8C-8B6D-08CB66503D16 |
| | F04F1C1C-6D6D-4A2B-BA88-110B08FCC596 |
| | f090838e-7cc0-4ba8-b3a2-faaa99743785 |
| | F0BC4F65-211B-4AE1-8E68-AAEF3F3C6DB8 |
| | F0CFAACC-30E5-4E54-B33A-F0FD5DAD620E |
| | F0DE67FF-8967-4FCE-9C12-0E07193285CD |
| | F1099653-FD32-4459-86EB-40E0412CE2FA |
| | F11B1E99-C14F-442E-ACAB-6353A5C77266 |
| | F13DFDCC-02F8-4A4C-A2FC-E6A45499BD4C |
| | f1b7b418-80d2-4ddf-94cd-f79abba0a2a4 |
| | F248D7B9-6EA7-44EF-8AF6-8BEDA6C6BA4A |
| | F24ED9C0-92BB-4003-86D2-42AB26617A05 |
| | f251cb6c-d18e-4691-9c82-24ed1ac0abf9 |
| | f25bae1a-8627-46a8-87af-07f82773c68a |
| | F2622CDB-AC92-42B3-A1CA-67E133C3DD4A |
| | f2a8deb2-0e2c-4bbd-8fa1-99b7abb2f3aa |
| | f2ad720e-579f-4e2c-a3f0-17877ca91067 |
| | F2BEA37E-0D19-49E1-91A0-A4DE4DFA2D35 |
| | F35A0655-7504-4BD6-A9F3-6F3E59FD84B1 |
| | F3665B89-957B-4D7B-B67F-39615C47D28B |
| | f383df81-a334-43c6-a5be-ea00013a4f6a |
| | f384eb3f-92ab-4327-87eb-45b5f72a1580 |
| | f3aa40b9-7b4d-465a-b7c9-591aebdb56c2 |
| | f3ba71f9-25f3-4784-bf2d-3aa522a0cba8 |
| | F3CBADB3-3B83-47CD-B8A0-B0E10343A295 |
| | F3EC5BF3-97AE-4033-91A6-084A1AA152A4 |
| | f4350d9c-3f40-4829-bbc4-8acb2f3ff512 |
| | f5539207-6534-4edc-a5a5-7666b5d16ba4 |
| | f59b9c65-46e8-4cc3-aa71-8844f6c2273c |
| | F5D90810-8C4B-4CED-9360-383E42CB54B3 |
| | F5F4B937-8B81-4BE5-B93E-B3224E357C15 |
| | f6381367-142c-45d0-92b3-c1727d1813ce |
| | f68237e7-7fc0-4b6b-8095-b49515ddd169 |
| | F69FF5FB-7E0B-4B16-9FC9-498379B11562 |
| | f6ebe511-a4df-4310-bd59-3a46d43f100b |
| | F6EEBD4B-B63A-4A9C-92D3-0D954A8A6655 |
| | F7C891EB-3B09-4E01-BB8A-53D570518CBE |
| | F852DE27-B600-44A4-AB30-D74108DFC2E9 |
| | F854CE67-C586-4424-A674-2DD67AD0ED7F |
| | f87f7cee-e9bd-45cc-a740-3d8ffa9746d3 |
| | F88EC55C-4C1E-4593-9C89-1981F220C0F8 |
| | F89588E9-CA73-4465-A7FB-7246EDB45E3A |
| | F8B98552-AE2A-4609-9929-7864485B3C51 |
| | f947d67f-4aa2-4eb9-977e-21a5ae563613 |
| | F9A7AB94-70DF-4194-8102-66021D01DADD |
| | F9B55748-605A-4313-BB19-DCF5476590C2 |
| | F9DB9757-04D8-40BD-A58D-605F4255B0E6 |
| | f9eb88f7-9293-46bf-ace4-a746e4ff80dc |
| | FA4E082A-D213-412E-8D91-842A6F4BEF12 |
| | FAE3DBD7-1C4E-45C0-8187-C09892279565 |
| | FB143117-C3AD-4F4B-8F39-7586279E8EAA |
| | FB14BD0D-B356-42A4-A0E9-2FCDADA8F76B |
| | FBFED398-2A44-44A9-83CF-657C29CB7D28 |
| | FC2BCE29-4685-49F5-B901-2117BF673BB8 |
| | FCB0C5E5-F63C-41B3-AE0E-3D1D6AAA66E4 |
| | FCF14E9D-4671-453D-B8E0-5008E916F56D |
| | FD7E2226-8086-4A56-867B-6457B87A7FED |

| | |
|---|---|
| | FDFF46A7-F92A-45CF-8756-6C73A406C923 |
| | FE1AD544-3189-4495-86BE-0C68F3234238 |
| | FE28F76E-7EAD-4D29-8C53-A576AE6021F7 |
| | fec0da58-1047-44d2-b6d1-c18cceed43dc |
| | FEF9C64F-5959-4DA0-AAA2-66B56FC7B4C3 |
| | ff004403-8cc5-43e6-bd1c-dde4eb1e6193 |
| | FF38628E-8ABE-44D6-8D3B-7CFE46BD2E35 |
| | FF49E0F9-4007-4761-845E-B676B0FC9646 |
| | FF52FA2F-BD12-4DE0-9F5C-CD330FFB5FB3 |
| | FF6B5FC8-0572-4B58-B3A5-BCDA41BADBC8 |
| | FFCFA005-A04F-458E-9D1D-86143DD823E5 |

| treatment_best_response | Count of bcr_ radiation_barcode |
|---|---|
| Complete Response | 1360 |
| Partial Response | 140 |
| Radiographic Progressive Disease | 293 |
| Stable Disease | 206 |
| Grand Total | 1999 |

| | | | After removing duplicate Ids and common Ids |
|---|---|---|---|
| | Total cases numbers | case numbers common in RR & RS | Unique case numbers |
| RS | 1500 | | 1052 |
| RR | 499 | | 393 |
| Common | | 25 | |
| Total | 1999 | | 1445 |

| Primary site | Cancer type | No. of cases | | | GWAS | Transcriptomics |
|---|---|---|---|---|---|---|
| | | Radiosensitive | Radioresistant | Total | | |
| Thyroid | THCA | 187 | 13 | 200 | 0 | 2 |
| Breast | BRCA | 175 | 12 | 187 | 0 | 1 |
| Larynx | HNSC | 154 | 17 | 171 | 22 | 1 |
| Brain | LGG | 43 | 124 | 167 | 11 | 0 |
| Cervix | CESC | 87 | 17 | 104 | 0 | 3 |
| Prostate | PRAD | 51 | 16 | 67 | 1 | 1 |
| Stomach | STAD | 41 | 15 | 56 | 0 | 85 |
| Uterus | UCEC | 49 | 7 | 56 | 0 | 4 |
| Esophagus | ESCA | 35 | 14 | 49 | 0 | 0 |
| Skin | SKCM | 24 | 23 | 47 | 0 | 0 |
| Bronchus & Lung | LUAD | 17 | 29 | 46 | 0 | 0 |
| Connective_Subcutaneous-SARC | SARC | 16 | 18 | 34 | 0 | 11 |
| Pancreas | PAAD | 14 | 14 | 28 | 2 | 0 |
| Bladder | BLCA | 16 | 12 | 28 | 0 | 0 |
| Bronchus & Lung | LUSC | 14 | 13 | 27 | 0 | 0 |
| Thymus | THYM | 24 | 3 | 27 | NA | NA |
| Testis | TCGT | 24 | 0 | 24 | NA | NA |
| Uterus | UCS | 17 | 5 | 22 | NA | NA |
| Heart | MESO | 11 | 6 | 17 | NA | NA |
| Rectum | READ | 15 | 0 | 15 | NA | NA |
| Brain | GBM | 4 | 8 | 12 | NA | NA |
| Colon | COAD | 1 | 9 | 10 | NA | NA |
| Liver & Bile | LIHC | 3 | 6 | 9 | NA | NA |
| Adrenal gland | ACC | 2 | 6 | 8 | NA | NA |
| Lymph | DLBC | 6 | 2 | 8 | NA | NA |
| Kidney | KICH | 1 | 4 | 5 | NA | NA |
| Adrenal | PCPG | 1 | 3 | 4 | NA | NA |
| Eye | UVM | 1 | 1 | 2 | NA | NA |
| Gall Bladder | CHOL | 0 | 1 | 1 | NA | NA |

**Figure 8a- gene-counts in GWAS & Transcriptomics graph**
