## Supplementary table_2 for "Direct Pan-Cancer Multi-Omic Analysis of Radiotherapy Response Reveals Coding and Regulatory Genomic Variation, and Extracellular Adaptive Programs Underlying Radioresistance"

| High/Moderate Impact Variants | | | | | | | | | | | |
|---|---|---|---|---|---|---|---|---|---|---|---|
| **RR** | | | | | | | | | | | |
| **HNSC** | | | | | | | | | | | |
| **Location** | **Allele** | **Consequence** | **IMPACT** | **SYMBOL** | **Gene** | **Feature_type** | **Feature** | **BIOTYPE** | **Existing_variatio** | **CANONICAL** | **MANE** |
| **BLGG** | | | | | | | | | | | |
| 1:214383705-2143 | - | inframe_deletion | MODERATE | PTPN14 | ENSG0000015210 | Transcript | ENST0000036695 | protein_coding | - | YES | MANE_Select |
| 1:225847138-2258 | - | inframe_deletion | MODERATE | TMEM63A | ENSG0000019618 | Transcript | ENST0000036683 | protein_coding | - | YES | MANE_Select |
| 10:128106847-128 | - | frameshift_variant | HIGH | MKI67 | ENSG0000014877 | Transcript | ENST0000036865 | protein_coding | - | YES | MANE_Select |
| 16:50334766-5033 | - | frameshift_variant | HIGH | BRD7 | ENSG0000016616 | Transcript | ENST0000039468 | protein_coding | - | YES | MANE_Select |
| 19:38408861-3840 | - | inframe_deletion | MODERATE | FAM98C | ENSG0000013024 | Transcript | ENST0000025253 | protein_coding | - | YES | MANE_Select |
| **Adenocarcinoma** | | | | | | | | | | | |
| 1:6469122-646913 | - | inframe_deletion | MODERATE | PLEKHG5 | ENSG0000017168 | Transcript | ENST0000037772 | protein_coding | - | YES | MANE_Select |
| 11:115209591-115 | - | inframe_deletion | MODERATE | CADM1 | ENSG0000018298 | Transcript | ENST0000033158 | protein_coding | - | YES | MANE_Select |
| 13:99982753-9998 | - | inframe_deletion | MODERATE | ZIC2 | ENSG0000004335 | Transcript | ENST0000037633 | protein_coding | - | YES | MANE_Select |
| 16:72788664-7278 | - | inframe_deletion | MODERATE | ZFHX3 | ENSG0000014083 | Transcript | ENST0000026848 | protein_coding | - | YES | MANE_Select |
| 16:72797458-7279 | - | inframe_deletion | MODERATE | ZFHX3 | ENSG0000014083 | Transcript | ENST0000026848 | protein_coding | - | YES | MANE_Select |
| 20:47238822-4723 | - | inframe_deletion | MODERATE | ZMYND8 | ENSG0000010104 | Transcript | ENST0000047195 | protein_coding | - | YES | MANE_Select |
| 8:28351708-28351 | - | inframe_deletion | MODERATE | ZNF395 | ENSG0000018691 | Transcript | ENST0000034442 | protein_coding | - | YES | MANE_Select |

| High/Moderate Impact Variants | | | | | | | | | | | |
|---|---|---|---|---|---|---|---|---|---|---|---|
| **RS** | | | | | | | | | | | |
| **HNSC** | | | | | | | | | | | |
| **Location** | **Allele** | **Consequence** | **IMPACT** | **SYMBOL** | **Gene** | **Feature_type** | **Feature** | **BIOTYPE** | **Existing_variatio** | **CANONICAL** | **MANE** |
| 10:128106847-128106851 | - | frameshift_variant | HIGH | MKI67 | ENSG0000014877 | Transcript | ENST0000036865 | protein_coding | - | YES | MANE_Select |
| **BLGG** | | | | | | | | | | | |
| 4:53453080-53453084 | - | frameshift_variant | HIGH | FIP1L1 | ENSG0000014521 | Transcript | ENST0000033748 | protein_coding | - | YES | MANE_Select |
| 8:28351708-28351714 | - | inframe_deletion | MODERATE | ZNF395 | ENSG0000018691 | Transcript | ENST0000034442 | protein_coding | - | YES | MANE_Select |
| **Adenocarcinoma** | | | | | | | | | | | |
| 1:6469122-6469128 | - | inframe_deletion | MODERATE | PLEKHG5 | ENSG0000017168 | Transcript | ENST0000037772 | protein_coding | - | YES | MANE_Select |
| 10:30027571-30027577 | - | inframe_deletion | MODERATE | JCAD | ENSG0000016575 | Transcript | ENST0000037537 | protein_coding | - | YES | MANE_Select |
| 11:115209591-115209606 | - | inframe_deletion | MODERATE | CADM1 | ENSG0000018298 | Transcript | ENST0000033158 | protein_coding | - | YES | MANE_Select |
| 12:8047961-8047967 | - | inframe_deletion | MODERATE | FOXJ2 | ENSG0000006597 | Transcript | ENST0000016239 | protein_coding | - | YES | MANE_Select |
| 13:99982753-99982768 | - | inframe_deletion | MODERATE | ZIC2 | ENSG0000004335 | Transcript | ENST0000037633 | protein_coding | - | YES | MANE_Select |
| 14:24163922-24163928 | - | inframe_deletion | MODERATE | IRF9 | ENSG0000021392 | Transcript | ENST0000039686 | protein_coding | - | YES | MANE_Select |
| 14:24300643-24300649 | - | inframe_deletion | MODERATE | NOP9 | ENSG0000019694 | Transcript | ENST0000026742 | protein_coding | - | YES | MANE_Select |
| 15:40036395-40036401 | - | inframe_deletion | MODERATE | SRP14 | ENSG0000014031 | Transcript | ENST0000026788 | protein_coding | - | YES | MANE_Select |
| 16:50334766-50334770 | - | frameshift_variant | HIGH | BRD7 | ENSG0000016616 | Transcript | ENST0000039468 | protein_coding | - | YES | MANE_Select |
| 19:1578372-1578378 | - | inframe_deletion | MODERATE | MBD3 | ENSG0000007165 | Transcript | ENST0000043443 | protein_coding | - | YES | MANE_Select |
| 2:232378818-232378824 | - | inframe_deletion | MODERATE | ALPP | ENSG0000016328 | Transcript | ENST0000039202 | protein_coding | - | YES | MANE_Select |
| 20:23365283-23365289 | - | inframe_deletion | MODERATE | GZF1 | ENSG0000012581 | Transcript | ENST0000033812 | protein_coding | - | YES | MANE_Select |
| 20:34743163-34743169 | - | inframe_deletion | MODERATE | NCOA6 | ENSG0000019864 | Transcript | ENST0000035900 | protein_coding | - | YES | MANE_Select |
| 22:42214566-42214572 | - | inframe_deletion | MODERATE | TCF20 | ENSG0000010020 | Transcript | ENST0000067762 | protein_coding | - | YES | MANE_Select |
| 3:65439885-65439891 | - | inframe_deletion | MODERATE | MAGI1 | ENSG0000015127 | Transcript | ENST0000040293 | protein_coding | - | YES | MANE_Select |
| 4:53453080-53453084 | - | frameshift_variant | HIGH | FIP1L1 | ENSG0000014521 | Transcript | ENST0000033748 | protein_coding | - | YES | MANE_Select |

| High/Moderate Impact Variants | | | | | | | | | | | |
|---|---|---|---|---|---|---|---|---|---|---|---|
| **RR** | | | | | | | | | | | |
| **HNSC** | | | | | | | | | | | |
| **Location** | **Allele** | **Consequence** | **IMPACT** | **SYMBOL** | **Gene** | **Feature_type** | **Feature** | **BIOTYPE** | **Existing_variatio** | **CANONICAL** | **MANE** |
| 1:6469122-646912 | - | inframe_deletion | MODERATE | PLEKHG5 | ENSG0000017168 | Transcript | ENST0000037772 | protein_coding | rs113541584 | YES | MANE_Select |
| 10:128106847-128 | - | frameshift_variant | HIGH | MKI67 | ENSG0000014877 | Transcript | ENST0000036865 | protein_coding | rs145960091 | YES | MANE_Select |
| 13:99982753-9998 | - | inframe_deletion | MODERATE | ZIC2 | ENSG0000004335 | Transcript | ENST0000037633 | protein_coding | rs398124241 | YES | MANE_Select |
| 14:73105898-7310 | - | frameshift_variant | HIGH | RBM25 | ENSG0000011970 | Transcript | ENST0000026197 | protein_coding | rs150988201 | YES | MANE_Select |
| 17:40822513-4082 | - | inframe_deletion | MODERATE | KRT10 | ENSG0000018639 | Transcript | ENST0000026957 | protein_coding | rs148510452 | YES | MANE_Select |
| 2:226796679-2267 | - | frameshift_variant | HIGH | IRS1 | ENSG0000016904 | Transcript | ENST0000030512 | protein_coding | CD951753 | YES | MANE_Select |
| 6:161098318-1610 | - | inframe_deletion | MODERATE | MAP3K4 | ENSG0000008551 | Transcript | ENST0000039214 | protein_coding | rs5881391 | YES | MANE_Select |
| 9:117214709-1172 | - | inframe_deletion | MODERATE | ASTN2 | ENSG0000014821 | Transcript | ENST0000031340 | protein_coding | rs750505669 | YES | MANE_Select |
| X:151648669-151 | - | inframe_deletion | MODERATE | PASD1 | ENSG0000016604 | Transcript | ENST0000037035 | protein_coding | rs750036801 | YES | MANE_Select |
| **BLGG** | | | | | | | | | | | |
| 1:214383705-2143 | - | inframe_deletion | MODERATE | PTPN14 | ENSG0000015210 | Transcript | ENST0000036695 | protein_coding | - | YES | MANE_Select |
| 1:225847138-2258 | - | inframe_deletion | MODERATE | TMEM63A | ENSG0000019618 | Transcript | ENST0000036683 | protein_coding | - | YES | MANE_Select |
| 10:128106847-128 | - | frameshift_variant | HIGH | MKI67 | ENSG0000014877 | Transcript | ENST0000036865 | protein_coding | - | YES | MANE_Select |
| 16:50334766-5033 | - | frameshift_variant | HIGH | BRD7 | ENSG0000016616 | Transcript | ENST0000039468 | protein_coding | - | YES | MANE_Select |
| 19:38408861-3840 | - | inframe_deletion | MODERATE | FAM98C | ENSG0000013024 | Transcript | ENST0000025253 | protein_coding | - | YES | MANE_Select |
| 8:28351708-28351 | - | inframe_deletion | MODERATE | ZNF395 | ENSG0000018691 | Transcript | ENST0000034442 | protein_coding | rs547945591 | YES | MANE_Select |
| **PRAD** | | | | | | | | | | | |
| 16:50334766-5033 | - | frameshift_variant | HIGH | BRD7 | ENSG0000016616 | Transcript | ENST0000039468 | protein_coding | rs145896392 | YES | MANE_Select |
| **Adenocarcinoma** | | | | | | | | | | | |
| 1:6469122-646913 | - | inframe_deletion | MODERATE | PLEKHG5 | ENSG0000017168 | Transcript | ENST0000037772 | protein_coding | - | YES | MANE_Select |
| 10:30027571-3002 | - | inframe_deletion | MODERATE | JCAD | ENSG0000016575 | Transcript | ENST0000037537 | protein_coding | rs34594193 | YES | MANE_Select |
| 11:115209591-115 | - | inframe_deletion | MODERATE | CADM1 | ENSG0000018298 | Transcript | ENST0000033158 | protein_coding | - | YES | MANE_Select |
| 12:8047961-80479 | - | inframe_deletion | MODERATE | FOXJ2 | ENSG0000006597 | Transcript | ENST0000016239 | protein_coding | rs372118289 | YES | MANE_Select |
| 13:99982753-9998 | - | inframe_deletion | MODERATE | ZIC2 | ENSG0000004335 | Transcript | ENST0000037633 | protein_coding | - | YES | MANE_Select |
| 14:24163922-2416 | - | inframe_deletion | MODERATE | IRF9 | ENSG0000021392 | Transcript | ENST0000039686 | protein_coding | rs746378882 | YES | MANE_Select |
| 14:24300643-2430 | - | inframe_deletion | MODERATE | NOP9 | ENSG0000019694 | Transcript | ENST0000026742 | protein_coding | rs71119069 | YES | MANE_Select |
| 15:40036395-4003 | - | inframe_deletion | MODERATE | SRP14 | ENSG0000014031 | Transcript | ENST0000026788 | protein_coding | rs371085676 | YES | MANE_Select |
| 16:50334766-5033 | - | frameshift_variant | HIGH | BRD7 | ENSG0000016616 | Transcript | ENST0000039468 | protein_coding | rs145896392 | YES | MANE_Select |
| 16:51141744-5114 | - | inframe_deletion | MODERATE | SALL1 | ENSG0000010344 | Transcript | ENST0000025102 | protein_coding | rs113614842 | YES | MANE_Select |
| 16:72788664-7278 | - | inframe_deletion | MODERATE | ZFHX3 | ENSG0000014083 | Transcript | ENST0000026848 | protein_coding | - | YES | MANE_Select |
| 16:72797458-7279 | - | inframe_deletion | MODERATE | ZFHX3 | ENSG0000014083 | Transcript | ENST0000026848 | protein_coding | - | YES | MANE_Select |
| 19:1578372-15783 | - | inframe_deletion | MODERATE | MBD3 | ENSG0000007165 | Transcript | ENST0000043443 | protein_coding | rs371220154 | YES | MANE_Select |
| 2:232378818-2323 | - | inframe_deletion | MODERATE | ALPP | ENSG0000016328 | Transcript | ENST0000039202 | protein_coding | rs377162921 | YES | MANE_Select |

| | | | | | | | | | | | |
|---|---|---|---|---|---|---|---|---|---|---|---|
| 20:23365283-2336 | - | inframe_deletion | MODERATE | GZF1 | ENSG0000012581 | Transcript | ENST0000033812 | protein_coding | rs750609719 | YES | MANE_Select |
| 20:34743163-3474 | - | inframe_deletion | MODERATE | NCOA6 | ENSG0000019864 | Transcript | ENST0000035900 | protein_coding | rs112051697 | YES | MANE_Select |
| 20:47238822-4723 | - | inframe_deletion | MODERATE | ZMYND8 | ENSG0000010104 | Transcript | ENST0000047195 | protein_coding | - | YES | MANE_Select |
| 3:65439885-65439 | - | inframe_deletion | MODERATE | MAGI1 | ENSG0000015127 | Transcript | ENST0000040293 | protein_coding | rs142043619 | YES | MANE_Select |
| 4:53453080-53453 | - | frameshift_variant | HIGH | FIP1L1 | ENSG0000014521 | Transcript | ENST0000033748 | protein_coding | rs143671659 | YES | MANE_Select |
| 8:28351708-28351 | - | inframe_deletion | MODERATE | ZNF395 | ENSG0000018691 | Transcript | ENST0000034442 | protein_coding | - | YES | MANE_Select |
| 9:33795614-33795 | TA | splice_donor_varia | HIGH | PRSS3 | ENSG0000001043 | Transcript | ENST0000037940 | protein_coding | rs1188045977 | YES | MANE_Select |

| High/Moderate Impact Variants | | | | | | | | | | | |
|---|---|---|---|---|---|---|---|---|---|---|---|
| **RS** | | | | | | | | | | | |
| **HNSC** | | | | | | | | | | | |
| **Location** | **Allele** | **Consequence** | **IMPACT** | **SYMBOL** | **Gene** | **Feature_type** | **Feature** | **BIOTYPE** | **Existing_variatio** | **CANONICAL** | **MANE** |
| 10:128106847-128 | - | frameshift_variant | HIGH | MKI67 | ENSG0000014877 | Transcript | ENST0000036865 | protein_coding | - | YES | MANE_Select |
| **BLGG** | | | | | | | | | | | |
| 1:214383705-2143 | - | inframe_deletion | MODERATE | PTPN14 | ENSG0000015210 | Transcript | ENST0000036695 | protein_coding | rs143136196 | YES | MANE_Select |
| 1:225847138-2258 | - | inframe_deletion | MODERATE | TMEM63A | ENSG0000019618 | Transcript | ENST0000036683 | protein_coding | rs750876984 | YES | MANE_Select |
| 10:128106847-128 | - | frameshift_variant | HIGH | MKI67 | ENSG0000014877 | Transcript | ENST0000036865 | protein_coding | rs145960091 | YES | MANE_Select |
| 16:50334766-5033 | - | frameshift_variant | HIGH | BRD7 | ENSG0000016616 | Transcript | ENST0000039468 | protein_coding | rs145896392 | YES | MANE_Select |
| 16:51141744-5114 | - | inframe_deletion | MODERATE | SALL1 | ENSG0000010344 | Transcript | ENST0000025102 | protein_coding | rs113614842 | YES | MANE_Select |
| 16:72788664-7278 | - | inframe_deletion | MODERATE | ZFHX3 | ENSG0000014083 | Transcript | ENST0000026848 | protein_coding | rs376311468 | YES | MANE_Select |
| 16:72797458-7279 | - | inframe_deletion | MODERATE | ZFHX3 | ENSG0000014083 | Transcript | ENST0000026848 | protein_coding | rs34918837 | YES | MANE_Select |
| 19:6531137-65311 | - | inframe_deletion | MODERATE | TNFSF9 | ENSG0000012565 | Transcript | ENST0000024581 | protein_coding | rs564151103 | YES | MANE_Select |
| 19:38408861-3840 | - | inframe_deletion | MODERATE | FAM98C | ENSG0000013024 | Transcript | ENST0000025253 | protein_coding | rs59917662 | YES | MANE_Select |
| 3:65439885-65439 | - | inframe_deletion | MODERATE | MAGI1 | ENSG0000015127 | Transcript | ENST0000040293 | protein_coding | rs142043619 | YES | MANE_Select |
| 4:53453080-53453 | - | frameshift_variant | HIGH | FIP1L1 | ENSG0000014521 | Transcript | ENST0000033748 | protein_coding | - | YES | MANE_Select |
| 6:161098318-1610 | - | inframe_deletion | MODERATE | MAP3K4 | ENSG0000008551 | Transcript | ENST0000039214 | protein_coding | rs5881391 | YES | MANE_Select |
| 8:28351708-28351 | - | inframe_deletion | MODERATE | ZNF395 | ENSG0000018691 | Transcript | ENST0000034442 | protein_coding | - | YES | MANE_Select |
| 9:130668313-1306 | - | splice_donor_varia | HIGH | PRDM12 | ENSG0000013071 | Transcript | ENST0000025300 | protein_coding | rs138789124 | YES | MANE_Select |
| **Adenocarcinoma** | | | | | | | | | | | |
| 1:6469122-646912 | - | inframe_deletion | MODERATE | PLEKHG5 | ENSG0000017168 | Transcript | ENST0000037772 | protein_coding | - | YES | MANE_Select |
| 10:30027571-3002 | - | inframe_deletion | MODERATE | JCAD | ENSG0000016575 | Transcript | ENST0000037537 | protein_coding | - | YES | MANE_Select |
| 11:115209591-115 | - | inframe_deletion | MODERATE | CADM1 | ENSG0000018298 | Transcript | ENST0000033158 | protein_coding | - | YES | MANE_Select |
| 12:8047961-80479 | - | inframe_deletion | MODERATE | FOXJ2 | ENSG0000006597 | Transcript | ENST0000016239 | protein_coding | - | YES | MANE_Select |
| 13:99982753-9998 | - | inframe_deletion | MODERATE | ZIC2 | ENSG0000004335 | Transcript | ENST0000037633 | protein_coding | - | YES | MANE_Select |
| 14:24163922-2416 | - | inframe_deletion | MODERATE | IRF9 | ENSG0000021392 | Transcript | ENST0000039686 | protein_coding | - | YES | MANE_Select |
| 14:24300643-2430 | - | inframe_deletion | MODERATE | NOP9 | ENSG0000019694 | Transcript | ENST0000026742 | protein_coding | - | YES | MANE_Select |
| 15:40036395-4003 | - | inframe_deletion | MODERATE | SRP14 | ENSG0000014031 | Transcript | ENST0000026788 | protein_coding | - | YES | MANE_Select |
| 16:50334766-5033 | - | frameshift_variant | HIGH | BRD7 | ENSG0000016616 | Transcript | ENST0000039468 | protein_coding | - | YES | MANE_Select |
| 16:72788664-7278 | - | inframe_deletion | MODERATE | ZFHX3 | ENSG0000014083 | Transcript | ENST0000026848 | protein_coding | rs376311468 | YES | MANE_Select |
| 19:1578372-15783 | - | inframe_deletion | MODERATE | MBD3 | ENSG0000007165 | Transcript | ENST0000043443 | protein_coding | - | YES | MANE_Select |
| 2:232378818-2323 | - | inframe_deletion | MODERATE | ALPP | ENSG0000016328 | Transcript | ENST0000039202 | protein_coding | - | YES | MANE_Select |
| 20:23365283-2336 | - | inframe_deletion | MODERATE | GZF1 | ENSG0000012581 | Transcript | ENST0000033812 | protein_coding | - | YES | MANE_Select |
| 20:34743163-3474 | - | inframe_deletion | MODERATE | NCOA6 | ENSG0000019864 | Transcript | ENST0000035900 | protein_coding | - | YES | MANE_Select |
| 20:47238822-4723 | - | inframe_deletion | MODERATE | ZMYND8 | ENSG0000010104 | Transcript | ENST0000047195 | protein_coding | rs569370037 | YES | MANE_Select |
| 22:42214566-4221 | - | inframe_deletion | MODERATE | TCF20 | ENSG0000010020 | Transcript | ENST0000067762 | protein_coding | - | YES | MANE_Select |

| | | | | | | | | | | | |
|---|---|---|---|---|---|---|---|---|---|---|---|
| 3:65439885-65439 | - | inframe_deletion | MODERATE | MAGI1 | ENSG0000015127 | Transcript | ENST0000040293 | protein_coding | - | YES | MANE_Select |
| 4:53453080-53453 | - | frameshift_variant | HIGH | FIP1L1 | ENSG0000014521 | Transcript | ENST0000033748 | protein_coding | - | YES | MANE_Select |
| 8:28351708-28351 | - | inframe_deletion | MODERATE | ZNF395 | ENSG0000018691 | Transcript | ENST0000034442 | protein_coding | rs547945591 | YES | MANE_Select |

| | | | | | | | | | | | |
|---|---|---|---|---|---|---|---|---|---|---|---|
| **Low/Modifier Impact Variants** | | | | | | | | | | | |
| **RR** | | | | | | | | | | | |
| **Squamous** | | | | | | | | | | | |
| **Location** | **Allele** | **Consequence** | **IMPACT** | **SYMBOL** | **Gene** | **Feature_type** | **Feature** | **BIOTYPE** | **Existing_variatio** | **CANONICAL** | **MANE** |
| 2:37372476-37372480 | - | splice_region_vari | LOW | QPCT | ENSG0000011582 | Transcript | ENST0000033841 | protein_coding | - | YES | MANE_Select |
| 2:37372476-37372480 | - | downstream_gene_ | MODIFIER | - | ENSG0000029646 | Transcript | ENST0000073982 | lncRNA | - | YES | - |
| 6:151458285-151458289 | - | intron_variant | MODIFIER | ARMT1 | ENSG0000014647 | Transcript | ENST0000036729 | protein_coding | - | YES | MANE_Select |
| **Carcinoma** | | | | | | | | | | | |
| 12:56417805-56417809 | - | intron_variant | MODIFIER | TIMELESS | ENSG0000011160 | Transcript | ENST0000055353 | protein_coding | - | YES | MANE_Select |
| 12:56417805-56417809 | - | upstream_gene_va | MODIFIER | - | ENSG0000027627 | Transcript | ENST0000061044 | lncRNA | - | YES | - |
| 12:56417805-56417809 | - | upstream_gene_va | MODIFIER | - | ENSG0000027456 | Transcript | ENST0000062058 | unitary_pseudogen | - | YES | - |
| 2:222293705-222293709 | - | intron_variant | MODIFIER | PAX3 | ENSG0000013590 | Transcript | ENST0000039207 | protein_coding | - | YES | MANE_Select |
| 2:222293705-222293709 | - | upstream_gene_va | MODIFIER | CCDC140 | ENSG0000016308 | Transcript | ENST0000064776 | lncRNA | - | YES | - |
| **Adenocarcinoma** | | | | | | | | | | | |
| 1:6469122-6469131 | - | upstream_gene_va | MODIFIER | TNFRSF25 | ENSG0000021578 | Transcript | ENST0000035687 | protein_coding | - | YES | MANE_Select |
| 1:17027878-17027884 | - | splice_polypyrimic | LOW | SDHB | ENSG0000011711 | Transcript | ENST0000037549 | protein_coding | - | YES | MANE_Select |
| 1:210684141-210684145 | - | splice_region_vari | LOW | KCNH1 | ENSG0000014347 | Transcript | ENST0000027175 | protein_coding | - | YES | MANE_Select |
| 1:210684141-210684145 | - | downstream_gene_ | MODIFIER | - | ENSG0000027933 | Transcript | ENST0000062514 | TEC | - | YES | - |
| 10:112404668-112404672 | - | intron_variant | MODIFIER | ACSL5 | ENSG0000019714 | Transcript | ENST0000035465 | protein_coding | - | YES | MANE_Select |
| 10:112404668-112404672 | - | downstream_gene_ | MODIFIER | - | ENSG0000023293 | Transcript | ENST0000063108 | lncRNA | - | YES | - |
| 10:113848892-113848896 | - | intron_variant | MODIFIER | DCLRE1A | ENSG0000019892 | Transcript | ENST0000036138 | protein_coding | - | YES | MANE_Select |
| 10:113848892-113848896 | - | intron_variant,non | MODIFIER | - | ENSG0000028893 | Transcript | ENST0000069264 | lncRNA | - | YES | - |
| 11:75797346-75797350 | - | intron_variant | MODIFIER | DGAT2 | ENSG0000006228 | Transcript | ENST0000022802 | protein_coding | - | YES | MANE_Select |
| 11:75797346-75797350 | - | intron_variant,non | MODIFIER | UVRAG-DT | ENSG0000029362 | Transcript | ENST0000071620 | lncRNA | - | YES | - |
| 12:95898502-95898510 | - | intron_variant | MODIFIER | CCDC38 | ENSG0000016597 | Transcript | ENST0000034428 | protein_coding | - | YES | MANE_Select |
| 12:113947257-113947261 | - | intron_variant | MODIFIER | RBM19 | ENSG0000012296 | Transcript | ENST0000026174 | protein_coding | - | YES | MANE_Select |
| 13:99982753-99982759 | - | upstream_gene_va | MODIFIER | - | ENSG0000028806 | Transcript | ENST0000065377 | lncRNA | - | YES | - |
| 13:101090060-101090064 | - | intron_variant | MODIFIER | NALCN | ENSG0000010245 | Transcript | ENST0000025112 | protein_coding | - | YES | MANE_Select |
| 15:73288149-73288155 | - | intron_variant | MODIFIER | NEO1 | ENSG0000006714 | Transcript | ENST0000026190 | protein_coding | - | YES | MANE_Select |
| 16:72788664-72788670 | - | intron_variant,non | MODIFIER | ZFHX3-AS1 | ENSG0000025976 | Transcript | ENST0000076645 | lncRNA | - | YES | - |
| 16:72797458-72797464 | - | intron_variant,non | MODIFIER | ZFHX3-AS1 | ENSG0000025976 | Transcript | ENST0000076645 | lncRNA | - | YES | - |
| 17:4809268-4809272 | - | intron_variant | MODIFIER | PLD2 | ENSG0000012921 | Transcript | ENST0000026308 | protein_coding | - | YES | MANE_Select |
| 17:28584629-28584633 | - | intron_variant | MODIFIER | SPAG5 | ENSG0000007638 | Transcript | ENST0000032176 | protein_coding | - | YES | MANE_Select |
| 17:39745549-39745553 | - | intron_variant | MODIFIER | GRB7 | ENSG0000014173 | Transcript | ENST0000030915 | protein_coding | - | YES | MANE_Select |
| 19:2733503-2733507 | - | intron_variant | MODIFIER | SLC39A3 | ENSG0000014187 | Transcript | ENST0000026974 | protein_coding | - | YES | MANE_Select |
| 19:2733503-2733507 | - | upstream_gene_va | MODIFIER | - | ENSG0000026134 | Transcript | ENST0000056790 | lncRNA | - | YES | - |
| 19:58557202-58557206 | - | downstream_gene_ | MODIFIER | MZF1 | ENSG0000009932 | Transcript | ENST0000021505 | protein_coding | - | YES | MANE_Select |
| 19:58557202-58557206 | - | intron_variant | MODIFIER | UBE2M | ENSG0000013072 | Transcript | ENST0000025302 | protein_coding | - | YES | MANE_Select |
| 19:58557202-58557206 | - | upstream_gene_va | MODIFIER | CHMP2A | ENSG0000013072 | Transcript | ENST0000031254 | protein_coding | - | YES | MANE_Select |
| 19:58557202-58557206 | - | upstream_gene_va | MODIFIER | MZF1-AS1 | ENSG0000026785 | Transcript | ENST0000078983 | lncRNA | - | YES | - |
| 2:38300255-38300259 | - | intron_variant | MODIFIER | ATL2 | ENSG0000011978 | Transcript | ENST0000037895 | protein_coding | - | YES | MANE_Select |
| 2:233769464-233769468 | - | intron_variant | MODIFIER | UGT1A6 | ENSG0000016716 | Transcript | ENST0000030513 | protein_coding | - | YES | MANE_Select |

| | | | | | | | | | | | |
|---|---|---|---|---|---|---|---|---|---|---|---|
| 2:233769464-233769468 | - | intron_variant | MODIFIER | UGT1A1 | ENSG0000024163 | Transcript | ENST0000030520 | protein_coding | - | YES | MANE_Select |
| 2:233769464-233769468 | - | intron_variant | MODIFIER | UGT1A10 | ENSG0000024251 | Transcript | ENST0000034464 | protein_coding | - | YES | MANE_Select |
| 2:233769464-233769468 | - | intron_variant | MODIFIER | UGT1A9 | ENSG0000024111 | Transcript | ENST0000035472 | protein_coding | - | YES | MANE_Select |
| 2:233769464-233769468 | - | intron_variant | MODIFIER | UGT1A4 | ENSG0000024447 | Transcript | ENST0000037340 | protein_coding | - | YES | MANE_Select |
| 2:233769464-233769468 | - | intron_variant | MODIFIER | UGT1A5 | ENSG0000028870 | Transcript | ENST0000037341 | protein_coding | - | YES | MANE_Select |
| 2:233769464-233769468 | - | intron_variant | MODIFIER | UGT1A7 | ENSG0000024412 | Transcript | ENST0000037342 | protein_coding | - | YES | MANE_Select |
| 2:233769464-233769468 | - | intron_variant | MODIFIER | UGT1A8 | ENSG0000024236 | Transcript | ENST0000037345 | protein_coding | - | YES | MANE_Select |
| 2:233769464-233769468 | - | intron_variant | MODIFIER | UGT1A3 | ENSG0000028870 | Transcript | ENST0000048202 | protein_coding | - | YES | MANE_Select |
| 2:233769464-233769468 | - | downstream_gene_ | MODIFIER | - | ENSG0000029847 | Transcript | ENST0000075573 | lncRNA | - | YES | - |
| 20:31472917-31472921 | - | upstream_gene_va | MODIFIER | REM1 | ENSG0000008832 | Transcript | ENST0000020197 | protein_coding | - | YES | MANE_Select |
| 20:31472917-31472921 | - | intron_variant | MODIFIER | DEFB124 | ENSG0000018038 | Transcript | ENST0000031767 | protein_coding | - | YES | MANE_Select |
| 20:47169101-47169105 | - | intron_variant | MODIFIER | EYA2 | ENSG0000006465 | Transcript | ENST0000032761 | protein_coding | - | YES | MANE_Select |
| 20:47169101-47169105 | - | downstream_gene_ | MODIFIER | MIR3616 | ENSG0000026490 | Transcript | ENST0000058407 | miRNA | - | YES | - |
| 20:49011831-49011835 | - | intron_variant | MODIFIER | ARFGEF2 | ENSG0000012419 | Transcript | ENST0000037191 | protein_coding | - | YES | MANE_Select |
| 20:56248628-56248632 | - | upstream_gene_va | MODIFIER | MC3R | ENSG0000012408 | Transcript | ENST0000024391 | protein_coding | - | YES | MANE_Select |
| 21:34708146-34708150 | - | intron_variant | MODIFIER | CLIC6 | ENSG0000015921 | Transcript | ENST0000034949 | protein_coding | - | YES | MANE_Select |
| 21:45917570-45917574 | - | splice_polypyrimic | LOW | - | ENSG0000028060 | Transcript | ENST0000062623 | lncRNA | - | YES | - |
| 21:45917570-45917574 | - | splice_polypyrimic | LOW | PCBP3 | ENSG0000018357 | Transcript | ENST0000068168 | protein_coding | - | YES | MANE_Select |
| 3:58134591-58134595 | - | intron_variant | MODIFIER | FLNB | ENSG0000013606 | Transcript | ENST0000029595 | protein_coding | - | YES | MANE_Select |
| 4:654753-654757 | - | intron_variant,non_ | MODIFIER | PDE6B-AS1 | ENSG0000024268 | Transcript | ENST0000046835 | lncRNA | - | YES | - |
| 4:654753-654757 | - | intron_variant | MODIFIER | PDE6B | ENSG0000013325 | Transcript | ENST0000049651 | protein_coding | - | YES | MANE_Select |
| 7:123689006-123689020 | - | intron_variant | MODIFIER | WASL | ENSG0000010629 | Transcript | ENST0000022302 | protein_coding | - | YES | MANE_Select |
| 9:18474375-18474379 | - | intron_variant | MODIFIER | ADAMTSL1 | ENSG0000017803 | Transcript | ENST0000038054 | protein_coding | - | YES | MANE_Select |
| 9:36923306-36923310 | - | intron_variant | MODIFIER | PAX5 | ENSG0000019609 | Transcript | ENST0000035812 | protein_coding | - | YES | MANE_Select |
| **PRAD** | | | | | | | | | | | |
| **BLGG** | | | | | | | | | | | |
| 1:20857326-20857330 | - | intron_variant | MODIFIER | EIF4G3 | ENSG0000007515 | Transcript | ENST0000060232 | protein_coding | - | YES | MANE_Select |
| 1:225847138-225847144 | - | downstream_gene_ | MODIFIER | EPHX1 | ENSG0000014381 | Transcript | ENST0000027216 | protein_coding | - | YES | MANE_Select |
| 1:225847138-225847144 | - | upstream_gene_va | MODIFIER | - | ENSG0000024286 | Transcript | ENST0000042433 | lncRNA | - | YES | - |
| 17:32997377-32997381 | - | intron_variant | MODIFIER | SPACA3 | ENSG0000014131 | Transcript | ENST0000026905 | protein_coding | - | YES | MANE_Select |
| 19:7678296-7678300 | - | intron_variant | MODIFIER | MCEMP1 | ENSG0000018301 | Transcript | ENST0000033359 | protein_coding | - | YES | MANE_Select |
| 19:7678296-7678300 | - | upstream_gene_va | MODIFIER | TRAPPC5 | ENSG0000018102 | Transcript | ENST0000059614 | protein_coding | - | YES | MANE_Select |
| 19:19833449-19833453 | - | intron_variant,non_ | MODIFIER | ZNF56P | ENSG0000029117 | Transcript | ENST0000059188 | lncRNA | - | YES | - |
| 19:19833449-19833453 | - | upstream_gene_va | MODIFIER | ZNF56P | ENSG0000026741 | Transcript | ENST0000062341 | transcribed_unproc | - | YES | - |
| 19:19833449-19833453 | - | intron_variant,non_ | MODIFIER | - | ENSG0000029615 | Transcript | ENST0000073692 | lncRNA | - | YES | - |
| 19:38408861-38408867 | - | downstream_gene_ | MODIFIER | RASGRP4 | ENSG0000017177 | Transcript | ENST0000061543 | protein_coding | - | YES | MANE_Select |
| 20:56248628-56248632 | - | upstream_gene_va | MODIFIER | MC3R | ENSG0000012408 | Transcript | ENST0000024391 | protein_coding | - | YES | MANE_Select |
| 6:133889359-133889363 | - | 5_prime_UTR_var | MODIFIER | TCF21 | ENSG0000011852 | Transcript | ENST0000036788 | protein_coding | - | YES | MANE_Select |
| 6:133889359-133889363 | - | upstream_gene_va | MODIFIER | TARID | ENSG0000022795 | Transcript | ENST0000079540 | lncRNA | - | YES | - |
| 6:151458285-151458289 | - | intron_variant | MODIFIER | ARMT1 | ENSG0000014647 | Transcript | ENST0000036729 | protein_coding | - | YES | MANE_Select |
| 7:2945755-2945759 | - | intron_variant | MODIFIER | CARD11 | ENSG0000019828 | Transcript | ENST0000039694 | protein_coding | - | YES | MANE_Select |
| 7:2945755-2945759 | - | intron_variant,non_ | MODIFIER | CARD11-AS1 | ENSG0000023728 | Transcript | ENST0000081660 | lncRNA | - | YES | - |

| | | | | | | | | | | | |
|---|---|---|---|---|---|---|---|---|---|---|---|
| 7:130041131-130041135 | - | intron_variant | MODIFIER | ZC3HC1 | ENSG0000009173 | Transcript | ENST0000035830 | protein_coding | - | YES | MANE_Select |
| 9:35825544-35825550 | - | intron_variant | MODIFIER | FAM221B | ENSG0000020493 | Transcript | ENST0000042353 | protein_coding | - | YES | MANE_Select |
| 9:35825544-35825550 | - | upstream_gene_va | MODIFIER | TMEM8B | ENSG0000013710 | Transcript | ENST0000064393 | protein_coding | - | YES | MANE_Select |
| 9:130668313-130668317 | - | splice_donor_regio | LOW | PRDM12 | ENSG0000013071 | Transcript | ENST0000025300 | protein_coding | - | YES | MANE_Select |
| **HNSC** | | | | | | | | | | | |
| 1:20857326-20857330 | - | intron_variant | MODIFIER | EIF4G3 | ENSG0000007515 | Transcript | ENST0000060232 | protein_coding | - | YES | MANE_Select |
| 6:132463932-132463936 | - | intron_variant | MODIFIER | STX7 | ENSG0000007995 | Transcript | ENST0000036794 | protein_coding | - | YES | MANE_Select |
| **BRCA** | | | | | | | | | | | |
| 12:49656368-49656372 | - | intron_variant | MODIFIER | FMNL3 | ENSG0000016179 | Transcript | ENST0000033515 | protein_coding | - | YES | MANE_Select |

| | | | | | | | | | | | |
|---|---|---|---|---|---|---|---|---|---|---|---|
| **Low/Modifier Impact Variants** | | | | | | | | | | | |
| **RS** | | | | | | | | | | | |
| **Squamous** | | | | | | | | | | | |
| **Location** | **Allele** | **Consequence** | **IMPACT** | **SYMBOL** | **Gene** | **Feature_type** | **Feature** | **BIOTYPE** | **Existing_variatio** | **CANONICAL** | **MANE** |
| 2:37372476-37372480 | - | splice_region_vari | LOW | QPCT | ENSG0000011582 | Transcript | ENST0000033841 | protein_coding | - | YES | MANE_Select |
| 2:37372476-37372480 | - | downstream_gene_ | MODIFIER | - | ENSG0000029646 | Transcript | ENST0000073982 | lncRNA | - | YES | - |
| 6:151458285-151458289 | - | intron_variant | MODIFIER | ARMT1 | ENSG0000014647 | Transcript | ENST0000036729 | protein_coding | - | YES | MANE_Select |
| **Adenocarcinoma** | | | | | | | | | | | |
| 1:6469122-6469128 | - | upstream_gene_va | MODIFIER | TNFRSF25 | ENSG0000021578 | Transcript | ENST0000035687 | protein_coding | - | YES | MANE_Select |
| 1:20857326-20857330 | - | intron_variant | MODIFIER | EIF4G3 | ENSG0000007515 | Transcript | ENST0000060232 | protein_coding | - | YES | MANE_Select |
| 1:25227386-25227390 | - | intron_variant | MODIFIER | SYF2 | ENSG0000011761 | Transcript | ENST0000023627 | protein_coding | - | YES | MANE_Select |
| 10:30027571-30027577 | - | non_coding_transc | MODIFIER | - | ENSG0000030452 | Transcript | ENST0000080435 | lncRNA | - | YES | - |
| 10:50743043-50743047 | - | intron_variant | MODIFIER | ASAH2B | ENSG0000020414 | Transcript | ENST0000064731 | protein_coding | - | YES | MANE_Select |
| 10:50743043-50743047 | - | upstream_gene_va | MODIFIER | - | ENSG0000028722 | Transcript | ENST0000067139 | lncRNA | - | YES | - |
| 10:94358221-94358225 | - | splice_polypyrimi | LOW | NOC3L | ENSG0000017314 | Transcript | ENST0000037136 | protein_coding | - | YES | MANE_Select |
| 11:90071124-90071128 | - | downstream_gene_ | MODIFIER | - | ENSG0000025488 | Transcript | ENST0000052781 | processed_pseudo | - | YES | - |
| 11:90071124-90071128 | - | upstream_gene_va | MODIFIER | - | ENSG0000025501 | Transcript | ENST0000053203 | unprocessed_pseu | - | YES | - |
| 12:8868422-8868426 | - | intron_variant | MODIFIER | A2ML1 | ENSG0000016653 | Transcript | ENST0000029969 | protein_coding | - | YES | MANE_Select |
| 12:8868422-8868426 | - | intron_variant,non | MODIFIER | - | ENSG0000028202 | Transcript | ENST0000063183 | lncRNA | - | YES | - |
| 12:56417805-56417809 | - | intron_variant | MODIFIER | TIMELESS | ENSG0000011160 | Transcript | ENST0000055353 | protein_coding | - | YES | MANE_Select |
| 12:56417805-56417809 | - | upstream_gene_va | MODIFIER | - | ENSG0000027627 | Transcript | ENST0000061044 | lncRNA | - | YES | - |
| 12:56417805-56417809 | - | upstream_gene_va | MODIFIER | - | ENSG0000027456 | Transcript | ENST0000062058 | unitary_pseudogen | - | YES | - |
| 12:89598780-89598784 | - | intron_variant | MODIFIER | ATP2B1 | ENSG0000007096 | Transcript | ENST0000042867 | protein_coding | - | YES | MANE_Select |
| 13:77070573-77070577 | - | intron_variant | MODIFIER | MYCBP2 | ENSG0000000581 | Transcript | ENST0000054444 | protein_coding | - | YES | MANE_Select |
| 13:99982753-99982768 | - | upstream_gene_va | MODIFIER | - | ENSG0000028806 | Transcript | ENST0000065377 | lncRNA | - | YES | - |
| 14:24163922-24163928 | - | downstream_gene_ | MODIFIER | RNF31 | ENSG0000009209 | Transcript | ENST0000032410 | protein_coding | - | YES | MANE_Select |
| 14:24163922-24163928 | - | downstream_gene_ | MODIFIER | - | ENSG0000028975 | Transcript | ENST0000069902 | lncRNA | - | YES | - |
| 14:24300643-24300649 | - | upstream_gene_va | MODIFIER | DHRS1 | ENSG0000015737 | Transcript | ENST0000028811 | protein_coding | - | YES | MANE_Select |
| 14:24300643-24300649 | - | downstream_gene_ | MODIFIER | CIDEB | ENSG0000013630 | Transcript | ENST0000055441 | protein_coding | - | YES | MANE_Select |
| 14:24300643-24300649 | - | downstream_gene_ | MODIFIER | - | ENSG0000028804 | Transcript | ENST0000073790 | lncRNA | - | YES | - |
| 14:61457374-61457378 | - | intron_variant | MODIFIER | PRKCH | ENSG0000002707 | Transcript | ENST0000033298 | protein_coding | - | YES | MANE_Select |
| 15:40036395-40036401 | - | downstream_gene_ | MODIFIER | EIF2AK4 | ENSG0000012882 | Transcript | ENST0000026379 | protein_coding | - | YES | MANE_Select |
| 15:40036395-40036401 | - | upstream_gene_va | MODIFIER | SRP14-DT | ENSG0000024850 | Transcript | ENST0000074667 | lncRNA | - | YES | - |
| 16:3284207-3284211 | - | splice_donor_5th_ | LOW | ZNF263 | ENSG0000000619 | Transcript | ENST0000021906 | protein_coding | - | YES | MANE_Select |
| 17:8160737-8160741 | - | 3_prime_UTR_vai | MODIFIER | VAMP2 | ENSG0000022020 | Transcript | ENST0000031650 | protein_coding | - | YES | MANE_Select |
| 17:8160737-8160741 | - | upstream_gene_va | MODIFIER | - | ENSG0000029995 | Transcript | ENST0000076770 | lncRNA | - | YES | - |
| 17:42955992-42955996 | - | intron_variant | MODIFIER | AARSD1 | ENSG0000026696 | Transcript | ENST0000042756 | protein_coding | - | YES | MANE_Select |
| 17:60069867-60069871 | - | intron_variant | MODIFIER | HEATR6 | ENSG0000006809 | Transcript | ENST0000018495 | protein_coding | - | YES | MANE_Select |
| 19:1578372-1578378 | - | downstream_gene_ | MODIFIER | - | ENSG0000027900 | Transcript | ENST0000062442 | TEC | - | YES | - |
| 19:20132594-20132598 | - | splice_polypyrimi | LOW | - | ENSG0000026722 | Transcript | ENST0000059292 | transcribed_unpro | - | YES | - |
| 19:20132594-20132598 | - | splice_polypyrimi | LOW | - | ENSG0000026738 | Transcript | ENST0000059365 | lncRNA | - | YES | - |

| | | | | | | | | | | | |
|---|---|---|---|---|---|---|---|---|---|---|---|
| 19:20132594-20132598 | - | upstream_gene_va | MODIFIER | BNIP3P15 | ENSG0000027071 | Transcript | ENST0000060460 | processed_pseudog | - | YES | - |
| 19:42739195-42739199 | - | intron_variant | MODIFIER | PSG3 | ENSG0000022182 | Transcript | ENST0000032749 | protein_coding | - | YES | MANE_Select |
| 19:48836425-48836429 | - | downstream_gene_ | MODIFIER | PLEKHA4 | ENSG0000010555 | Transcript | ENST0000026326 | protein_coding | - | YES | MANE_Select |
| 19:48836425-48836429 | - | 5_prime_UTR_var | MODIFIER | HSD17B14 | ENSG0000008707 | Transcript | ENST0000026327 | protein_coding | - | YES | MANE_Select |
| 2:37372476-37372480 | - | splice_region_vari | LOW | QPCT | ENSG0000011582 | Transcript | ENST0000033841 | protein_coding | - | YES | MANE_Select |
| 2:37372476-37372480 | - | downstream_gene_ | MODIFIER | - | ENSG0000029646 | Transcript | ENST0000073982 | lncRNA | - | YES | - |
| 2:222293705-222293709 | - | intron_variant | MODIFIER | PAX3 | ENSG0000013590 | Transcript | ENST0000039207 | protein_coding | - | YES | MANE_Select |
| 2:222293705-222293709 | - | upstream_gene_va | MODIFIER | CCDC140 | ENSG0000016308 | Transcript | ENST0000064776 | lncRNA | - | YES | - |
| 2:232378818-232378824 | - | intron_variant,non | MODIFIER | ECEL1P2 | ENSG0000022451 | Transcript | ENST0000071529 | lncRNA | - | YES | - |
| 20:34743163-34743169 | - | intron_variant,non | MODIFIER | - | ENSG0000028680 | Transcript | ENST0000065504 | lncRNA | - | YES | - |
| 20:47169101-47169107 | - | intron_variant | MODIFIER | EYA2 | ENSG0000006465 | Transcript | ENST0000032761 | protein_coding | - | YES | MANE_Select |
| 20:47169101-47169107 | - | downstream_gene_ | MODIFIER | MIR3616 | ENSG0000026490 | Transcript | ENST0000058407 | miRNA | - | YES | - |
| 3:65375708-65375712 | - | intron_variant | MODIFIER | MAGI1 | ENSG0000015127 | Transcript | ENST0000040293 | protein_coding | - | YES | MANE_Select |
| 3:65387174-65387178 | - | intron_variant | MODIFIER | MAGI1 | ENSG0000015127 | Transcript | ENST0000040293 | protein_coding | - | YES | MANE_Select |
| 3:128573475-128573479 | - | intron_variant,non | MODIFIER | LINC01565 | ENSG0000019868 | Transcript | ENST0000075324 | lncRNA | - | YES | - |
| 5:170080126-170080130 | - | intron_variant | MODIFIER | DOCK2 | ENSG0000013451 | Transcript | ENST0000052090 | protein_coding | - | YES | MANE_Select |
| 6:132463932-132463936 | - | intron_variant | MODIFIER | STX7 | ENSG0000007995 | Transcript | ENST0000036794 | protein_coding | - | YES | MANE_Select |
| 7:20140884-20140888 | - | 3_prime_UTR_var | MODIFIER | MACC1 | ENSG0000018374 | Transcript | ENST0000040033 | protein_coding | - | YES | MANE_Select |
| 7:20140884-20140888 | - | intron_variant,non | MODIFIER | MACC1-AS1 | ENSG0000022859 | Transcript | ENST0000073867 | lncRNA | - | YES | - |
| 7:29504721-29504725 | - | intron_variant | MODIFIER | CHN2 | ENSG0000010606 | Transcript | ENST0000022279 | protein_coding | - | YES | MANE_Select |
| 7:38245284-38245288 | - | intron_variant | MODIFIER | TRGC2 | ENSG0000022719 | Transcript | ENST0000043691 | TR_C_gene | - | YES | - |
| 7:47413908-47413914 | - | intron_variant | MODIFIER | TNS3 | ENSG0000013620 | Transcript | ENST0000031116 | protein_coding | - | YES | MANE_Select |
| 7:96028144-96028148 | - | intron_variant | MODIFIER | DYNC1I1 | ENSG0000015856 | Transcript | ENST0000044746 | protein_coding | - | YES | MANE_Select |
| 7:142474204-142474208 | - | downstream_gene_ | MODIFIER | TRBV6-6 | ENSG0000021172 | Transcript | ENST0000039037 | TR_V_gene | - | YES | - |
| 7:142474204-142474208 | - | intron_variant,non | MODIFIER | TRBV7-5 | ENSG0000022755 | Transcript | ENST0000039038 | TR_V_pseudogene | - | YES | - |
| 8:67161738-67161742 | - | intron_variant | MODIFIER | CSPP1 | ENSG0000010421 | Transcript | ENST0000067861 | protein_coding | - | YES | MANE_Select |
| 8:140532162-140532166 | - | splice_polypyrimic | LOW | AGO2 | ENSG0000012390 | Transcript | ENST0000022059 | protein_coding | - | YES | MANE_Select |
| 8:144317162-144317166 | - | upstream_gene_va | MODIFIER | - | ENSG0000025469 | Transcript | ENST0000052502 | lncRNA | - | YES | - |
| 8:144317162-144317166 | - | intron_variant | MODIFIER | DGAT1 | ENSG0000018500 | Transcript | ENST0000052871 | protein_coding | - | YES | MANE_Select |
| 8:144317162-144317166 | - | downstream_gene_ | MODIFIER | HSF1 | ENSG0000018512 | Transcript | ENST0000052883 | protein_coding | - | YES | MANE_Select |
| 8:144317162-144317166 | - | downstream_gene_ | MODIFIER | MIR6848 | ENSG0000028422 | Transcript | ENST0000061121 | miRNA | - | YES | - |
| 9:35825544-35825550 | - | intron_variant | MODIFIER | FAM221B | ENSG0000020493 | Transcript | ENST0000042353 | protein_coding | - | YES | MANE_Select |
| 9:35825544-35825550 | - | upstream_gene_va | MODIFIER | TMEM8B | ENSG0000013710 | Transcript | ENST0000064393 | protein_coding | - | YES | MANE_Select |
| X:139958016-139958020 | - | intron_variant | MODIFIER | CXorf66 | ENSG0000020393 | Transcript | ENST0000037054 | protein_coding | - | YES | MANE_Select |
| **PAAD** | | | | | | | | | | | |
| **BLGG** | | | | | | | | | | | |
| 19:20184331-20184335 | - | intron_variant | MODIFIER | ZNF486 | ENSG0000025622 | Transcript | ENST0000033511 | protein_coding | - | YES | MANE_Select |
| 19:20184331-20184335 | - | intron_variant,non | MODIFIER | - | ENSG0000026738 | Transcript | ENST0000059365 | lncRNA | - | YES | - |
| 2:11133310-11133314 | - | 5_prime_UTR_var | MODIFIER | CIMIP5 | ENSG0000015087 | Transcript | ENST0000038158 | protein_coding | - | YES | MANE_Select |
| 2:11133310-11133314 | - | upstream_gene_va | MODIFIER | - | ENSG0000014506 | Transcript | ENST0000054430 | lncRNA | - | YES | - |
| 2:11133310-11133314 | - | downstream_gene_ | MODIFIER | - | ENSG0000029616 | Transcript | ENST0000073702 | lncRNA | - | YES | - |

| | | | | | | | | | | | |
|---|---|---|---|---|---|---|---|---|---|---|---|
| 3:187253326-187253330 | - | splice_polypyrimic | LOW | MASP1 | ENSG0000012724 | Transcript | ENST0000029628 | protein_coding | - | YES | MANE_Select |
| 4:654753-654757 | - | intron_variant,non_ | MODIFIER | PDE6B-AS1 | ENSG0000024268 | Transcript | ENST0000046835 | lncRNA | - | YES | - |
| 4:654753-654757 | - | intron_variant | MODIFIER | PDE6B | ENSG0000013325 | Transcript | ENST0000049651 | protein_coding | - | YES | MANE_Select |
| 6:144700057-144700063 | - | intron_variant | MODIFIER | UTRN | ENSG0000015281 | Transcript | ENST0000036754 | protein_coding | - | YES | MANE_Select |
| **LUAD** | | | | | | | | | | | |
| **HNSC** | | | | | | | | | | | |
| 19:20646412-20646416 | - | splice_polypyrimic | LOW | ZNF626 | ENSG0000018817 | Transcript | ENST0000060144 | protein_coding | - | YES | MANE_Select |

| Low/Modifier Impact Variants | | | | | | | | | | | |
|---|---|---|---|---|---|---|---|---|---|---|---|
| **RR** | | | | | | | | | | | |
| **Squamous** | | | | | | | | | | | |
| **Location** | **Allele** | **Consequence** | **IMPACT** | **SYMBOL** | **Gene** | **Feature_type** | **Feature** | **BIOTYPE** | **Existing_variatio** | **CANONICAL** | **MANE** |
| 1:20857326-20857 | - | intron_variant | MODIFIER | EIF4G3 | ENSG0000007515 | Transcript | ENST0000060232 | protein_coding | rs145504996 | YES | MANE_Select |
| 11:26671047-2667 | - | 3_prime_UTR_va | MODIFIER | SLC5A12 | ENSG0000014894 | Transcript | ENST0000039600 | protein_coding | rs71449136 | YES | MANE_Select |
| 17:17819267-1781 | - | intron_variant | MODIFIER | SREBF1 | ENSG0000007231 | Transcript | ENST0000026164 | protein_coding | rs72248634 | YES | MANE_Select |
| 19:20184331-2018 | - | intron_variant | MODIFIER | ZNF486 | ENSG0000025622 | Transcript | ENST0000033511 | protein_coding | rs367731486 | YES | MANE_Select |
| 19:20184331-2018 | - | intron_variant,non | MODIFIER | - | ENSG0000026738 | Transcript | ENST0000059365 | lncRNA | rs367731486 | YES | - |
| 2:37372476-37372 | - | splice_region_vari | LOW | QPCT | ENSG0000011582 | Transcript | ENST0000033841 | protein_coding | - | YES | MANE_Select |
| 2:37372476-37372 | - | downstream_gene_ | MODIFIER | - | ENSG0000029646 | Transcript | ENST0000073982 | lncRNA | - | YES | - |
| 6:151458285-1514 | - | intron_variant | MODIFIER | ARMT1 | ENSG0000014647 | Transcript | ENST0000036729 | protein_coding | - | YES | MANE_Select |
| **Carcinoma** | | | | | | | | | | | |
| 12:56417805-5641 | - | intron_variant | MODIFIER | TIMELESS | ENSG0000011160 | Transcript | ENST0000055353 | protein_coding | - | YES | MANE_Select |
| 12:56417805-5641 | - | upstream_gene_va | MODIFIER | - | ENSG0000027627 | Transcript | ENST0000061044 | lncRNA | - | YES | - |
| 12:56417805-5641 | - | upstream_gene_va | MODIFIER | - | ENSG0000027456 | Transcript | ENST0000062058 | unitary_pseudogen | - | YES | - |
| 2:222293705-2222 | - | intron_variant | MODIFIER | PAX3 | ENSG0000013590 | Transcript | ENST0000039207 | protein_coding | - | YES | MANE_Select |
| 2:222293705-2222 | - | upstream_gene_va | MODIFIER | CCDC140 | ENSG0000016308 | Transcript | ENST0000064776 | lncRNA | - | YES | - |
| **Adenocarcinoma** | | | | | | | | | | | |
| 1:6469122-646913 | - | upstream_gene_va | MODIFIER | TNFRSF25 | ENSG0000021578 | Transcript | ENST0000035687 | protein_coding | - | YES | MANE_Select |
| 1:17027878-17027 | - | splice_polypyrimi | LOW | SDHB | ENSG0000011711 | Transcript | ENST0000037549 | protein_coding | - | YES | MANE_Select |
| 1:20857326-20857 | - | intron_variant | MODIFIER | EIF4G3 | ENSG0000007515 | Transcript | ENST0000060232 | protein_coding | rs145504996 | YES | MANE_Select |
| 1:25227386-25227 | - | intron_variant | MODIFIER | SYF2 | ENSG0000011761 | Transcript | ENST0000023627 | protein_coding | rs111662912 | YES | MANE_Select |
| 1:210684141-2106 | - | splice_region_vari | LOW | KCNH1 | ENSG0000014347 | Transcript | ENST0000027175 | protein_coding | - | YES | MANE_Select |
| 1:210684141-2106 | - | downstream_gene_ | MODIFIER | - | ENSG0000027933 | Transcript | ENST0000062514 | TEC | - | YES | - |
| 10:30027571-3002 | - | non_coding_transc | MODIFIER | - | ENSG0000030452 | Transcript | ENST0000080435 | lncRNA | rs34594193 | YES | - |
| 10:36522778-3652 | - | non_coding_transc | MODIFIER | NAMPTP1 | ENSG0000022964 | Transcript | ENST0000044046 | processed_pseudo | rs112295830 | YES | - |
| 10:50743043-5074 | - | intron_variant | MODIFIER | ASAH2B | ENSG0000020414 | Transcript | ENST0000064731 | protein_coding | rs112405752 | YES | MANE_Select |
| 10:50743043-5074 | - | upstream_gene_va | MODIFIER | - | ENSG0000028722 | Transcript | ENST0000067139 | lncRNA | rs112405752 | YES | - |
| 10:94358221-9435 | - | splice_region_vari | LOW | NOC3L | ENSG0000017314 | Transcript | ENST0000037136 | protein_coding | rs111938683 | YES | MANE_Select |
| 10:112404668-112 | - | intron_variant | MODIFIER | ACSL5 | ENSG0000019714 | Transcript | ENST0000035465 | protein_coding | - | YES | MANE_Select |
| 10:112404668-112 | - | downstream_gene_ | MODIFIER | - | ENSG0000023293 | Transcript | ENST0000063108 | lncRNA | - | YES | - |
| 10:113848892-113 | - | intron_variant | MODIFIER | DCLRE1A | ENSG0000019892 | Transcript | ENST0000036138 | protein_coding | - | YES | MANE_Select |
| 10:113848892-113 | - | intron_variant,non | MODIFIER | - | ENSG0000028893 | Transcript | ENST0000069264 | lncRNA | - | YES | - |
| 11:66850936-6685 | - | downstream_gene_ | MODIFIER | RCE1 | ENSG0000017365 | Transcript | ENST0000030965 | protein_coding | rs139065746 | YES | MANE_Select |
| 11:66850936-6685 | - | splice_polypyrimi | LOW | PC | ENSG0000017359 | Transcript | ENST0000039396 | protein_coding | rs139065746 | YES | MANE_Select |
| 11:75797346-7579 | - | intron_variant | MODIFIER | DGAT2 | ENSG0000006228 | Transcript | ENST0000022802 | protein_coding | - | YES | MANE_Select |

| | | | | | | | | | | | |
|---|---|---|---|---|---|---|---|---|---|---|---|
| 11:75797346-7579 | - | intron_variant,non_ | MODIFIER | UVRAG-DT | ENSG0000029362 | Transcript | ENST0000071620 | lncRNA | - | YES | - |
| 11:90071124-9007 | - | downstream_gene_ | MODIFIER | - | ENSG0000025488 | Transcript | ENST0000052781 | processed_pseudog | rs374502718 | YES | - |
| 11:90071124-9007 | - | upstream_gene_va | MODIFIER | - | ENSG0000025501 | Transcript | ENST0000053203 | unprocessed_pseud | rs374502718 | YES | - |
| 12:8868422-88684 | - | intron_variant | MODIFIER | A2ML1 | ENSG0000016653 | Transcript | ENST0000029969 | protein_coding | rs150983014 | YES | MANE_Select |
| 12:8868422-88684 | - | intron_variant,non_ | MODIFIER | - | ENSG0000028202 | Transcript | ENST0000063183 | lncRNA | rs150983014 | YES | - |
| 12:49656368-4965 | - | intron_variant | MODIFIER | FMNL3 | ENSG0000016179 | Transcript | ENST0000033515 | protein_coding | rs139157446 | YES | MANE_Select |
| 12:56417805-5641 | - | intron_variant | MODIFIER | TIMELESS | ENSG0000011160 | Transcript | ENST0000055353 | protein_coding | rs145784429 | YES | MANE_Select |
| 12:56417805-5641 | - | upstream_gene_va | MODIFIER | - | ENSG0000027627 | Transcript | ENST0000061044 | lncRNA | rs145784429 | YES | - |
| 12:56417805-5641 | - | upstream_gene_va | MODIFIER | - | ENSG0000027456 | Transcript | ENST0000062058 | unitary_pseudogen | rs145784429 | YES | - |
| 12:95898502-9589 | - | intron_variant | MODIFIER | CCDC38 | ENSG0000016597 | Transcript | ENST0000034428 | protein_coding | - | YES | MANE_Select |
| 12:113947257-113 | - | intron_variant | MODIFIER | RBM19 | ENSG0000012296 | Transcript | ENST0000026174 | protein_coding | - | YES | MANE_Select |
| 13:77070573-7707 | - | intron_variant | MODIFIER | MYCBP2 | ENSG0000000581 | Transcript | ENST0000054444 | protein_coding | rs60371737 | YES | MANE_Select |
| 13:99982753-9998 | - | upstream_gene_va | MODIFIER | - | ENSG0000028806 | Transcript | ENST0000065377 | lncRNA | - | YES | - |
| 13:101074692-101 | - | intron_variant | MODIFIER | NALCN | ENSG0000010245 | Transcript | ENST0000025112 | protein_coding | rs34229756 | YES | MANE_Select |
| 13:101090060-101 | - | intron_variant | MODIFIER | NALCN | ENSG0000010245 | Transcript | ENST0000025112 | protein_coding | - | YES | MANE_Select |
| 14:24163922-2416 | - | downstream_gene_ | MODIFIER | RNF31 | ENSG0000009209 | Transcript | ENST0000032410 | protein_coding | rs746378882 | YES | MANE_Select |
| 14:24163922-2416 | - | downstream_gene_ | MODIFIER | - | ENSG0000028975 | Transcript | ENST0000069902 | lncRNA | rs746378882 | YES | - |
| 14:24300643-2430 | - | upstream_gene_va | MODIFIER | DHRS1 | ENSG0000015737 | Transcript | ENST0000028811 | protein_coding | rs71119069 | YES | MANE_Select |
| 14:24300643-2430 | - | downstream_gene_ | MODIFIER | CIDEB | ENSG0000013630 | Transcript | ENST0000055441 | protein_coding | rs71119069 | YES | MANE_Select |
| 14:24300643-2430 | - | downstream_gene_ | MODIFIER | - | ENSG0000028804 | Transcript | ENST0000073790 | lncRNA | rs71119069 | YES | - |
| 14:61457374-6145 | - | intron_variant | MODIFIER | PRKCH | ENSG0000002707 | Transcript | ENST0000033298 | protein_coding | rs146597366 | YES | MANE_Select |
| 15:40036395-4003 | - | downstream_gene_ | MODIFIER | EIF2AK4 | ENSG0000012882 | Transcript | ENST0000026379 | protein_coding | rs371085676 | YES | MANE_Select |
| 15:40036395-4003 | - | upstream_gene_va | MODIFIER | SRP14-DT | ENSG0000024850 | Transcript | ENST0000074667 | lncRNA | rs371085676 | YES | - |
| 15:73288149-7328 | - | intron_variant | MODIFIER | NEO1 | ENSG0000006714 | Transcript | ENST0000026190 | protein_coding | - | YES | MANE_Select |
| 16:3284207-32842 | - | splice_donor_regio | LOW | ZNF263 | ENSG0000000619 | Transcript | ENST0000021906 | protein_coding | rs142708827 | YES | MANE_Select |
| 16:72788664-7278 | - | intron_variant,non_ | MODIFIER | ZFHX3-AS1 | ENSG0000025976 | Transcript | ENST0000076645 | lncRNA | - | YES | - |
| 16:72797458-7279 | - | intron_variant,non_ | MODIFIER | ZFHX3-AS1 | ENSG0000025976 | Transcript | ENST0000076645 | lncRNA | - | YES | - |
| 17:4809268-48092 | - | intron_variant | MODIFIER | PLD2 | ENSG0000012921 | Transcript | ENST0000026308 | protein_coding | - | YES | MANE_Select |
| 17:8160737-81607 | - | 3_prime_UTR_var | MODIFIER | VAMP2 | ENSG0000022020 | Transcript | ENST0000031650 | protein_coding | rs377440573 | YES | MANE_Select |
| 17:8160737-81607 | - | upstream_gene_va | MODIFIER | - | ENSG0000029995 | Transcript | ENST0000076770 | lncRNA | rs377440573 | YES | - |
| 17:28584629-2858 | - | intron_variant | MODIFIER | SPAG5 | ENSG0000007638 | Transcript | ENST0000032176 | protein_coding | - | YES | MANE_Select |
| 17:33112079-3311 | - | splice_polypyrimid | LOW | ASIC2 | ENSG0000010868 | Transcript | ENST0000022582 | protein_coding | rs140895516 | YES | MANE_Select |
| 17:33112079-3311 | - | non_coding_transc | MODIFIER | - | ENSG0000026653 | Transcript | ENST0000077595 | lncRNA | rs140895516 | YES | - |
| 17:39745549-3974 | - | intron_variant | MODIFIER | GRB7 | ENSG0000014173 | Transcript | ENST0000030915 | protein_coding | - | YES | MANE_Select |
| 17:42955992-4295 | - | intron_variant | MODIFIER | AARSD1 | ENSG0000026696 | Transcript | ENST0000042756 | protein_coding | rs151078413 | YES | MANE_Select |
| 17:60069867-6006 | - | intron_variant | MODIFIER | HEATR6 | ENSG0000006809 | Transcript | ENST0000018495 | protein_coding | rs201877781 | YES | MANE_Select |
| 19:1578372-15783 | - | downstream_gene_ | MODIFIER | - | ENSG0000027900 | Transcript | ENST0000062442 | TEC | rs371220154 | YES | - |

| | | | | | | | | | | | |
|---|---|---|---|---|---|---|---|---|---|---|---|
| 19:2733503-27335 | - | intron_variant | MODIFIER | SLC39A3 | ENSG0000014187 | Transcript | ENST0000026974 | protein_coding | - | YES | MANE_Select |
| 19:2733503-27335 | - | upstream_gene_va | MODIFIER | - | ENSG0000026134 | Transcript | ENST0000056790 | lncRNA | - | YES | - |
| 19:5239100-52391 | - | intron_variant | MODIFIER | PTPRS | ENSG0000010542 | Transcript | ENST0000026296 | protein_coding | rs149487300 | YES | MANE_Select |
| 19:20132594-2013 | - | splice_region_vari | LOW | - | ENSG0000026722 | Transcript | ENST0000059292 | transcribed_unproc | rs1184014518 | YES | - |
| 19:20132594-2013 | - | splice_region_vari | LOW | - | ENSG0000026738 | Transcript | ENST0000059365 | lncRNA | rs1184014518 | YES | - |
| 19:20132594-2013 | - | upstream_gene_va | MODIFIER | BNIP3P15 | ENSG0000027071 | Transcript | ENST0000060460 | processed_pseudog | rs1184014518 | YES | - |
| 19:41206254-4120 | - | intron_variant | MODIFIER | CYP2S1 | ENSG0000016760 | Transcript | ENST0000031005 | protein_coding | rs55800603 | YES | MANE_Select |
| 19:42739195-4273 | - | intron_variant | MODIFIER | PSG3 | ENSG0000022182 | Transcript | ENST0000032749 | protein_coding | rs372182118 | YES | MANE_Select |
| 19:48836425-4883 | - | downstream_gene_ | MODIFIER | PLEKHA4 | ENSG0000010555 | Transcript | ENST0000026326 | protein_coding | rs146134551 | YES | MANE_Select |
| 19:48836425-4883 | - | 5_prime_UTR_var | MODIFIER | HSD17B14 | ENSG0000008707 | Transcript | ENST0000026327 | protein_coding | rs146134551 | YES | MANE_Select |
| 19:58557202-5855 | - | downstream_gene_ | MODIFIER | MZF1 | ENSG0000009932 | Transcript | ENST0000021505 | protein_coding | - | YES | MANE_Select |
| 19:58557202-5855 | - | intron_variant | MODIFIER | UBE2M | ENSG0000013072 | Transcript | ENST0000025302 | protein_coding | - | YES | MANE_Select |
| 19:58557202-5855 | - | upstream_gene_va | MODIFIER | CHMP2A | ENSG0000013072 | Transcript | ENST0000031254 | protein_coding | - | YES | MANE_Select |
| 19:58557202-5855 | - | upstream_gene_va | MODIFIER | MZF1-AS1 | ENSG0000026785 | Transcript | ENST0000078983 | lncRNA | - | YES | - |
| 2:37372476-37372 | - | splice_donor_5th_ | LOW | QPCT | ENSG0000011582 | Transcript | ENST0000033841 | protein_coding | rs112031803 | YES | MANE_Select |
| 2:37372476-37372 | - | downstream_gene_ | MODIFIER | - | ENSG0000029646 | Transcript | ENST0000073982 | lncRNA | rs112031803 | YES | - |
| 2:38300255-38300 | - | intron_variant | MODIFIER | ATL2 | ENSG0000011978 | Transcript | ENST0000037895 | protein_coding | - | YES | MANE_Select |
| 2:70988753-70988 | - | intron_variant | MODIFIER | TEX261 | ENSG0000014404 | Transcript | ENST0000027243 | protein_coding | rs536484771 | YES | MANE_Select |
| 2:70988753-70988 | - | downstream_gene_ | MODIFIER | ANKRD53 | ENSG0000014403 | Transcript | ENST0000036058 | protein_coding | rs536484771 | YES | MANE_Select |
| 2:70988753-70988 | - | upstream_gene_va | MODIFIER | - | ENSG0000029913 | Transcript | ENST0000076070 | lncRNA | rs536484771 | YES | - |
| 2:134867548-1348 | - | intron_variant | MODIFIER | ACMSD | ENSG0000015308 | Transcript | ENST0000035614 | protein_coding | rs138317668 | YES | MANE_Select |
| 2:134867548-1348 | - | intron_variant,non | MODIFIER | CCNT2-AS1 | ENSG0000022404 | Transcript | ENST0000074780 | lncRNA | rs138317668 | YES | - |
| 2:164699566-1646 | - | intron_variant | MODIFIER | COBLL1 | ENSG0000008243 | Transcript | ENST0000065265 | protein_coding | rs372370676 | YES | MANE_Select |
| 2:196201452-1962 | - | intron_variant | MODIFIER | HECW2 | ENSG0000013841 | Transcript | ENST0000064497 | protein_coding | rs561575955,COS | YES | MANE_Select |
| 2:222293705-2222 | - | intron_variant | MODIFIER | PAX3 | ENSG0000013590 | Transcript | ENST0000039207 | protein_coding | rs150420509 | YES | MANE_Select |
| 2:222293705-2222 | - | upstream_gene_va | MODIFIER | CCDC140 | ENSG0000016308 | Transcript | ENST0000064776 | lncRNA | rs150420509 | YES | - |
| 2:223895642-2238 | - | splice_polypyrimid | LOW | WDFY1 | ENSG0000008544 | Transcript | ENST0000023305 | protein_coding | rs150347314 | YES | MANE_Select |
| 2:232378818-2323 | - | intron_variant,non | MODIFIER | ECEL1P2 | ENSG0000022451 | Transcript | ENST0000071529 | lncRNA | rs377162921 | YES | - |
| 2:233769464-2337 | - | intron_variant | MODIFIER | UGT1A6 | ENSG0000016716 | Transcript | ENST0000030513 | protein_coding | - | YES | MANE_Select |
| 2:233769464-2337 | - | intron_variant | MODIFIER | UGT1A1 | ENSG0000024163 | Transcript | ENST0000030520 | protein_coding | - | YES | MANE_Select |
| 2:233769464-2337 | - | intron_variant | MODIFIER | UGT1A10 | ENSG0000024251 | Transcript | ENST0000034464 | protein_coding | - | YES | MANE_Select |
| 2:233769464-2337 | - | intron_variant | MODIFIER | UGT1A9 | ENSG0000024111 | Transcript | ENST0000035472 | protein_coding | - | YES | MANE_Select |
| 2:233769464-2337 | - | intron_variant | MODIFIER | UGT1A4 | ENSG0000024447 | Transcript | ENST0000037340 | protein_coding | - | YES | MANE_Select |
| 2:233769464-2337 | - | intron_variant | MODIFIER | UGT1A5 | ENSG0000028870 | Transcript | ENST0000037341 | protein_coding | - | YES | MANE_Select |
| 2:233769464-2337 | - | intron_variant | MODIFIER | UGT1A7 | ENSG0000024412 | Transcript | ENST0000037342 | protein_coding | - | YES | MANE_Select |
| 2:233769464-2337 | - | intron_variant | MODIFIER | UGT1A8 | ENSG0000024236 | Transcript | ENST0000037345 | protein_coding | - | YES | MANE_Select |
| 2:233769464-2337 | - | intron_variant | MODIFIER | UGT1A3 | ENSG0000028870 | Transcript | ENST0000048202 | protein_coding | - | YES | MANE_Select |

| | | | | | | | | | | | |
|---|---|---|---|---|---|---|---|---|---|---|---|
| 2:233769464-2337 | - | downstream_gene_ | MODIFIER | - | ENSG0000029847 | Transcript | ENST0000075573 | lncRNA | - | YES | - |
| 20:31472917-3147 | - | upstream_gene_va | MODIFIER | REM1 | ENSG0000008832 | Transcript | ENST0000020197 | protein_coding | - | YES | MANE_Select |
| 20:31472917-3147 | - | intron_variant | MODIFIER | DEFB124 | ENSG0000018038 | Transcript | ENST0000031767 | protein_coding | - | YES | MANE_Select |
| 20:34743163-3474 | - | intron_variant,non | MODIFIER | - | ENSG0000028680 | Transcript | ENST0000065504 | lncRNA | rs112051697 | YES | - |
| 20:47169101-4716 | - | intron_variant | MODIFIER | EYA2 | ENSG0000006465 | Transcript | ENST0000032761 | protein_coding | - | YES | MANE_Select |
| 20:47169101-4716 | - | downstream_gene_ | MODIFIER | MIR3616 | ENSG0000026490 | Transcript | ENST0000058407 | miRNA | - | YES | - |
| 20:49011831-4901 | - | intron_variant | MODIFIER | ARFGEF2 | ENSG0000012419 | Transcript | ENST0000037191 | protein_coding | - | YES | MANE_Select |
| 20:56248628-5624 | - | upstream_gene_va | MODIFIER | MC3R | ENSG0000012408 | Transcript | ENST0000024391 | protein_coding | - | YES | MANE_Select |
| 21:34708146-3470 | - | intron_variant | MODIFIER | CLIC6 | ENSG0000015921 | Transcript | ENST0000034949 | protein_coding | - | YES | MANE_Select |
| 21:45917570-4591 | - | splice_polypyrimic | LOW | - | ENSG0000028060 | Transcript | ENST0000062623 | lncRNA | - | YES | - |
| 21:45917570-4591 | - | splice_polypyrimic | LOW | PCBP3 | ENSG0000018357 | Transcript | ENST0000068168 | protein_coding | - | YES | MANE_Select |
| 3:58134591-58134 | - | intron_variant | MODIFIER | FLNB | ENSG0000013606 | Transcript | ENST0000029595 | protein_coding | - | YES | MANE_Select |
| 3:65375708-65375 | - | intron_variant | MODIFIER | MAGI1 | ENSG0000015127 | Transcript | ENST0000040293 | protein_coding | rs3836224 | YES | MANE_Select |
| 3:65387174-65387 | - | intron_variant | MODIFIER | MAGI1 | ENSG0000015127 | Transcript | ENST0000040293 | protein_coding | rs139764373 | YES | MANE_Select |
| 3:128573475-1285 | - | intron_variant,non | MODIFIER | LINC01565 | ENSG0000019868 | Transcript | ENST0000075324 | lncRNA | rs147519916 | YES | - |
| 4:654753-654757 | - | intron_variant,non | MODIFIER | PDE6B-AS1 | ENSG0000024268 | Transcript | ENST0000046835 | lncRNA | - | YES | - |
| 4:654753-654757 | - | intron_variant | MODIFIER | PDE6B | ENSG0000013325 | Transcript | ENST0000049651 | protein_coding | - | YES | MANE_Select |
| 5:43645561-43645 | - | intron_variant | MODIFIER | NNT | ENSG0000011299 | Transcript | ENST0000034492 | protein_coding | rs71974629 | YES | MANE_Select |
| 5:170080126-1700 | - | intron_variant | MODIFIER | DOCK2 | ENSG0000013451 | Transcript | ENST0000052090 | protein_coding | rs138478687 | YES | MANE_Select |
| 6:83407914-83407 | - | splice_polypyrimic | LOW | ME1 | ENSG0000006583 | Transcript | ENST0000036970 | protein_coding | rs747274917 | YES | MANE_Select |
| 6:112141268-1121 | - | intron_variant | MODIFIER | LAMA4 | ENSG0000011276 | Transcript | ENST0000023053 | protein_coding | rs149046391 | YES | MANE_Select |
| 6:112141268-1121 | - | intron_variant,non | MODIFIER | - | ENSG0000023723 | Transcript | ENST0000078458 | lncRNA | rs149046391 | YES | - |
| 6:117703542-1177 | - | intron_variant | MODIFIER | NUS1 | ENSG0000015398 | Transcript | ENST0000036849 | protein_coding | rs146116464 | YES | MANE_Select |
| 6:132463932-1324 | - | intron_variant | MODIFIER | STX7 | ENSG0000007995 | Transcript | ENST0000036794 | protein_coding | rs112655932 | YES | MANE_Select |
| 6:144700057-1447 | - | intron_variant | MODIFIER | UTRN | ENSG0000015281 | Transcript | ENST0000036754 | protein_coding | rs140389119 | YES | MANE_Select |
| 7:20140884-20140 | - | 3_prime_UTR_var | MODIFIER | MACC1 | ENSG0000018374 | Transcript | ENST0000040033 | protein_coding | rs55989776 | YES | MANE_Select |
| 7:20140884-20140 | - | intron_variant,non | MODIFIER | MACC1-AS1 | ENSG0000022859 | Transcript | ENST0000073867 | lncRNA | rs55989776 | YES | - |
| 7:26725372-26725 | - | intron_variant | MODIFIER | SKAP2 | ENSG0000000502 | Transcript | ENST0000034531 | protein_coding | rs139615753 | YES | MANE_Select |
| 7:29504721-29504 | - | intron_variant | MODIFIER | CHN2 | ENSG0000010606 | Transcript | ENST0000022279 | protein_coding | rs10633708 | YES | MANE_Select |
| 7:38242529-38242 | - | intron_variant | MODIFIER | TRGC2 | ENSG0000022719 | Transcript | ENST0000043691 | TR_C_gene | rs35917252 | YES | - |
| 7:38245284-38245 | - | intron_variant | MODIFIER | TRGC2 | ENSG0000022719 | Transcript | ENST0000043691 | TR_C_gene | rs138091320 | YES | - |
| 7:47413908-47413 | - | intron_variant | MODIFIER | TNS3 | ENSG0000013620 | Transcript | ENST0000031116 | protein_coding | rs548858209 | YES | MANE_Select |
| 7:96028144-96028 | - | intron_variant | MODIFIER | DYNC1I1 | ENSG0000015856 | Transcript | ENST0000044746 | protein_coding | rs145322007 | YES | MANE_Select |
| 7:123689006-1236 | - | intron_variant | MODIFIER | WASL | ENSG0000010629 | Transcript | ENST0000022302 | protein_coding | - | YES | MANE_Select |
| 7:142474204-1424 | - | downstream_gene_ | MODIFIER | TRBV6-6 | ENSG0000021172 | Transcript | ENST0000039037 | TR_V_gene | rs140850096 | YES | - |
| 7:142474204-1424 | - | intron_variant,non | MODIFIER | TRBV7-5 | ENSG0000022755 | Transcript | ENST0000039038 | TR_V_pseudogene | rs140850096 | YES | - |
| 8:140532162-1405 | - | splice_polypyrimic | LOW | AGO2 | ENSG0000012390 | Transcript | ENST0000022059 | protein_coding | rs565333105 | YES | MANE_Select |

| | | | | | | | | | | | |
|---|---|---|---|---|---|---|---|---|---|---|---|
| 8:144317162-1443 | - | upstream_gene_va | MODIFIER | - | ENSG0000025469 | Transcript | ENST0000052502 | lncRNA | rs201213845 | YES | - |
| 8:144317162-1443 | - | intron_variant | MODIFIER | DGAT1 | ENSG0000018500 | Transcript | ENST0000052871 | protein_coding | rs201213845 | YES | MANE_Select |
| 8:144317162-1443 | - | downstream_gene_ | MODIFIER | HSF1 | ENSG0000018512 | Transcript | ENST0000052883 | protein_coding | rs201213845 | YES | MANE_Select |
| 8:144317162-1443 | - | downstream_gene_ | MODIFIER | MIR6848 | ENSG0000028422 | Transcript | ENST0000061121 | miRNA | rs201213845 | YES | - |
| 9:18474375-18474 | - | intron_variant | MODIFIER | ADAMTSL1 | ENSG0000017803 | Transcript | ENST0000038054 | protein_coding | - | YES | MANE_Select |
| 9:33795614-33795 | TA | intron_variant,non_ | MODIFIER | UBE2R2-AS1 | ENSG0000023548 | Transcript | ENST0000070503 | lncRNA | rs1188045977 | YES | - |
| 9:35825544-35825 | - | intron_variant | MODIFIER | FAM221B | ENSG0000020493 | Transcript | ENST0000042353 | protein_coding | rs201802596 | YES | MANE_Select |
| 9:35825544-35825 | - | upstream_gene_va | MODIFIER | TMEM8B | ENSG0000013710 | Transcript | ENST0000064393 | protein_coding | rs201802596 | YES | MANE_Select |
| 9:36923306-36923 | - | intron_variant | MODIFIER | PAX5 | ENSG0000019609 | Transcript | ENST0000035812 | protein_coding | - | YES | MANE_Select |
| X:139958016-139 | - | intron_variant | MODIFIER | CXorf66 | ENSG0000020393 | Transcript | ENST0000037054 | protein_coding | rs753088382 | YES | MANE_Select |
| **PRAD** | | | | | | | | | | | |
| 10:69414880-6941 | - | intron_variant | MODIFIER | TACR2 | ENSG0000007507 | Transcript | ENST0000037330 | protein_coding | rs150956058 | YES | MANE_Select |
| 10:69414880-6941 | - | intron_variant,non_ | MODIFIER | - | ENSG0000029948 | Transcript | ENST0000076385 | lncRNA | rs150956058 | YES | - |
| **BLGG** | | | | | | | | | | | |
| 1:20857326-20857 | - | intron_variant | MODIFIER | EIF4G3 | ENSG0000007515 | Transcript | ENST0000060232 | protein_coding | - | YES | MANE_Select |
| 1:225847138-2258 | - | downstream_gene_ | MODIFIER | EPHX1 | ENSG0000014381 | Transcript | ENST0000027216 | protein_coding | - | YES | MANE_Select |
| 1:225847138-2258 | - | upstream_gene_va | MODIFIER | - | ENSG0000024286 | Transcript | ENST0000042433 | lncRNA | - | YES | - |
| 17:32997377-3299 | - | intron_variant | MODIFIER | SPACA3 | ENSG0000014131 | Transcript | ENST0000026905 | protein_coding | - | YES | MANE_Select |
| 19:7678296-76783 | - | intron_variant | MODIFIER | MCEMP1 | ENSG0000018301 | Transcript | ENST0000033359 | protein_coding | - | YES | MANE_Select |
| 19:7678296-76783 | - | upstream_gene_va | MODIFIER | TRAPPC5 | ENSG0000018102 | Transcript | ENST0000059614 | protein_coding | - | YES | MANE_Select |
| 19:19833449-1983 | - | intron_variant,non_ | MODIFIER | ZNF56P | ENSG0000029117 | Transcript | ENST0000059188 | lncRNA | - | YES | - |
| 19:19833449-1983 | - | upstream_gene_va | MODIFIER | ZNF56P | ENSG0000026741 | Transcript | ENST0000062341 | transcribed_unproc | - | YES | - |
| 19:19833449-1983 | - | intron_variant,non_ | MODIFIER | - | ENSG0000029615 | Transcript | ENST0000073692 | lncRNA | - | YES | - |
| 19:20184331-2018 | - | intron_variant | MODIFIER | ZNF486 | ENSG0000025622 | Transcript | ENST0000033511 | protein_coding | rs367731486 | YES | MANE_Select |
| 19:20184331-2018 | - | intron_variant,non_ | MODIFIER | - | ENSG0000026738 | Transcript | ENST0000059365 | lncRNA | rs367731486 | YES | - |
| 19:38408861-3840 | - | downstream_gene_ | MODIFIER | RASGRP4 | ENSG0000017177 | Transcript | ENST0000061543 | protein_coding | - | YES | MANE_Select |
| 20:56248628-5624 | - | upstream_gene_va | MODIFIER | MC3R | ENSG0000012408 | Transcript | ENST0000024391 | protein_coding | - | YES | MANE_Select |
| 3:187253326-1872 | - | splice_polypyrimic | LOW | MASP1 | ENSG0000012724 | Transcript | ENST0000029628 | protein_coding | rs745796040 | YES | MANE_Select |
| 6:133889359-1338 | - | 5_prime_UTR_var | MODIFIER | TCF21 | ENSG0000011852 | Transcript | ENST0000036788 | protein_coding | - | YES | MANE_Select |
| 6:133889359-1338 | - | upstream_gene_va | MODIFIER | TARID | ENSG0000022795 | Transcript | ENST0000079540 | lncRNA | - | YES | - |
| 6:151458285-1514 | - | intron_variant | MODIFIER | ARMT1 | ENSG0000014647 | Transcript | ENST0000036729 | protein_coding | - | YES | MANE_Select |
| 7:2945755-294575 | - | intron_variant | MODIFIER | CARD11 | ENSG0000019828 | Transcript | ENST0000039694 | protein_coding | - | YES | MANE_Select |
| 7:2945755-294575 | - | intron_variant,non_ | MODIFIER | CARD11-AS1 | ENSG0000023728 | Transcript | ENST0000081660 | lncRNA | - | YES | - |
| 7:130041131-1300 | - | intron_variant | MODIFIER | ZC3HC1 | ENSG0000009173 | Transcript | ENST0000035830 | protein_coding | - | YES | MANE_Select |
| 9:35825544-35825 | - | intron_variant | MODIFIER | FAM221B | ENSG0000020493 | Transcript | ENST0000042353 | protein_coding | - | YES | MANE_Select |
| 9:35825544-35825 | - | upstream_gene_va | MODIFIER | TMEM8B | ENSG0000013710 | Transcript | ENST0000064393 | protein_coding | - | YES | MANE_Select |
| 9:130668313-1306 | - | splice_donor_regic | LOW | PRDM12 | ENSG0000013071 | Transcript | ENST0000025300 | protein_coding | - | YES | MANE_Select |

| HNSC | | | | | | | | | | | |
|---|---|---|---|---|---|---|---|---|---|---|---|
| 1:6469122-646912 | - | upstream_gene_va | MODIFIER | TNFRSF25 | ENSG0000021578 | Transcript | ENST0000035687 | protein_coding | rs113541584 | YES | MANE_Select |
| 1:20857326-20857 | - | intron_variant | MODIFIER | EIF4G3 | ENSG0000007515 | Transcript | ENST0000060232 | protein_coding | - | YES | MANE_Select |
| 1:155047968-1550 | - | intron_variant | MODIFIER | DCST1 | ENSG0000016335 | Transcript | ENST0000029554 | protein_coding | rs111941107 | YES | MANE_Select |
| 1:155047968-1550 | - | upstream_gene_va | MODIFIER | ADAM15 | ENSG0000014353 | Transcript | ENST0000035695 | protein_coding | rs111941107 | YES | MANE_Select |
| 1:155047968-1550 | - | intron_variant,non | MODIFIER | DCST1-AS1 | ENSG0000023209 | Transcript | ENST0000045296 | lncRNA | rs111941107 | YES | - |
| 1:184719103-1847 | - | intron_variant | MODIFIER | EDEM3 | ENSG0000011640 | Transcript | ENST0000031813 | protein_coding | rs111917956 | YES | MANE_Select |
| 1:233353959-2333 | - | splice_donor_5th_ | LOW | MAP3K21 | ENSG0000014367 | Transcript | ENST0000036662 | protein_coding | rs367946480 | YES | MANE_Select |
| 1:233353962-2333 | - | splice_region_vari | LOW | MAP3K21 | ENSG0000014367 | Transcript | ENST0000036662 | protein_coding | rs537474473 | YES | MANE_Select |
| 10:50743043-5074 | - | intron_variant | MODIFIER | ASAH2B | ENSG0000020414 | Transcript | ENST0000064731 | protein_coding | rs112405752 | YES | MANE_Select |
| 10:50743043-5074 | - | upstream_gene_va | MODIFIER | - | ENSG0000028722 | Transcript | ENST0000067139 | lncRNA | rs112405752 | YES | - |
| 10:69414880-6941 | - | intron_variant | MODIFIER | TACR2 | ENSG0000007507 | Transcript | ENST0000037330 | protein_coding | rs150956058 | YES | MANE_Select |
| 10:69414880-6941 | - | intron_variant,non | MODIFIER | - | ENSG0000029948 | Transcript | ENST0000076385 | lncRNA | rs150956058 | YES | - |
| 10:112404668-112 | - | intron_variant | MODIFIER | ACSL5 | ENSG0000019714 | Transcript | ENST0000035465 | protein_coding | rs139652367 | YES | MANE_Select |
| 10:112404668-112 | - | downstream_gene_ | MODIFIER | - | ENSG0000023293 | Transcript | ENST0000063108 | lncRNA | rs139652367 | YES | - |
| 11:62516707-6251 | - | 3_prime_UTR_var | MODIFIER | AHNAK | ENSG0000012494 | Transcript | ENST0000037802 | protein_coding | rs142437234 | YES | MANE_Select |
| 12:6548236-65482 | - | downstream_gene_ | MODIFIER | - | ENSG0000024566 | Transcript | ENST0000049920 | lncRNA | rs138979230 | YES | - |
| 12:6548236-65482 | - | intron_variant | MODIFIER | IFFO1 | ENSG0000001029 | Transcript | ENST0000061957 | protein_coding | rs138979230 | YES | MANE_Select |
| 12:7067792-70677 | - | intron_variant | MODIFIER | C1S | ENSG0000018232 | Transcript | ENST0000036081 | protein_coding | rs150772912 | YES | MANE_Select |
| 12:57535460-5753 | - | intron_variant | MODIFIER | DCTN2 | ENSG0000017520 | Transcript | ENST0000054824 | protein_coding | rs143028062 | YES | MANE_Select |
| 12:110914159-110 | - | intron_variant | MODIFIER | MYL2 | ENSG0000011124 | Transcript | ENST0000022884 | protein_coding | rs142567411 | YES | MANE_Select |
| 12:113947257-113 | - | intron_variant | MODIFIER | RBM19 | ENSG0000012296 | Transcript | ENST0000026174 | protein_coding | rs369688809 | YES | MANE_Select |
| 13:73062772-7306 | - | intron_variant | MODIFIER | KLF5 | ENSG0000010255 | Transcript | ENST0000037768 | protein_coding | rs113148226 | YES | MANE_Select |
| 13:77070573-7707 | - | intron_variant | MODIFIER | MYCBP2 | ENSG0000000581 | Transcript | ENST0000054444 | protein_coding | rs60371737 | YES | MANE_Select |
| 13:99982753-9998 | - | upstream_gene_va | MODIFIER | - | ENSG0000028806 | Transcript | ENST0000065377 | lncRNA | rs398124241 | YES | - |
| 16:3284207-32842 | - | splice_donor_regic | LOW | ZNF263 | ENSG0000000619 | Transcript | ENST0000021906 | protein_coding | rs142708827 | YES | MANE_Select |
| 17:32997377-3299 | - | intron_variant | MODIFIER | SPACA3 | ENSG0000014131 | Transcript | ENST0000026905 | protein_coding | rs538091519 | YES | MANE_Select |
| 17:40822513-4082 | - | intron_variant,non | MODIFIER | KRT10-AS1 | ENSG0000016792 | Transcript | ENST0000030166 | lncRNA | rs148510452 | YES | - |
| 18:77268542-7726 | - | intron_variant | MODIFIER | GALR1 | ENSG0000016657 | Transcript | ENST0000029972 | protein_coding | rs71876994 | YES | MANE_Select |
| 19:2733503-27335 | - | intron_variant | MODIFIER | SLC39A3 | ENSG0000014187 | Transcript | ENST0000026974 | protein_coding | rs139950756 | YES | MANE_Select |
| 19:2733503-27335 | - | upstream_gene_va | MODIFIER | - | ENSG0000026134 | Transcript | ENST0000056790 | lncRNA | rs139950756 | YES | - |
| 19:20646412-2064 | - | splice_region_vari | LOW | ZNF626 | ENSG0000018817 | Transcript | ENST0000060144 | protein_coding | rs374238712 | YES | MANE_Select |
| 19:42739195-4273 | - | intron_variant | MODIFIER | PSG3 | ENSG0000022182 | Transcript | ENST0000032749 | protein_coding | rs372182118 | YES | MANE_Select |
| 19:58557202-5855 | - | downstream_gene_ | MODIFIER | MZF1 | ENSG0000009932 | Transcript | ENST0000021505 | protein_coding | rs145976499 | YES | MANE_Select |
| 19:58557202-5855 | - | intron_variant | MODIFIER | UBE2M | ENSG0000013072 | Transcript | ENST0000025302 | protein_coding | rs145976499 | YES | MANE_Select |
| 19:58557202-5855 | - | upstream_gene_va | MODIFIER | CHMP2A | ENSG0000013072 | Transcript | ENST0000031254 | protein_coding | rs145976499 | YES | MANE_Select |
| 19:58557202-5855 | - | upstream_gene_va | MODIFIER | MZF1-AS1 | ENSG0000026785 | Transcript | ENST0000078983 | lncRNA | rs145976499 | YES | - |

| | | | | | | | | | | | |
|---|---|---|---|---|---|---|---|---|---|---|---|
| 2:226796679-2267 | - | intron_variant,non | MODIFIER | - | ENSG0000027262 | Transcript | ENST0000072765 | lncRNA | CD951753 | YES | - |
| 2:226796679-2267 | - | downstream_gene_ | MODIFIER | - | ENSG0000029507 | Transcript | ENST0000072776 | lncRNA | CD951753 | YES | - |
| 3:128573475-1285 | - | intron_variant,non | MODIFIER | LINC01565 | ENSG0000019868 | Transcript | ENST0000075324 | lncRNA | rs147519916 | YES | - |
| 3:160502049-1605 | - | 3_prime_UTR_var | MODIFIER | KPNA4 | ENSG0000018643 | Transcript | ENST0000033425 | protein_coding | rs139516169 | YES | MANE_Select |
| 3:187262497-1872 | - | intron_variant | MODIFIER | MASP1 | ENSG0000012724 | Transcript | ENST0000029628 | protein_coding | rs141348175 | YES | MANE_Select |
| 5:170080126-1700 | - | intron_variant | MODIFIER | DOCK2 | ENSG0000013451 | Transcript | ENST0000052090 | protein_coding | rs138478687 | YES | MANE_Select |
| 5:180567815-1805 | - | intron_variant | MODIFIER | CNOT6 | ENSG0000011330 | Transcript | ENST0000026195 | protein_coding | rs146190811 | YES | MANE_Select |
| 6:32179514-32179 | - | downstream_gene_ | MODIFIER | AGER | ENSG0000020430 | Transcript | ENST0000037507 | protein_coding | rs374442418 | YES | MANE_Select |
| 6:32179514-32179 | - | intron_variant | MODIFIER | RNF5 | ENSG0000020430 | Transcript | ENST0000037509 | protein_coding | rs374442418 | YES | MANE_Select |
| 6:32179514-32179 | - | upstream_gene_va | MODIFIER | AGPAT1 | ENSG0000020431 | Transcript | ENST0000037510 | protein_coding | rs374442418 | YES | MANE_Select |
| 6:32179514-32179 | - | upstream_gene_va | MODIFIER | MIR6833 | ENSG0000027726 | Transcript | ENST0000062068 | miRNA | rs374442418 | YES | - |
| 6:32179514-32179 | - | downstream_gene_ | MODIFIER | - | ENSG0000030175 | Transcript | ENST0000078134 | lncRNA | rs374442418 | YES | - |
| 6:132463932-1324 | - | intron_variant | MODIFIER | STX7 | ENSG0000007995 | Transcript | ENST0000036794 | protein_coding | - | YES | MANE_Select |
| 6:151458285-1514 | - | intron_variant | MODIFIER | ARMT1 | ENSG0000014647 | Transcript | ENST0000036729 | protein_coding | rs142167376 | YES | MANE_Select |
| 6:159763105-1597 | - | intron_variant | MODIFIER | ACAT2 | ENSG0000012043 | Transcript | ENST0000036704 | protein_coding | rs112953742 | YES | MANE_Select |
| 6:159763105-1597 | - | upstream_gene_va | MODIFIER | SOD2-OT1 | ENSG0000028542 | Transcript | ENST0000053537 | lncRNA | rs112953742 | YES | - |
| 7:20140884-20140 | - | 3_prime_UTR_var | MODIFIER | MACC1 | ENSG0000018374 | Transcript | ENST0000040033 | protein_coding | rs55989776 | YES | MANE_Select |
| 7:20140884-20140 | - | intron_variant,non | MODIFIER | MACC1-AS1 | ENSG0000022859 | Transcript | ENST0000073867 | lncRNA | rs55989776 | YES | - |
| 7:29367902-29367 | - | intron_variant | MODIFIER | CHN2 | ENSG0000010606 | Transcript | ENST0000022279 | protein_coding | rs142914048 | YES | MANE_Select |
| 7:96028144-96028 | - | intron_variant | MODIFIER | DYNC1I1 | ENSG0000015856 | Transcript | ENST0000044746 | protein_coding | rs145322007 | YES | MANE_Select |
| 7:130041131-1300 | - | intron_variant | MODIFIER | ZC3HC1 | ENSG0000009173 | Transcript | ENST0000035830 | protein_coding | rs375888799 | YES | MANE_Select |
| 8:74849303-74849 | - | 3_prime_UTR_var | MODIFIER | PI15 | ENSG0000013755 | Transcript | ENST0000026011 | protein_coding | rs141226749 | YES | MANE_Select |
| 8:74849303-74849 | - | intron_variant,non | MODIFIER | - | ENSG0000025370 | Transcript | ENST0000081017 | lncRNA | rs141226749 | YES | - |
| 8:144317162-1443 | - | upstream_gene_va | MODIFIER | - | ENSG0000025469 | Transcript | ENST0000052502 | lncRNA | rs201213845 | YES | - |
| 8:144317162-1443 | - | intron_variant | MODIFIER | DGAT1 | ENSG0000018500 | Transcript | ENST0000052871 | protein_coding | rs201213845 | YES | MANE_Select |
| 8:144317162-1443 | - | downstream_gene_ | MODIFIER | HSF1 | ENSG0000018512 | Transcript | ENST0000052883 | protein_coding | rs201213845 | YES | MANE_Select |
| 8:144317162-1443 | - | downstream_gene_ | MODIFIER | MIR6848 | ENSG0000028422 | Transcript | ENST0000061121 | miRNA | rs201213845 | YES | - |
| **BRCA** | | | | | | | | | | | |
| 1:155047968-1550 | - | intron_variant | MODIFIER | DCST1 | ENSG0000016335 | Transcript | ENST0000029554 | protein_coding | rs111941107 | YES | MANE_Select |
| 1:155047968-1550 | - | upstream_gene_va | MODIFIER | ADAM15 | ENSG0000014353 | Transcript | ENST0000035695 | protein_coding | rs111941107 | YES | MANE_Select |
| 1:155047968-1550 | - | intron_variant,non | MODIFIER | DCST1-AS1 | ENSG0000023209 | Transcript | ENST0000045296 | lncRNA | rs111941107 | YES | - |
| 1:193142098-1931 | - | intron_variant | MODIFIER | CDC73 | ENSG0000013437 | Transcript | ENST0000036743 | protein_coding | rs80356646 | YES | MANE_Select |
| 1:193142098-1931 | - | intron_variant,non | MODIFIER | - | ENSG0000030828 | Transcript | ENST0000083298 | lncRNA | rs80356646 | YES | - |
| 12:49656368-4965 | - | intron_variant | MODIFIER | FMNL3 | ENSG0000016179 | Transcript | ENST0000033515 | protein_coding | - | YES | MANE_Select |
| 12:80238816-8023 | - | intron_variant | MODIFIER | OTOGL | ENSG0000016589 | Transcript | ENST0000054710 | protein_coding | rs143126749 | YES | MANE_Select |
| 2:217808507-2178 | - | intron_variant | MODIFIER | TNS1 | ENSG0000007930 | Transcript | ENST0000068225 | protein_coding | rs112279658 | YES | MANE_Select |
| X:52584505-5258 | - | intron_variant,non | MODIFIER | - | ENSG0000022877 | Transcript | ENST0000042216 | transcribed_unproc | rs373343477 | YES | - |

| | | | | | | | | | | | |
|---|---|---|---|---|---|---|---|---|---|---|---|
| X:52584505-52584 | - | intron_variant,non_ | MODIFIER | - | ENSG0000030118 | Transcript | ENST0000077683 | lncRNA | rs373343477 | YES | - |

| Low/Modifier Impact Variants | | | | | | | | | | | |
|---|---|---|---|---|---|---|---|---|---|---|---|
| RS | | | | | | | | | | | |
| Squamous | | | | | | | | | | | |
| Location | Allele | Consequence | IMPACT | SYMBOL | Gene | Feature_type | Feature | BIOTYPE | Existing_variation | CANONICAL | MANE |
| 1:20857326-20857 | - | intron_variant | MODIFIER | EIF4G3 | ENSG0000007515 | Transcript | ENST0000060232 | protein_coding | rs145504996 | YES | MANE_Select |
| 11:26671047-2667 | - | 3_prime_UTR_var | MODIFIER | SLC5A12 | ENSG0000014894 | Transcript | ENST0000039600 | protein_coding | rs71449136 | YES | MANE_Select |
| 17:17819267-1781 | - | intron_variant | MODIFIER | SREBF1 | ENSG0000007231 | Transcript | ENST0000026164 | protein_coding | rs72248634 | YES | MANE_Select |
| 19:20184331-2018 | - | intron_variant | MODIFIER | ZNF486 | ENSG0000025622 | Transcript | ENST0000033511 | protein_coding | rs367731486 | YES | MANE_Select |
| 19:20184331-2018 | - | intron_variant,non | MODIFIER | - | ENSG0000026738 | Transcript | ENST0000059365 | lncRNA | rs367731486 | YES | - |
| 2:37372476-37372 | - | splice_region_vari | LOW | QPCT | ENSG0000011582 | Transcript | ENST0000033841 | protein_coding | - | YES | MANE_Select |
| 2:37372476-37372 | - | downstream_gene_ | MODIFIER | - | ENSG0000029646 | Transcript | ENST0000073982 | lncRNA | - | YES | - |
| 6:151458285-1514 | - | intron_variant | MODIFIER | ARMT1 | ENSG0000014647 | Transcript | ENST0000036729 | protein_coding | - | YES | MANE_Select |
| Adenocarcinoma | | | | | | | | | | | |
| 1:6469122-646912 | - | upstream_gene_va | MODIFIER | TNFRSF25 | ENSG0000021578 | Transcript | ENST0000035687 | protein_coding | - | YES | MANE_Select |
| 1:20857326-20857 | - | intron_variant | MODIFIER | EIF4G3 | ENSG0000007515 | Transcript | ENST0000060232 | protein_coding | - | YES | MANE_Select |
| 1:25227386-25227 | - | intron_variant | MODIFIER | SYF2 | ENSG0000011761 | Transcript | ENST0000023627 | protein_coding | - | YES | MANE_Select |
| 10:30027571-3002 | - | non_coding_transc | MODIFIER | - | ENSG0000030452 | Transcript | ENST0000080435 | lncRNA | - | YES | - |
| 10:50743043-5074 | - | intron_variant | MODIFIER | ASAH2B | ENSG0000020414 | Transcript | ENST0000064731 | protein_coding | - | YES | MANE_Select |
| 10:50743043-5074 | - | upstream_gene_va | MODIFIER | - | ENSG0000028722 | Transcript | ENST0000067139 | lncRNA | - | YES | - |
| 10:94358221-9435 | - | splice_polypyrimid | LOW | NOC3L | ENSG0000017314 | Transcript | ENST0000037136 | protein_coding | - | YES | MANE_Select |
| 10:112404668-112 | - | intron_variant | MODIFIER | ACSL5 | ENSG0000019714 | Transcript | ENST0000035465 | protein_coding | rs139652367 | YES | MANE_Select |
| 10:112404668-112 | - | downstream_gene_ | MODIFIER | - | ENSG0000023293 | Transcript | ENST0000063108 | lncRNA | rs139652367 | YES | - |
| 11:75797346-7579 | - | intron_variant | MODIFIER | DGAT2 | ENSG0000006228 | Transcript | ENST0000022802 | protein_coding | rs149770132 | YES | MANE_Select |
| 11:75797346-7579 | - | intron_variant,non | MODIFIER | UVRAG-DT | ENSG0000029362 | Transcript | ENST0000071620 | lncRNA | rs149770132 | YES | - |
| 11:90071124-9007 | - | downstream_gene_ | MODIFIER | - | ENSG0000025488 | Transcript | ENST0000052781 | processed_pseudogene | - | YES | - |
| 11:90071124-9007 | - | upstream_gene_va | MODIFIER | - | ENSG0000025501 | Transcript | ENST0000053203 | unprocessed_pseudogene | - | YES | - |
| 12:8868422-88684 | - | intron_variant | MODIFIER | A2ML1 | ENSG0000016653 | Transcript | ENST0000029969 | protein_coding | - | YES | MANE_Select |
| 12:8868422-88684 | - | intron_variant,non | MODIFIER | - | ENSG0000028202 | Transcript | ENST0000063183 | lncRNA | - | YES | - |
| 12:56417805-5641 | - | intron_variant | MODIFIER | TIMELESS | ENSG0000011160 | Transcript | ENST0000055353 | protein_coding | - | YES | MANE_Select |
| 12:56417805-5641 | - | upstream_gene_va | MODIFIER | - | ENSG0000027627 | Transcript | ENST0000061044 | lncRNA | - | YES | - |
| 12:56417805-5641 | - | upstream_gene_va | MODIFIER | - | ENSG0000027456 | Transcript | ENST0000062058 | unitary_pseudogene | - | YES | - |
| 12:89598780-8959 | - | intron_variant | MODIFIER | ATP2B1 | ENSG0000007096 | Transcript | ENST0000042867 | protein_coding | - | YES | MANE_Select |
| 12:95898502-9589 | - | intron_variant | MODIFIER | CCDC38 | ENSG0000016597 | Transcript | ENST0000034428 | protein_coding | rs60008366 | YES | MANE_Select |
| 12:113947257-113 | - | intron_variant | MODIFIER | RBM19 | ENSG0000012296 | Transcript | ENST0000026174 | protein_coding | rs369688809 | YES | MANE_Select |
| 13:77070573-7707 | - | intron_variant | MODIFIER | MYCBP2 | ENSG0000000581 | Transcript | ENST0000054444 | protein_coding | - | YES | MANE_Select |
| 13:99982753-9998 | - | upstream_gene_va | MODIFIER | - | ENSG0000028806 | Transcript | ENST0000065377 | lncRNA | - | YES | - |
| 13:101090060-101 | - | intron_variant | MODIFIER | NALCN | ENSG0000010245 | Transcript | ENST0000025112 | protein_coding | rs148372023 | YES | MANE_Select |
| 14:24163922-2416 | - | downstream_gene_ | MODIFIER | RNF31 | ENSG0000009209 | Transcript | ENST0000032410 | protein_coding | - | YES | MANE_Select |
| 14:24163922-2416 | - | downstream_gene_ | MODIFIER | - | ENSG0000028975 | Transcript | ENST0000069902 | lncRNA | - | YES | - |
| 14:24300643-2430 | - | upstream_gene_va | MODIFIER | DHRS1 | ENSG0000015737 | Transcript | ENST0000028811 | protein_coding | - | YES | MANE_Select |
| 14:24300643-2430 | - | downstream_gene_ | MODIFIER | CIDEB | ENSG0000013630 | Transcript | ENST0000055441 | protein_coding | - | YES | MANE_Select |
| 14:24300643-2430 | - | downstream_gene_ | MODIFIER | - | ENSG0000028804 | Transcript | ENST0000073790 | lncRNA | - | YES | - |

| | | | | | | | | | | | |
|---|---|---|---|---|---|---|---|---|---|---|---|
| 14:61457374-6145 | - | intron_variant | MODIFIER | PRKCH | ENSG0000002707 | Transcript | ENST0000033298 | protein_coding | - | YES | MANE_Select |
| 15:40036395-4003 | - | downstream_gene_ | MODIFIER | EIF2AK4 | ENSG0000012882 | Transcript | ENST0000026379 | protein_coding | - | YES | MANE_Select |
| 15:40036395-4003 | - | upstream_gene_va | MODIFIER | SRP14-DT | ENSG0000024850 | Transcript | ENST0000074667 | lncRNA | - | YES | - |
| 15:53523330-5352 | - | splice_polypyrimi | LOW | WDR72 | ENSG0000016641 | Transcript | ENST0000036050 | protein_coding | rs57737580 | YES | MANE_Select |
| 15:53523330-5352 | - | intron_variant,non | MODIFIER | - | ENSG0000028759 | Transcript | ENST0000085101 | lncRNA | rs57737580 | YES | - |
| 15:73288149-7328 | - | intron_variant | MODIFIER | NEO1 | ENSG0000006714 | Transcript | ENST0000026190 | protein_coding | rs150219014 | YES | MANE_Select |
| 16:3284207-32842 | - | splice_donor_5th_ | LOW | ZNF263 | ENSG0000000619 | Transcript | ENST0000021906 | protein_coding | - | YES | MANE_Select |
| 16:72788664-7278 | - | intron_variant,non | MODIFIER | ZFHX3-AS1 | ENSG0000025976 | Transcript | ENST0000076645 | lncRNA | rs376311468 | YES | - |
| 17:4809268-48092 | - | intron_variant | MODIFIER | PLD2 | ENSG0000012921 | Transcript | ENST0000026308 | protein_coding | rs59096986 | YES | MANE_Select |
| 17:8160737-81607 | - | 3_prime_UTR_var | MODIFIER | VAMP2 | ENSG0000022020 | Transcript | ENST0000031650 | protein_coding | - | YES | MANE_Select |
| 17:8160737-81607 | - | upstream_gene_va | MODIFIER | - | ENSG0000029995 | Transcript | ENST0000076770 | lncRNA | - | YES | - |
| 17:28584629-2858 | - | intron_variant | MODIFIER | SPAG5 | ENSG0000007638 | Transcript | ENST0000032176 | protein_coding | rs112114466 | YES | MANE_Select |
| 17:39745549-3974 | - | intron_variant | MODIFIER | GRB7 | ENSG0000014173 | Transcript | ENST0000030915 | protein_coding | rs67306131 | YES | MANE_Select |
| 17:42955992-4295 | - | intron_variant | MODIFIER | AARSD1 | ENSG0000026696 | Transcript | ENST0000042756 | protein_coding | - | YES | MANE_Select |
| 17:60069867-6006 | - | intron_variant | MODIFIER | HEATR6 | ENSG0000006809 | Transcript | ENST0000018495 | protein_coding | - | YES | MANE_Select |
| 19:1578372-15783 | - | downstream_gene_ | MODIFIER | - | ENSG0000027900 | Transcript | ENST0000062442 | TEC | - | YES | - |
| 19:2733503-27335 | - | intron_variant | MODIFIER | SLC39A3 | ENSG0000014187 | Transcript | ENST0000026974 | protein_coding | rs139950756 | YES | MANE_Select |
| 19:2733503-27335 | - | upstream_gene_va | MODIFIER | - | ENSG0000026134 | Transcript | ENST0000056790 | lncRNA | rs139950756 | YES | - |
| 19:20132594-2013 | - | splice_polypyrimi | LOW | - | ENSG0000026722 | Transcript | ENST0000059292 | transcribed_unprocessed_p | - | YES | - |
| 19:20132594-2013 | - | splice_polypyrimi | LOW | - | ENSG0000026738 | Transcript | ENST0000059365 | lncRNA | - | YES | - |
| 19:20132594-2013 | - | upstream_gene_va | MODIFIER | BNIP3P15 | ENSG0000027071 | Transcript | ENST0000060460 | processed_pseudogene | - | YES | - |
| 19:42739195-4273 | - | intron_variant | MODIFIER | PSG3 | ENSG0000022182 | Transcript | ENST0000032749 | protein_coding | - | YES | MANE_Select |
| 19:48836425-4883 | - | downstream_gene_ | MODIFIER | PLEKHA4 | ENSG0000010555 | Transcript | ENST0000026326 | protein_coding | - | YES | MANE_Select |
| 19:48836425-4883 | - | 5_prime_UTR_var | MODIFIER | HSD17B14 | ENSG0000008707 | Transcript | ENST0000026327 | protein_coding | - | YES | MANE_Select |
| 19:58557202-5855 | - | downstream_gene_ | MODIFIER | MZF1 | ENSG0000009932 | Transcript | ENST0000021505 | protein_coding | rs145976499 | YES | MANE_Select |
| 19:58557202-5855 | - | intron_variant | MODIFIER | UBE2M | ENSG0000013072 | Transcript | ENST0000025302 | protein_coding | rs145976499 | YES | MANE_Select |
| 19:58557202-5855 | - | upstream_gene_va | MODIFIER | CHMP2A | ENSG0000013072 | Transcript | ENST0000031254 | protein_coding | rs145976499 | YES | MANE_Select |
| 19:58557202-5855 | - | upstream_gene_va | MODIFIER | MZF1-AS1 | ENSG0000026785 | Transcript | ENST0000078983 | lncRNA | rs145976499 | YES | - |
| 2:37372476-37372 | - | splice_region_vari | LOW | QPCT | ENSG0000011582 | Transcript | ENST0000033841 | protein_coding | - | YES | MANE_Select |
| 2:37372476-37372 | - | downstream_gene_ | MODIFIER | - | ENSG0000029646 | Transcript | ENST0000073982 | lncRNA | - | YES | - |
| 2:222293705-2222 | - | intron_variant | MODIFIER | PAX3 | ENSG0000013590 | Transcript | ENST0000039207 | protein_coding | - | YES | MANE_Select |
| 2:222293705-2222 | - | upstream_gene_va | MODIFIER | CCDC140 | ENSG0000016308 | Transcript | ENST0000064776 | lncRNA | - | YES | - |
| 2:232378818-2323 | - | intron_variant,non | MODIFIER | ECEL1P2 | ENSG0000022451 | Transcript | ENST0000071529 | lncRNA | - | YES | - |
| 20:34743163-3474 | - | intron_variant,non | MODIFIER | - | ENSG0000028680 | Transcript | ENST0000065504 | lncRNA | - | YES | - |
| 20:47169101-4716 | - | intron_variant | MODIFIER | EYA2 | ENSG0000006465 | Transcript | ENST0000032761 | protein_coding | - | YES | MANE_Select |
| 20:47169101-4716 | - | downstream_gene_ | MODIFIER | MIR3616 | ENSG0000026490 | Transcript | ENST0000058407 | miRNA | - | YES | - |
| 20:56248628-5624 | - | upstream_gene_va | MODIFIER | MC3R | ENSG0000012408 | Transcript | ENST0000024391 | protein_coding | rs368604087 | YES | MANE_Select |
| 21:45917570-4591 | - | splice_polypyrimi | LOW | - | ENSG0000028060 | Transcript | ENST0000062623 | lncRNA | rs548673780 | YES | - |
| 21:45917570-4591 | - | splice_polypyrimi | LOW | PCBP3 | ENSG0000018357 | Transcript | ENST0000068168 | protein_coding | rs548673780 | YES | MANE_Select |
| 3:65375708-65375 | - | intron_variant | MODIFIER | MAGI1 | ENSG0000015127 | Transcript | ENST0000040293 | protein_coding | - | YES | MANE_Select |
| 3:65387174-65387 | - | intron_variant | MODIFIER | MAGI1 | ENSG0000015127 | Transcript | ENST0000040293 | protein_coding | - | YES | MANE_Select |
| 3:128573475-1285 | - | intron_variant,non | MODIFIER | LINC01565 | ENSG0000019868 | Transcript | ENST0000075324 | lncRNA | - | YES | - |

| | | | | | | | | | | | |
|---|---|---|---|---|---|---|---|---|---|---|---|
| 4:654753-654755 | - | intron_variant,non | MODIFIER | PDE6B-AS1 | ENSG0000024268 | Transcript | ENST0000046835 | lncRNA | rs144362048 | YES | - |
| 4:654753-654755 | - | intron_variant | MODIFIER | PDE6B | ENSG0000013325 | Transcript | ENST0000049651 | protein_coding | rs144362048 | YES | MANE_Select |
| 5:170080126-1700 | - | intron_variant | MODIFIER | DOCK2 | ENSG0000013451 | Transcript | ENST0000052090 | protein_coding | - | YES | MANE_Select |
| 6:132463932-1324 | - | intron_variant | MODIFIER | STX7 | ENSG0000007995 | Transcript | ENST0000036794 | protein_coding | - | YES | MANE_Select |
| 7:20140884-20140 | - | 3_prime_UTR_var | MODIFIER | MACC1 | ENSG0000018374 | Transcript | ENST0000040033 | protein_coding | - | YES | MANE_Select |
| 7:20140884-20140 | - | intron_variant,non | MODIFIER | MACC1-AS1 | ENSG0000022859 | Transcript | ENST0000073867 | lncRNA | - | YES | - |
| 7:29504721-29504 | - | intron_variant | MODIFIER | CHN2 | ENSG0000010606 | Transcript | ENST0000022279 | protein_coding | - | YES | MANE_Select |
| 7:38245284-38245 | - | intron_variant | MODIFIER | TRGC2 | ENSG0000022719 | Transcript | ENST0000043691 | TR_C_gene | - | YES | - |
| 7:47413908-47413 | - | intron_variant | MODIFIER | TNS3 | ENSG0000013620 | Transcript | ENST0000031116 | protein_coding | - | YES | MANE_Select |
| 7:96028144-96028 | - | intron_variant | MODIFIER | DYNC1I1 | ENSG0000015856 | Transcript | ENST0000044746 | protein_coding | - | YES | MANE_Select |
| 7:142474204-1424 | - | downstream_gene_ | MODIFIER | TRBV6-6 | ENSG0000021172 | Transcript | ENST0000039037 | TR_V_gene | - | YES | - |
| 7:142474204-1424 | - | intron_variant,non | MODIFIER | TRBV7-5 | ENSG0000022755 | Transcript | ENST0000039038 | TR_V_pseudogene | - | YES | - |
| 8:67161738-67161 | - | intron_variant | MODIFIER | CSPP1 | ENSG0000010421 | Transcript | ENST0000067861 | protein_coding | - | YES | MANE_Select |
| 8:140532162-1405 | - | splice_polypyrimic | LOW | AGO2 | ENSG0000012390 | Transcript | ENST0000022059 | protein_coding | - | YES | MANE_Select |
| 8:144317162-1443 | - | upstream_gene_va | MODIFIER | - | ENSG0000025469 | Transcript | ENST0000052502 | lncRNA | - | YES | - |
| 8:144317162-1443 | - | intron_variant | MODIFIER | DGAT1 | ENSG0000018500 | Transcript | ENST0000052871 | protein_coding | - | YES | MANE_Select |
| 8:144317162-1443 | - | downstream_gene_ | MODIFIER | HSF1 | ENSG0000018512 | Transcript | ENST0000052883 | protein_coding | - | YES | MANE_Select |
| 8:144317162-1443 | - | downstream_gene_ | MODIFIER | MIR6848 | ENSG0000028422 | Transcript | ENST0000061121 | miRNA | - | YES | - |
| 9:35825544-35825 | - | intron_variant | MODIFIER | FAM221B | ENSG0000020493 | Transcript | ENST0000042353 | protein_coding | - | YES | MANE_Select |
| 9:35825544-35825 | - | upstream_gene_va | MODIFIER | TMEM8B | ENSG0000013710 | Transcript | ENST0000064393 | protein_coding | - | YES | MANE_Select |
| 9:36923306-36923 | - | intron_variant | MODIFIER | PAX5 | ENSG0000019609 | Transcript | ENST0000035812 | protein_coding | rs768276588 | YES | MANE_Select |
| X:139958016-139 | - | intron_variant | MODIFIER | CXorf66 | ENSG0000020393 | Transcript | ENST0000037054 | protein_coding | - | YES | MANE_Select |
| **PAAD** | | | | | | | | | | | |
| 19:19833449-1983 | - | intron_variant,non | MODIFIER | ZNF56P | ENSG0000029117 | Transcript | ENST0000059188 | lncRNA | rs372149164 | YES | - |
| 19:19833449-1983 | - | upstream_gene_va | MODIFIER | ZNF56P | ENSG0000026741 | Transcript | ENST0000062341 | transcribed_unprocessed_p | rs372149164 | YES | - |
| 19:19833449-1983 | - | intron_variant,non | MODIFIER | - | ENSG0000029615 | Transcript | ENST0000073692 | lncRNA | rs372149164 | YES | - |
| **BLGG** | | | | | | | | | | | |
| 1:20857326-20857 | - | intron_variant | MODIFIER | EIF4G3 | ENSG0000007515 | Transcript | ENST0000060232 | protein_coding | rs145504996 | YES | MANE_Select |
| 1:210684141-2106 | - | splice_region_vari | LOW | KCNH1 | ENSG0000014347 | Transcript | ENST0000027175 | protein_coding | rs144706702 | YES | MANE_Select |
| 1:210684141-2106 | - | downstream_gene_ | MODIFIER | - | ENSG0000027933 | Transcript | ENST0000062514 | TEC | rs144706702 | YES | - |
| 1:225847138-2258 | - | downstream_gene_ | MODIFIER | EPHX1 | ENSG0000014381 | Transcript | ENST0000027216 | protein_coding | rs750876984 | YES | MANE_Select |
| 1:225847138-2258 | - | upstream_gene_va | MODIFIER | - | ENSG0000024286 | Transcript | ENST0000042433 | lncRNA | rs750876984 | YES | - |
| 10:50850592-5085 | - | intron_variant | MODIFIER | A1CF | ENSG0000014858 | Transcript | ENST0000037399 | protein_coding | rs58342464 | YES | MANE_Select |
| 10:69414880-6941 | - | intron_variant | MODIFIER | TACR2 | ENSG0000007507 | Transcript | ENST0000037330 | protein_coding | rs150956058 | YES | MANE_Select |
| 10:69414880-6941 | - | intron_variant,non | MODIFIER | - | ENSG0000029948 | Transcript | ENST0000076385 | lncRNA | rs150956058 | YES | - |
| 11:66850936-6685 | - | downstream_gene_ | MODIFIER | RCE1 | ENSG0000017365 | Transcript | ENST0000030965 | protein_coding | rs139065746 | YES | MANE_Select |
| 11:66850936-6685 | - | splice_polypyrimic | LOW | PC | ENSG0000017359 | Transcript | ENST0000039396 | protein_coding | rs139065746 | YES | MANE_Select |
| 11:75797346-7579 | - | intron_variant | MODIFIER | DGAT2 | ENSG0000006228 | Transcript | ENST0000022802 | protein_coding | rs149770132 | YES | MANE_Select |
| 11:75797346-7579 | - | intron_variant,non | MODIFIER | UVRAG-DT | ENSG0000029362 | Transcript | ENST0000071620 | lncRNA | rs149770132 | YES | - |
| 11:132436788-132 | - | splice_polypyrimic | LOW | OPCML | ENSG0000018371 | Transcript | ENST0000052438 | protein_coding | rs145479085 | YES | MANE_Select |
| 12:18381738-1838 | - | intron_variant | MODIFIER | PIK3C2G | ENSG0000013914 | Transcript | ENST0000053877 | protein_coding | rs761371594 | YES | MANE_Select |
| 16:72788664-7278 | - | intron_variant,non | MODIFIER | ZFHX3-AS1 | ENSG0000025976 | Transcript | ENST0000076645 | lncRNA | rs376311468 | YES | - |

| | | | | | | | | | | | |
|---|---|---|---|---|---|---|---|---|---|---|---|
| 16:72797458-7279 | - | intron_variant,non | MODIFIER | ZFHX3-AS1 | ENSG0000025976 | Transcript | ENST0000076645 | lncRNA | rs34918837 | YES | - |
| 17:4809268-48092 | - | intron_variant | MODIFIER | PLD2 | ENSG0000012921 | Transcript | ENST0000026308 | protein_coding | rs59096986 | YES | MANE_Select |
| 17:28584629-2858 | - | intron_variant | MODIFIER | SPAG5 | ENSG0000007638 | Transcript | ENST0000032176 | protein_coding | rs112114466 | YES | MANE_Select |
| 19:2733503-27335 | - | intron_variant | MODIFIER | SLC39A3 | ENSG0000014187 | Transcript | ENST0000026974 | protein_coding | rs139950756 | YES | MANE_Select |
| 19:2733503-27335 | - | upstream_gene_va | MODIFIER | - | ENSG0000026134 | Transcript | ENST0000056790 | lncRNA | rs139950756 | YES | - |
| 19:5239100-52391 | - | intron_variant | MODIFIER | PTPRS | ENSG0000010542 | Transcript | ENST0000026296 | protein_coding | rs149487300 | YES | MANE_Select |
| 19:6531137-65311 | - | downstream_gene_ | MODIFIER | - | ENSG0000029750 | Transcript | ENST0000074847 | lncRNA | rs564151103 | YES | - |
| 19:7678296-76782 | - | intron_variant | MODIFIER | MCEMP1 | ENSG0000018301 | Transcript | ENST0000033359 | protein_coding | rs151233009 | YES | MANE_Select |
| 19:7678296-76782 | - | upstream_gene_va | MODIFIER | TRAPPC5 | ENSG0000018102 | Transcript | ENST0000059614 | protein_coding | rs151233009 | YES | MANE_Select |
| 19:19833449-1983 | - | intron_variant,non | MODIFIER | ZNF56P | ENSG0000029117 | Transcript | ENST0000059188 | lncRNA | rs372149164 | YES | - |
| 19:19833449-1983 | - | upstream_gene_va | MODIFIER | ZNF56P | ENSG0000026741 | Transcript | ENST0000062341 | transcribed_unprocessed_p | rs372149164 | YES | - |
| 19:19833449-1983 | - | intron_variant,non | MODIFIER | - | ENSG0000029615 | Transcript | ENST0000073692 | lncRNA | rs372149164 | YES | - |
| 19:20132594-2013 | - | splice_region_vari | LOW | - | ENSG0000026722 | Transcript | ENST0000059292 | transcribed_unprocessed_p | rs1184014518 | YES | - |
| 19:20132594-2013 | - | splice_region_vari | LOW | - | ENSG0000026738 | Transcript | ENST0000059365 | lncRNA | rs1184014518 | YES | - |
| 19:20132594-2013 | - | upstream_gene_va | MODIFIER | BNIP3P15 | ENSG0000027071 | Transcript | ENST0000060460 | processed_pseudogene | rs1184014518 | YES | - |
| 19:20184331-2018 | - | intron_variant | MODIFIER | ZNF486 | ENSG0000025622 | Transcript | ENST0000033511 | protein_coding | - | YES | MANE_Select |
| 19:20184331-2018 | - | intron_variant,non | MODIFIER | - | ENSG0000026738 | Transcript | ENST0000059365 | lncRNA | - | YES | - |
| 19:21405088-2140 | - | intron_variant | MODIFIER | ZNF493 | ENSG0000019626 | Transcript | ENST0000039228 | protein_coding | rs35143973 | YES | MANE_Select |
| 19:38408861-3840 | - | downstream_gene_ | MODIFIER | RASGRP4 | ENSG0000017177 | Transcript | ENST0000061543 | protein_coding | rs59917662 | YES | MANE_Select |
| 2:11133310-11133 | - | 5_prime_UTR_var | MODIFIER | CIMIP5 | ENSG0000015087 | Transcript | ENST0000038158 | protein_coding | - | YES | MANE_Select |
| 2:11133310-11133 | - | upstream_gene_va | MODIFIER | - | ENSG0000014506 | Transcript | ENST0000054430 | lncRNA | - | YES | - |
| 2:11133310-11133 | - | downstream_gene_ | MODIFIER | - | ENSG0000029616 | Transcript | ENST0000073702 | lncRNA | - | YES | - |
| 2:37372476-37372 | - | splice_donor_5th_ | LOW | QPCT | ENSG0000011582 | Transcript | ENST0000033841 | protein_coding | rs112031803 | YES | MANE_Select |
| 2:37372476-37372 | - | downstream_gene_ | MODIFIER | - | ENSG0000029646 | Transcript | ENST0000073982 | lncRNA | rs112031803 | YES | - |
| 2:69193442-69193 | - | intron_variant | MODIFIER | ANTXR1 | ENSG0000016960 | Transcript | ENST0000030371 | protein_coding | rs10627389 | YES | MANE_Select |
| 20:56248628-5624 | - | upstream_gene_va | MODIFIER | MC3R | ENSG0000012408 | Transcript | ENST0000024391 | protein_coding | rs368604087 | YES | MANE_Select |
| 21:45917570-4591 | - | splice_polypyrimic | LOW | - | ENSG0000028060 | Transcript | ENST0000062623 | lncRNA | rs548673780 | YES | - |
| 21:45917570-4591 | - | splice_polypyrimic | LOW | PCBP3 | ENSG0000018357 | Transcript | ENST0000068168 | protein_coding | rs548673780 | YES | MANE_Select |
| 3:158102929-1581 | - | intron_variant | MODIFIER | SHOX2 | ENSG0000016877 | Transcript | ENST0000048385 | protein_coding | rs374383282 | YES | MANE_Select |
| 3:187253326-1872 | - | splice_polypyrimic | LOW | MASP1 | ENSG0000012724 | Transcript | ENST0000029628 | protein_coding | - | YES | MANE_Select |
| 4:654753-654757 | - | intron_variant,non | MODIFIER | PDE6B-AS1 | ENSG0000024268 | Transcript | ENST0000046835 | lncRNA | - | YES | - |
| 4:654753-654757 | - | intron_variant | MODIFIER | PDE6B | ENSG0000013325 | Transcript | ENST0000049651 | protein_coding | - | YES | MANE_Select |
| 6:112141268-1121 | - | intron_variant | MODIFIER | LAMA4 | ENSG0000011276 | Transcript | ENST0000023053 | protein_coding | rs149046391 | YES | MANE_Select |
| 6:112141268-1121 | - | intron_variant,non | MODIFIER | - | ENSG0000023723 | Transcript | ENST0000078458 | lncRNA | rs149046391 | YES | - |
| 6:133889359-1338 | - | 5_prime_UTR_var | MODIFIER | TCF21 | ENSG0000011852 | Transcript | ENST0000036788 | protein_coding | rs3068172 | YES | MANE_Select |
| 6:133889359-1338 | - | upstream_gene_va | MODIFIER | TARID | ENSG0000022795 | Transcript | ENST0000079540 | lncRNA | rs3068172 | YES | - |
| 6:144700057-1447 | - | intron_variant | MODIFIER | UTRN | ENSG0000015281 | Transcript | ENST0000036754 | protein_coding | - | YES | MANE_Select |
| 6:151458285-1514 | - | intron_variant | MODIFIER | ARMT1 | ENSG0000014647 | Transcript | ENST0000036729 | protein_coding | rs142167376 | YES | MANE_Select |
| 7:2945755-294575 | - | intron_variant | MODIFIER | CARD11 | ENSG0000019828 | Transcript | ENST0000039694 | protein_coding | rs779700969 | YES | MANE_Select |
| 7:2945755-294575 | - | intron_variant,non | MODIFIER | CARD11-AS1 | ENSG0000023728 | Transcript | ENST0000081660 | lncRNA | rs779700969 | YES | - |
| 7:38242529-38242 | - | intron_variant | MODIFIER | TRGC2 | ENSG0000022719 | Transcript | ENST0000043691 | TR_C_gene | rs35917252 | YES | - |
| 7:38245284-38245 | - | intron_variant | MODIFIER | TRGC2 | ENSG0000022719 | Transcript | ENST0000043691 | TR_C_gene | rs138091320 | YES | - |

| | | | | | | | | | | | |
|---|---|---|---|---|---|---|---|---|---|---|---|
| 7:65387877-65387 | - | intron_variant | MODIFIER | ZNF92 | ENSG0000014675 | Transcript | ENST0000032874 | protein_coding | rs143107154 | YES | MANE_Select |
| 7:66951150-66951 | - | intron_variant | MODIFIER | TMEM248 | ENSG0000010660 | Transcript | ENST0000034156 | protein_coding | rs148803801 | YES | MANE_Select |
| 7:66951150-66951 | - | intron_variant,non_ | MODIFIER | - | ENSG0000029976 | Transcript | ENST0000076623 | lncRNA | rs148803801 | YES | - |
| 7:130041131-1300 | - | intron_variant | MODIFIER | ZC3HC1 | ENSG0000009173 | Transcript | ENST0000035830 | protein_coding | rs375888799 | YES | MANE_Select |
| 9:35825544-35825 | - | intron_variant | MODIFIER | FAM221B | ENSG0000020493 | Transcript | ENST0000042353 | protein_coding | rs201802596 | YES | MANE_Select |
| 9:35825544-35825 | - | upstream_gene_va | MODIFIER | TMEM8B | ENSG0000013710 | Transcript | ENST0000064393 | protein_coding | rs201802596 | YES | MANE_Select |
| **LUAD** | | | | | | | | | | | |
| 19:20184331-2018 | - | intron_variant | MODIFIER | ZNF486 | ENSG0000025622 | Transcript | ENST0000033511 | protein_coding | rs367731486 | YES | MANE_Select |
| 19:20184331-2018 | - | intron_variant,non_ | MODIFIER | - | ENSG0000026738 | Transcript | ENST0000059365 | lncRNA | rs367731486 | YES | - |
| **HNSC** | | | | | | | | | | | |
| 19:20646412-2064 | - | splice_polypyrimid | LOW | ZNF626 | ENSG0000018817 | Transcript | ENST0000060144 | protein_coding | - | YES | MANE_Select |
