## Supplementary table_3 for "Direct Pan-Cancer Multi-Omic Analysis of Radiotherapy Response Reveals Coding and Regulatory Genomic Variation, and Extracellular Adaptive Programs Underlying Radioresistance"

| Supplementary Table 3: Shared variants with odds ratio association for both RR and RS | | | | | |
|---|---|---|---|---|---|
| **Variant** | **rs ID** | **Gene Symbol** | **RR** | **RS** | **Consequence** |
| High Impact | | High Impact | | | |
| chr10_128106847_CTG_C/ chr10_128106847_CTGTG_CTG | rs145960091 | MKI67 | HNSC, LGG | HNSC, LGG | Frameshift variant |
| chr16_50334766_CCT_C/ chr16_50334766_CCTCT_C/ chr16_50334766_CCTCT_CCT | rs145896392 | BRD7 | PRAD, LGG, Adenocarcinoma | LGG, Adenocarcinoma | Frameshift variant |
| chr3_65387174_TTC_T/ chr3_65387174_TTCTC_TTC | -, rs139764373 | MAGI1 | Adenocarcinoma | Adenocarcinoma | Frameshift variant |
| chr4_53453080_CAGAG_C/ chr4_53453080_CAGAG_CAG/ chr4_53453080_CAG_C | rs143671659 | FIP1L1 | Adenocarcinoma | LGG, Adenocarcinoma | Frameshift variant |
| Moderate Impact | | Moderate Impact | | | |
| chr6_161098318_CCTGCTGCTG_C | rs5881391 | MAP3K4 | HNSC | LGG | Inframe deletion |
| chr1_225847138_TCTGCTG_TCTG/ chr1_225847138_TCTG_T | rs750876984 | TMEM63A | LGG | LGG | Inframe deletion |
| chr19_38408861_CAAG_C/ chr19_38408861_CAAG**AAG**_C**AAG** | rs59917662 | FAM98C | LGG | LGG | Inframe deletion |
| chr16_51141744_CGCT_C | rs113614842 | SALL1 | Adenocarcinoma | LGG | Inframe deletion |
| chr16_72788664_TTGC_T/ chr16_72788664_TTGCTGC_TTGC | rs376311468 | ZFHX3 | Adenocarcinoma | LGG, Adenocarcinoma | Inframe deletion |
| chr16_72797458_CTTG_C/ chr16_72797458_CTTGTTG_CTTG | rs34918837 | ZFHX3 | Adenocarcinoma | LGG | Inframe deletion |
| chr8_28351708_GGCAGCA_GGCA/ chr8_28351708_GGCA_G | rs547945591 | ZNF395 | LGG, Adenocarcinoma | LGG, Adenocarcinoma | Inframe deletion |
| chr3_65439885_TCTG_T/ chr3_65439885_TCTGCTG_TCTG | rs142043619 | MAGI1 | Adenocarcinoma | LGG, Adenocarcinoma | Inframe deletion |
| chr19_1578372_GCTCCTC_GCTC/ chr19_1578372_GCTC_G | rs371220154 | MBD3 | Adenocarcinoma | Adenocarcinoma | Inframe deletion |
| chr2_232378818_ATGC_A/ chr2_232378818_ATGCTGC_ATGC | rs377162921 | ALPP | Adenocarcinoma | Adenocarcinoma | Inframe deletion |
| chr20_23365283_AGAG_A/ chr20_23365283_AGAGGAG_AGAG | rs750609719 | GZF1 | Adenocarcinoma | Adenocarcinoma | Inframe deletion |
| chr20_34743163_TTGC_T/ chr20_34743163_TTGCTGC_TTGC | rs112051697 | NCOA6 | Adenocarcinoma | Adenocarcinoma | Inframe deletion |
| chr20_47238822_TTGC_T/ chr20_47238822_TTGCTGC_TTGC | rs569370037 | ZMYND8 | Adenocarcinoma | Adenocarcinoma | Inframe deletion |
| chr10_30027571_ACTG_A/ chr10_30027571_ACTGCTG_ACTG | rs34594193 | JCAD | Adenocarcinoma | Adenocarcinoma | Inframe deletion |
| chr11_115209591_ATGGTGG_ATGG/ chr11_115209591_ATGGTGGTGGTGGT | - | CADM1 | Adenocarcinoma | Adenocarcinoma | Inframe deletion |
| chr12_8047961_ACAG_A/ chr12_8047961_ACAGCAG_ACAG | rs372118289 | FOXJ2 | Adenocarcinoma | Adenocarcinoma | Inframe deletion |
| chr13_99982753_CCCACCACCACCAC( chr13_99982753_CCCA_C/ chr13_99982753_CCCACCA_CCCA | rs398124241 | ZIC2 | HNSC, Adenocarcinom | Adenocarcinoma | Inframe deletion |
| chr14_24163922_GAGCAGC_GAGC/ chr14_24163922_GAGC_G | rs746378882 | IRF9 | Adenocarcinoma | Adenocarcinoma | Inframe deletion |
| chr14_24300643_AGAGGAG_AGAG/ chr14_24300643_AGAG_A | rs71119069 | NOP9 | Adenocarcinoma | Adenocarcinoma | Inframe deletion |
| chr15_40036395_GTGCTGC_GTGC/ chr15_40036395_GTGC_G | rs371085676 | SRP14 | Adenocarcinoma | Adenocarcinoma | Inframe deletion |
| chr1_6469122_TTCCTCC_TTCC/ chr1_6469122_TTCC_T/ chr1_6469122_TTCCTCCTCC_TTCCTC | rs113541584 | PLEKHG5 | HNSC, Adenocarcinom | Adenocarcinoma | Inframe deletion |
