## Supplementary table_4 for "Direct Pan-Cancer Multi-Omic Analysis of Radiotherapy Response Reveals Coding and Regulatory Genomic Variation, and Extracellular Adaptive Programs Underlying Radioresistance"

| Supplementary Figure 2: Unique variants with odds ratio association for either RR and RS | | | | | |
|---|---|---|---|---|---|
| **Supplementary Figure 2: Unique Variants** | | | | | |
| **Variant** | **rs ID** | **Gene Symbol** | **RR** | **RS** | **Consequence** |
| High Impact | | | | | |
| chr9_33795614_G_G | rs1188045977 | PRSS3 | Adenocarcinoma | | Splice donor variant |
| chr9_130668313_GGT_G | rs138789124 | PRDM12 | | LGG | Splice donor variant |
| chr14_73105898_AAG_A | rs150988201 | RBM25 | HNSC | | Frameshift variant |
| chr2_226796679_TTGCTGC_TTGC | CD951753 | IRS1 | HNSC | | Frameshift variant |
| Moderate Impact | | | | | |
| chr19_6531137_GGCT_G | rs564151103 | TNFSF9 | | LGG | Inframe deletion |
| chr9_117214709_CCAG_C | rs750505669 | ASTN2 | HNSC | | Inframe deletion |
| chrX_151648669_CGCT_C | rs750036801 | PASD1 | HNSC | | Inframe deletion |
| chr17_40822513_ATCC_A | rs148510452 | KRT10 | HNSC | | Inframe deletion |
| chr22_42214566_AAGGAGG_AAGG | - | TCF20 | | Adenocarcinoma | Inframe deletion |
| chr1_214383705_GCCT_G | rs143136196 | PTPN14 | | LGG | Inframe deletion |
| chr1_20857326_TTGTG_TTG | - | PTPN14 | LGG | | Inframe deletion |
