## Supplementary table_5 for "Direct Pan-Cancer Multi-Omic Analysis of Radiotherapy Response Reveals Coding and Regulatory Genomic Variation, and Extracellular Adaptive Programs Underlying Radioresistance"

### Transcriptomics results

#### List of genes differentially expressed between RR and RS across different cancer types

| Genes with adj.p-val < 0.05 | | | | | | | | | | | | | | | | |
|---|---|---|---|---|---|---|---|---|---|---|---|---|---|---|---|---|
| **TCGA-Project** | **source** | **type** | **score** | **phase** | **gene_id** | **gene_type** | **gene_name** | **level** | **hgnc_id** | **havana_gene** | **logFC** | **AveExpr** | **t** | **P.Value** | **adj.P.Val** | **B** |
| STAD | HAVANA | gene | NA | NA | ENSG00000151632.17 | protein_coding | **AKR1C2** | 2 | HGNC:385 | OTTHUMG00000 | 3.564604 | 2.385178 | 7.08E+00 | 2.94E-09 | **8.14E-05** | 10.71411 |
| UCEC | HAVANA | gene | NA | NA | ENSG00000235590.7 | lncRNA | **GNAS-AS1** | 2 | HGNC:24872 | OTTHUMG00000 | 3.515229 | -3.72039 | 5.91E+00 | 1.96E-07 | **0.004913778** | 4.159048 |
| UCEC | HAVANA | gene | NA | NA | ENSG00000229921.7 | lncRNA | **KIF25-AS1** | 2 | HGNC:20953 | OTTHUMG00000 | 4.951964 | -3.34982 | 5.72E+00 | 4.01E-07 | **0.005024474** | 4.154043 |
| BRCA | HAVANA | gene | NA | NA | ENSG00000168306.13 | protein_coding | **ACOX2** | 1 | HGNC:120 | OTTHUMG00000 | 2.490543 | 3.329929 | 5.34E+00 | 2.60E-07 | **0.007142095** | -2.02363 |
| STAD | HAVANA | gene | NA | NA | ENSG00000225670.4 | lncRNA | **CADM3-AS1** | 2 | HGNC:40812 | OTTHUMG00000 | 3.25448 | -3.06889 | 5.53E+00 | 9.47E-07 | **0.013102924** | 4.814164 |
| UCEC | HAVANA | gene | NA | NA | ENSG00000118137.10 | protein_coding | **APOA1** | 1 | HGNC:600 | OTTHUMG00000 | 4.608375 | -0.10271 | 5.28E+00 | 2.02E-06 | **0.016886763** | 4.168318 |
| STAD | HAVANA | gene | NA | NA | ENSG00000187134.14 | protein_coding | **AKR1C1** | 2 | HGNC:384 | OTTHUMG00000 | 2.424585 | 3.345921 | 5.34E+00 | 1.89E-06 | **0.017468576** | 4.864438 |
| STAD | HAVANA | gene | NA | NA | ENSG00000172348.15 | protein_coding | **RCAN2** | 2 | HGNC:3041 | OTTHUMG00000 | 1.509342 | 3.871317 | 5.13E+00 | 3.99E-06 | **0.022163304** | 4.186773 |
| STAD | HAVANA | gene | NA | NA | ENSG00000213088.12 | protein_coding | **ACKR1** | 2 | HGNC:4035 | OTTHUMG00000 | 2.779456 | 2.372859 | 5.13E+00 | 4.01E-06 | **0.022163304** | 4.152167 |
| STAD | HAVANA | gene | NA | NA | ENSG00000211649.3 | IG_V_gene | **IGLV7-46** | 2 | HGNC:5930 | OTTHUMG00000 | 2.861934 | 1.280942 | 5.01E+00 | 6.20E-06 | **0.024951848** | 3.705087 |
| STAD | HAVANA | gene | NA | NA | ENSG00000236466.2 | lncRNA | **AL035670.1** | 2 | NA | OTTHUMG00000 | 2.439607 | -4.26001 | 5.00E+00 | 6.31E-06 | **0.024951848** | 2.994317 |
| CESC | HAVANA | gene | NA | NA | ENSG00000109472.14 | protein_coding | **CPE** | 2 | HGNC:2303 | OTTHUMG00000 | 1.97879 | 4.613644 | 5.16E+00 | 1.18E-06 | **0.027492124** | 4.247326 |
| STAD | HAVANA | gene | NA | NA | ENSG00000099260.11 | protein_coding | **PALMD** | 2 | HGNC:15846 | OTTHUMG00000 | 1.505866 | 2.813637 | 4.93E+00 | 8.21E-06 | **0.028381627** | 3.50057 |
| STAD | HAVANA | gene | NA | NA | ENSG00000174640.15 | protein_coding | **SLCO2A1** | 1 | HGNC:10955 | OTTHUMG00000 | 1.675547 | 5.182119 | 4.89E+00 | 9.42E-06 | **0.028962381** | 3.390811 |
| THCA | HAVANA | gene | NA | NA | ENSG00000156076.10 | protein_coding | **WIF1** | 2 | HGNC:18081 | OTTHUMG00000 | 2.732169 | -4.79311 | 4.81E+00 | 2.98E-06 | **0.038428847** | -3.50463 |
| THCA | HAVANA | gene | NA | NA | ENSG00000166961.15 | protein_coding | **MS4A15** | 2 | HGNC:28573 | OTTHUMG00000 | 2.415004 | -4.06247 | 4.81E+00 | 2.99E-06 | **0.038428847** | -3.39811 |
| CESC | HAVANA | gene | NA | NA | ENSG00000164761.9 | protein_coding | **TNFRSF11B** | 2 | HGNC:11909 | OTTHUMG00000 | 2.296542 | -0.13678 | 4.88E+00 | 3.88E-06 | **0.045168779** | 1.125061 |
| Genes with adj.p-val < 0.1 | | | | | | | | | | | | | | | | |
| HNSC | HAVANA | gene | NA | NA | ENSG00000250041.3 | lncRNA | AC069360.1 | 2 | NA | OTTHUMG00000 | 1.936227 | -0.56352 | 4.92E+00 | 2.08E-06 | 0.050942074 | 3.488976 |
| STAD | HAVANA | gene | NA | NA | ENSG00000280048.1 | TEC | AC012409.5 | 2 | NA | OTTHUMG00000 | 2.222888 | -2.8373 | 4.69E+00 | 1.87E-05 | 0.051709033 | 2.419806 |
| STAD | HAVANA | gene | NA | NA | ENSG00000273771.1 | lncRNA | AC024337.2 | 2 | NA | OTTHUMG00000 | 1.94158 | -3.47948 | 4.66E+00 | 2.08E-05 | 0.05229886 | 2.225686 |
| STAD | HAVANA | gene | NA | NA | ENSG00000280339.1 | TEC | AP001528.2 | 2 | NA | OTTHUMG00000 | 1.548618 | 0.025784 | 4.63E+00 | 2.30E-05 | 0.05262824 | 2.440985 |
| STAD | HAVANA | gene | NA | NA | ENSG00000211937.3 | IG_V_gene | IGHV2-5 | 2 | HGNC:5576 | OTTHUMG00000 | 2.782551 | 2.577033 | 4.54E+00 | 3.12E-05 | 0.05262824 | 2.29589 |
| STAD | HAVANA | gene | NA | NA | ENSG00000163145.13 | protein_coding | C1QTNF7 | 1 | HGNC:14342 | OTTHUMG00000 | 2.089001 | 0.84539 | 4.54E+00 | 3.11E-05 | 0.05262824 | 2.234075 |
| STAD | HAVANA | gene | NA | NA | ENSG00000156486.8 | protein_coding | KCNS2 | 2 | HGNC:6301 | OTTHUMG00000 | 2.110774 | -2.41354 | 4.61E+00 | 2.50E-05 | 0.05262824 | 2.225174 |
| STAD | HAVANA | gene | NA | NA | ENSG00000138347.16 | protein_coding | MYPN | 2 | HGNC:23246 | OTTHUMG00000 | 2.652767 | -1.78199 | 4.58E+00 | 2.75E-05 | 0.05262824 | 2.202252 |
| STAD | HAVANA | gene | NA | NA | ENSG00000274997.2 | protein_coding | H2AC12 | 2 | HGNC:13671 | OTTHUMG00000 | -1.75485 | -2.92039 | -4.51E+00 | 3.55E-05 | 0.05262824 | 1.532354 |
| STAD | HAVANA | gene | NA | NA | ENSG00000109906.14 | protein_coding | ZBTB16 | 2 | HGNC:12930 | OTTHUMG00000 | 2.243217 | 1.903204 | 4.45E+00 | 4.34E-05 | 0.054515079 | 1.984716 |
| STAD | HAVANA | gene | NA | NA | ENSG00000196616.14 | protein_coding | ADH1B | 2 | HGNC:250 | OTTHUMG00000 | 3.348646 | 1.722373 | 4.44E+00 | 4.49E-05 | 0.054515079 | 1.961449 |
| STAD | HAVANA | gene | NA | NA | ENSG00000271178.1 | IG_V_gene | IGHV3OR16-13 | 2 | HGNC:5637 | OTTHUMG00000 | 2.657086 | -2.36108 | 4.41E+00 | 5.01E-05 | 0.056645607 | 1.656905 |
| STAD | HAVANA | gene | NA | NA | ENSG00000174175.17 | protein_coding | SELP | 2 | HGNC:10721 | OTTHUMG00000 | 2.255939 | 2.118581 | 4.37E+00 | 5.69E-05 | 0.056671685 | 1.74631 |
| STAD | HAVANA | gene | NA | NA | ENSG00000280304.1 | TEC | AC104260.2 | 2 | NA | OTTHUMG00000 | 1.857101 | -2.36108 | 4.37E+00 | 5.74E-05 | 0.056671685 | 1.540263 |
| STAD | HAVANA | gene | NA | NA | ENSG00000112964.14 | protein_coding | GHR | 2 | HGNC:4263 | OTTHUMG00000 | 1.6515 | 1.049557 | 4.35E+00 | 6.11E-05 | 0.057470299 | 1.645438 |
| STAD | HAVANA | gene | NA | NA | ENSG00000244116.3 | IG_V_gene | IGKV2-28 | 2 | HGNC:5783 | OTTHUMG00000 | 2.535944 | -1.49405 | 4.34E+00 | 6.25E-05 | 0.057470299 | 1.530386 |
| STAD | HAVANA | gene | NA | NA | ENSG00000288547.3 | lncRNA | C3orf36 | 2 | HGNC:26170 | OTTHUMG00000 | 1.516281 | -0.51704 | 4.31E+00 | 6.82E-05 | 0.057470299 | 1.484766 |
| STAD | HAVANA | gene | NA | NA | ENSG00000242534.2 | IG_V_gene | IGKV2D-28 | 2 | HGNC:5799 | OTTHUMG00000 | 2.390306 | -1.54485 | 4.31E+00 | 6.86E-05 | 0.057470299 | 1.448114 |
| STAD | HAVANA | gene | NA | NA | ENSG00000164161.10 | protein_coding | HHIP | 2 | HGNC:14866 | OTTHUMG00000 | 2.085949 | 1.262297 | 4.29E+00 | 7.35E-05 | 0.05831034 | 1.497853 |
| STAD | HAVANA | gene | NA | NA | ENSG00000172724.12 | protein_coding | CCL19 | 2 | HGNC:10617 | OTTHUMG00000 | 3.020425 | 0.8577 | 4.28E+00 | 7.62E-05 | 0.05831034 | 1.468781 |
| STAD | HAVANA | gene | NA | NA | ENSG00000163792.6 | protein_coding | TCF23 | 2 | HGNC:18602 | OTTHUMG00000 | 2.329108 | -2.76112 | 4.27E+00 | 7.95E-05 | 0.05831034 | 1.244409 |
| STAD | HAVANA | gene | NA | NA | ENSG00000152092.16 | protein_coding | ASTN1 | 1 | HGNC:773 | OTTHUMG00000 | 2.808182 | -2.70495 | 4.26E+00 | 8.22E-05 | 0.05831034 | 1.222682 |
| STAD | HAVANA | gene | NA | NA | ENSG00000165799.5 | protein_coding | RNASE7 | 2 | HGNC:19278 | OTTHUMG00000 | 2.516047 | -3.82205 | 4.28E+00 | 7.57E-05 | 0.05831034 | 1.167885 |
| STAD | HAVANA | gene | NA | NA | ENSG00000168079.17 | protein_coding | SCARA5 | 2 | HGNC:28701 | OTTHUMG00000 | 2.597997 | 0.744004 | 4.22E+00 | 9.35E-05 | 0.060572207 | 1.279327 |
| STAD | HAVANA | gene | NA | NA | ENSG00000134533.6 | protein_coding | RERG | 1 | HGNC:15980 | OTTHUMG00000 | 1.695207 | 2.115366 | 4.191135 | 0.0001029 | 0.060572207 | 1.213933 |
| STAD | HAVANA | gene | NA | NA | ENSG00000169252.6 | protein_coding | ADRB2 | 2 | HGNC:286 | OTTHUMG00000 | 1.708278 | -0.11922 | 4.20E+00 | 0.0001001 | 0.060572207 | 1.176281 |
| STAD | HAVANA | gene | NA | NA | ENSG00000108231.14 | protein_coding | LGI1 | 2 | HGNC:6572 | OTTHUMG00000 | 2.250999 | -3.09626 | 4.206503 | 9.78E-05 | 0.060572207 | 1.043978 |
| STAD | HAVANA | gene | NA | NA | ENSG00000287109.1 | lncRNA | AP000224.1 | 2 | NA | OTTHUMG00000 | 1.847126 | -3.35794 | 4.20E+00 | 0.0001008 | 0.060572207 | 0.980621 |
| STAD | HAVANA | gene | NA | NA | ENSG00000240654.6 | protein_coding | C1QTNF9 | 2 | HGNC:28732 | OTTHUMG00000 | 1.793881 | -4.30079 | 4.225264 | 9.19E-05 | 0.060572207 | 0.881456 |

| | | | | | | | | | | | | | | | | |
|---|---|---|---|---|---|---|---|---|---|---|---|---|---|---|---|---|
| STAD | HAVANA | gene | NA | NA | ENSG00000277631.5 | lncRNA | PGM5P3-AS1 | 2 | HGNC:51194 | OTTHUMG00000 | 1.986142 | -3.89729 | 4.179441 | 0.000107 | 0.061660909 | 0.85497 |
| STAD | HAVANA | gene | NA | NA | ENSG00000154721.15 | protein_coding | JAM2 | 2 | HGNC:14686 | OTTHUMG00000 | 1.502944 | 3.069214 | 4.141032 | 0.0001215 | 0.062429784 | 1.065054 |
| STAD | HAVANA | gene | NA | NA | ENSG00000141338.14 | protein_coding | ABCA8 | 2 | HGNC:38 | OTTHUMG00000 | 2.366091 | 1.615156 | 4.125036 | 0.0001281 | 0.062429784 | 1.017294 |
| STAD | HAVANA | gene | NA | NA | ENSG00000153162.9 | protein_coding | BMP6 | 2 | HGNC:1073 | OTTHUMG00000 | 1.509554 | 1.10944 | 4.130712 | 0.0001257 | 0.062429784 | 1.014272 |
| STAD | HAVANA | gene | NA | NA | ENSG00000019991.18 | protein_coding | HGF | 2 | HGNC:4893 | OTTHUMG00000 | 1.524117 | 2.39995 | 4.119021 | 0.0001307 | 0.062429784 | 1.002485 |
| STAD | HAVANA | gene | NA | NA | ENSG00000162687.19 | protein_coding | KCNT2 | 2 | HGNC:18866 | OTTHUMG00000 | 1.539676 | -0.26464 | 4.13E+00 | 0.0001281 | 0.062429784 | 0.958288 |
| STAD | HAVANA | gene | NA | NA | ENSG00000231246.2 | lncRNA | LINC02884 | 2 | HGNC:54808 | OTTHUMG00000 | 2.031528 | -0.54756 | 4.104811 | 0.000137 | 0.062429784 | 0.900069 |
| STAD | HAVANA | gene | NA | NA | ENSG00000187955.12 | protein_coding | COL14A1 | 1 | HGNC:2191 | OTTHUMG00000 | 1.891343 | 5.130438 | 4.10E+00 | 0.0001399 | 0.062429784 | 0.887441 |
| STAD | HAVANA | gene | NA | NA | ENSG00000241566.1 | IG_V_gene | IGKV2D-24 | 2 | HGNC:5797 | OTTHUMG00000 | 2.371749 | -2.33721 | 4.120349 | 0.0001301 | 0.062429784 | 0.867775 |
| STAD | HAVANA | gene | NA | NA | ENSG00000257277.2 | lncRNA | AC092652.1 | 2 | NA | OTTHUMG00000 | 2.159793 | -3.31494 | 4.142504 | 0.0001209 | 0.062429784 | 0.847252 |
| STAD | HAVANA | gene | NA | NA | ENSG00000181234.9 | protein_coding | TMEM132C | 2 | HGNC:25436 | OTTHUMG00000 | 2.632367 | -2.59571 | 4.100981 | 0.0001387 | 0.062429784 | 0.800216 |
| STAD | HAVANA | gene | NA | NA | ENSG00000254056.1 | IG_V_pseudogen | IGHV3-71 | 2 | HGNC:5621 | OTTHUMG00000 | 2.073098 | -4.00667 | 4.144831 | 0.00012 | 0.062429784 | 0.745787 |
| STAD | HAVANA | gene | NA | NA | ENSG00000187922.14 | protein_coding | LCN10 | 2 | HGNC:20892 | OTTHUMG00000 | 2.258847 | -4.59166 | 4.093135 | 0.0001423 | 0.062498568 | 0.520961 |
| STAD | HAVANA | gene | NA | NA | ENSG00000151892.15 | protein_coding | GFRA1 | 2 | HGNC:4243 | OTTHUMG00000 | 2.220109 | 2.370394 | 4.087244 | 0.0001451 | 0.062726507 | 0.908597 |
| STAD | HAVANA | gene | NA | NA | ENSG00000253763.1 | IG_V_pseudogen | IGHV3-6 | 2 | HGNC:5613 | OTTHUMG00000 | 2.450549 | -4.27638 | 4.077087 | 0.0001501 | 0.06385913 | 0.551813 |
| STAD | HAVANA | gene | NA | NA | ENSG00000248441.7 | lncRNA | LINC01197 | 2 | HGNC:49597 | OTTHUMG00000 | 1.661911 | -0.19158 | 4.056574 | 0.0001605 | 0.067272845 | 0.769592 |
| CESC | ENSEMBL | gene | NA | NA | ENSG00000199038.1 | miRNA | MIR210 | 3 | HGNC:31587 | NA | 1.712042 | -2.8511 | 4.672387 | 8.87E-06 | 0.0688362 | -0.28698 |
| STAD | HAVANA | gene | NA | NA | ENSG00000266524.3 | protein_coding | GDF10 | 2 | HGNC:4215 | OTTHUMG00000 | 2.501847 | -2.58155 | 4.037354 | 0.0001709 | 0.068946947 | 0.628943 |
| STAD | HAVANA | gene | NA | NA | ENSG00000153446.16 | protein_coding | C16orf89 | 1 | HGNC:28687 | OTTHUMG00000 | 2.405975 | -0.27518 | 4.009162 | 0.0001874 | 0.072508832 | 0.646071 |
| STAD | HAVANA | gene | NA | NA | ENSG00000116031.9 | protein_coding | CD207 | 2 | HGNC:17935 | OTTHUMG00000 | 2.249068 | -1.19934 | 4.00806 | 0.0001881 | 0.072508832 | 0.612686 |
| STAD | HAVANA | gene | NA | NA | ENSG00000055957.11 | protein_coding | ITIH1 | 2 | HGNC:6166 | OTTHUMG00000 | 2.019678 | -3.47177 | 3.998737 | 0.0001939 | 0.073479934 | 0.445366 |
| STAD | HAVANA | gene | NA | NA | ENSG00000013297.12 | protein_coding | CLDN11 | 1 | HGNC:8514 | OTTHUMG00000 | 1.831836 | 0.476305 | 3.970974 | 0.0002122 | 0.075700466 | 0.549541 |
| STAD | HAVANA | gene | NA | NA | ENSG00000250208.6 | lncRNA | FZD10-AS1 | 2 | HGNC:48632 | OTTHUMG00000 | 1.81081 | 0.027079 | 3.946702 | 0.0002296 | 0.075700466 | 0.471196 |
| STAD | HAVANA | gene | NA | NA | ENSG00000115353.11 | protein_coding | TACR1 | 2 | HGNC:11526 | OTTHUMG00000 | 1.947089 | -1.06314 | 3.943113 | 0.0002323 | 0.075700466 | 0.435224 |
| STAD | HAVANA | gene | NA | NA | ENSG00000111341.10 | protein_coding | MGP | 1 | HGNC:7060 | OTTHUMG00000 | 1.965435 | 5.911392 | 3.942713 | 0.0002326 | 0.075700466 | 0.397412 |
| STAD | HAVANA | gene | NA | NA | ENSG00000211890.4 | IG_C_gene | IGHA2 | 2 | HGNC:5479 | OTTHUMG00000 | 2.794082 | 6.272397 | 3.945766 | 0.0002303 | 0.075700466 | 0.393039 |
| STAD | HAVANA | gene | NA | NA | ENSG00000205018.2 | lncRNA | AC092384.1 | 2 | NA | OTTHUMG00000 | 1.55375 | -3.54544 | 3.949328 | 0.0002277 | 0.075700466 | 0.291177 |
| UCEC | HAVANA | gene | NA | NA | ENSG00000268649.5 | lncRNA | AL132655.2 | 2 | NA | OTTHUMG00000 | 3.106991 | -3.6813 | 4.789887 | 1.21E-05 | 0.075819649 | 1.502659 |
| STAD | HAVANA | gene | NA | NA | ENSG00000148488.17 | protein_coding | ST8SIA6 | 2 | HGNC:23317 | OTTHUMG00000 | 1.59583 | -0.64411 | 3.937997 | 0.0002362 | 0.075971221 | 0.428578 |
| STAD | HAVANA | gene | NA | NA | ENSG00000106809.11 | protein_coding | OGN | 2 | HGNC:8126 | OTTHUMG00000 | 2.834914 | 2.379277 | 3.920183 | 0.0002502 | 0.078393904 | 0.4174 |
| STAD | HAVANA | gene | NA | NA | ENSG00000169218.14 | protein_coding | RSPO1 | 2 | HGNC:21679 | OTTHUMG00000 | 2.281796 | -2.02842 | 3.917675 | 0.0002522 | 0.078393904 | 0.334798 |
| SARC | HAVANA | gene | NA | NA | ENSG00000254535.4 | protein_coding | PABPC4L | 2 | HGNC:31955 | OTTHUMG00000 | 1.924379 | 1.483409 | 4.682678 | 1.59E-05 | 0.079722283 | 2.688669 |
| SARC | HAVANA | gene | NA | NA | ENSG00000286451.1 | lncRNA | AL499602.1 | 2 | NA | OTTHUMG00000 | 3.349087 | -1.73564 | 4.695399 | 1.52E-05 | 0.079722283 | 2.318346 |
| SARC | HAVANA | gene | NA | NA | ENSG00000158164.7 | protein_coding | TMSB15A | 2 | HGNC:30744 | OTTHUMG00000 | 2.26054 | 1.729346 | 4.541615 | 2.63E-05 | 0.079722283 | 2.302076 |
| SARC | HAVANA | gene | NA | NA | ENSG00000285852.1 | lncRNA | AL353147.1 | 2 | NA | OTTHUMG00000 | 1.775466 | -2.04965 | 4.659706 | 1.73E-05 | 0.079722283 | 2.10543 |
| SARC | HAVANA | gene | NA | NA | ENSG00000139220.17 | protein_coding | PPFIA2 | 2 | HGNC:9246 | OTTHUMG00000 | 2.782829 | 0.021906 | 4.493264 | 3.13E-05 | 0.079722283 | 1.980901 |
| SARC | HAVANA | gene | NA | NA | ENSG00000165694.10 | protein_coding | FRMD7 | 2 | HGNC:8079 | OTTHUMG00000 | 3.155722 | -2.30106 | 4.588054 | 2.23E-05 | 0.079722283 | 1.924156 |
| STAD | HAVANA | gene | NA | NA | ENSG00000186479.5 | protein_coding | RGS7BP | 2 | HGNC:23271 | OTTHUMG00000 | 1.606608 | 0.429642 | 3.89258 | 0.0002735 | 0.080486528 | 0.327171 |
| STAD | HAVANA | gene | NA | NA | ENSG00000168309.18 | protein_coding | FAM107A | 1 | HGNC:30827 | OTTHUMG00000 | 1.548134 | 1.922547 | 3.885487 | 0.0002798 | 0.080486528 | 0.326054 |
| STAD | HAVANA | gene | NA | NA | ENSG00000281990.1 | IG_V_gene | IGHV1-69-2 | 2 | HGNC:5562 | OTTHUMG00000 | 3.559233 | -2.92366 | 3.893913 | 0.0002723 | 0.080486528 | 0.229073 |
| STAD | HAVANA | gene | NA | NA | ENSG00000269954.2 | lncRNA | AC022239.1 | 2 | NA | OTTHUMG00000 | 2.57018 | -2.79982 | 3.888042 | 0.0002775 | 0.080486528 | 0.219026 |
| STAD | HAVANA | gene | NA | NA | ENSG00000225398.3 | unprocessed_pse | PGM5P4 | 2 | HGNC:49605 | OTTHUMG00000 | 1.913055 | -3.27359 | 3.893131 | 0.000273 | 0.080486528 | 0.190833 |
| STAD | HAVANA | gene | NA | NA | ENSG00000254632.2 | lncRNA | AP003119.1 | 1 | NA | OTTHUMG00000 | 1.858194 | -3.87875 | 3.893655 | 0.0002725 | 0.080486528 | 0.115804 |
| STAD | HAVANA | gene | NA | NA | ENSG00000251003.9 | lncRNA | ZFPM2-AS1 | 2 | HGNC:50698 | OTTHUMG00000 | 2.049187 | -1.08614 | 3.811003 | 0.0003551 | 0.089305199 | 0.077744 |
| STAD | HAVANA | gene | NA | NA | ENSG00000172554.12 | protein_coding | SNTG2 | 2 | HGNC:13741 | OTTHUMG00000 | 2.038983 | -3.25394 | 3.813067 | 0.0003528 | 0.089305199 | -0.00991 |
| STAD | HAVANA | gene | NA | NA | ENSG00000268926.3 | lncRNA | AL354861.3 | 2 | NA | OTTHUMG00000 | 2.089296 | -3.44191 | 3.818453 | 0.0003468 | 0.089305199 | -0.01336 |
| STAD | HAVANA | gene | NA | NA | ENSG00000203506.5 | lncRNA | RBMS3-AS2 | 2 | HGNC:39988 | OTTHUMG00000 | 1.624969 | -3.4835 | 3.82E+00 | 0.0003485 | 0.089305199 | -0.03471 |
| STAD | HAVANA | gene | NA | NA | ENSG00000164591.14 | protein_coding | MYOZ3 | 2 | HGNC:18565 | OTTHUMG00000 | 1.697847 | -1.40973 | 3.81E+00 | 0.0003619 | 0.090091143 | 0.051462 |
| STAD | HAVANA | gene | NA | NA | ENSG00000181195.11 | protein_coding | PENK | 1 | HGNC:8831 | OTTHUMG00000 | 2.41435 | -2.92833 | 3.80E+00 | 0.0003702 | 0.090091143 | -0.02411 |
| STAD | HAVANA | gene | NA | NA | ENSG00000017427.17 | protein_coding | IGF1 | 1 | HGNC:5464 | OTTHUMG00000 | 2.186644 | 0.011452 | 3.79E+00 | 0.0003812 | 0.090408471 | 0.040993 |
| STAD | HAVANA | gene | NA | NA | ENSG00000249279.6 | lncRNA | LINC02057 | 1 | HGNC:52900 | OTTHUMG00000 | 2.003784 | -3.48342 | 3.77E+00 | 0.0004041 | 0.093147792 | -0.1407 |
| SARC | HAVANA | gene | NA | NA | ENSG00000167779.9 | protein_coding | IGFBP6 | 1 | HGNC:5475 | OTTHUMG00000 | -2.55583 | 6.118945 | -4.35E+00 | 5.25E-05 | 0.093762017 | 1.807329 |

| | | | | | | | | | | | | | | | | |
|---|---|---|---|---|---|---|---|---|---|---|---|---|---|---|---|---|
| SARC | HAVANA | gene | NA | NA | ENSG00000175928.6 | protein_coding | LRRN1 | 1 | HGNC:20980 | OTTHUMG00000 | 3.718743 | 1.372046 | 4.33E+00 | 5.62E-05 | 0.093762017 | 1.672278 |
| SARC | HAVANA | gene | NA | NA | ENSG00000279673.1 | TEC | AC092919.2 | 2 | NA | OTTHUMG00000 | 1.730822 | -0.33468 | 4.32E+00 | 5.80E-05 | 0.093762017 | 1.382864 |
| SARC | HAVANA | gene | NA | NA | ENSG00000257894.2 | lncRNA | AC027288.3 | 2 | NA | OTTHUMG00000 | 2.681677 | -2.2723 | 4.37E+00 | 4.80E-05 | 0.093762017 | 1.330531 |
| STAD | HAVANA | gene | NA | NA | ENSG00000179639.10 | protein_coding | FCER1A | 2 | HGNC:3609 | OTTHUMG00000 | 2.407001 | -1.03278 | 3.76E+00 | 0.0004143 | 0.094062662 | -0.04766 |
| STAD | HAVANA | gene | NA | NA | ENSG00000140538.16 | protein_coding | NTRK3 | 1 | HGNC:8033 | OTTHUMG00000 | 2.14625 | -0.6241 | 3.76E+00 | 0.0004185 | 0.094094501 | -0.05008 |
| STAD | HAVANA | gene | NA | NA | ENSG00000005102.14 | protein_coding | MEOX1 | 2 | HGNC:7013 | OTTHUMG00000 | 1.676338 | 0.883825 | 3.74E+00 | 0.0004472 | 0.095918771 | -0.08937 |
| SARC | HAVANA | gene | NA | NA | ENSG00000147180.17 | protein_coding | ZNF711 | 2 | HGNC:13128 | OTTHUMG00000 | 2.505332 | 1.834456 | 4.29E+00 | 6.44E-05 | 0.096768633 | 1.55329 |
| STAD | HAVANA | gene | NA | NA | ENSG00000152932.8 | protein_coding | RAB3C | 2 | HGNC:30269 | OTTHUMG00000 | 2.252676 | 0.159125 | 3.72E+00 | 0.0004795 | 0.099731342 | -0.1538 |
| STAD | HAVANA | gene | NA | NA | ENSG00000113594.10 | protein_coding | LIFR | 2 | HGNC:6597 | OTTHUMG00000 | 1.719336 | 3.705938 | 3.72089 | 0.0004725 | 0.099731342 | -0.17952 |
| **adj.p-val> 0.1** | | | | | | | | | | | | | | | | |
| **PRAD** | HAVANA | gene | NA | NA | ENSG00000106689.11 | protein_coding | LHX2 | 2 | HGNC:6594 | OTTHUMG00000 | 2.067652 | -3.36118 | 4.917567 | 5.55E-06 | 0.125102928 | -3.50387 |

| Gene names of differentially expressed genes (DEGs) between RR and RS across different cancer types | |
|---|---|
| **Gene symbol** | **David Gene name** |
| **protein_coding_genes** | |
| ABCA8 | ATP binding cassette subfamily A member 8(ABCA8) |
| ACKR1 | atypical chemokine receptor 1 (Duffy blood group)(ACKR1) |
| ACOX2 | acyl-CoA oxidase 2(ACOX2) |
| ADH1B | alcohol dehydrogenase 1B (class I), beta polypeptide(ADH1B) |
| ADRB2 | adrenoceptor beta 2(ADRB2) |
| AKR1C1 | aldo-keto reductase family 1 member C1(AKR1C1) |
| AKR1C2 | aldo-keto reductase family 1 member C2(AKR1C2) |
| APOA1 | apolipoprotein A1(APOA1) |
| ASTN1 | astrotactin 1(ASTN1) |
| BMP6 | bone morphogenetic protein 6(BMP6) |
| C16orf89 | chromosome 16 open reading frame 89(C16orf89) |
| C1QTNF7 | C1q and TNF related 7(C1QTNF7) |
| C1QTNF9 | C1q and TNF related 9(C1QTNF9) |
| C3orf36 | chromosome 3 putative open reading frame 36(C3orf36) |
| CADM3-AS1 | CADM3 antisense RNA 1(CADM3-AS1) |
| CCL19 | C-C motif chemokine ligand 19(CCL19) |
| CD207 | CD207 molecule(CD207) |
| CLDN11 | claudin 11(CLDN11) |
| COL14A1 | collagen type XIV alpha 1 chain(COL14A1) |
| CPE | carboxypeptidase E(CPE) |
| FAM107A | family with sequence similarity 107 member A(FAM107A) |
| FCER1A | Fc epsilon receptor Ia(FCER1A) |
| FRMD7 | FERM domain containing 7(FRMD7) |
| FZD10-AS1 | FZD10 antisense RNA 1(FZD10-AS1) |
| GDF10 | growth differentiation factor 10(GDF10) |
| GFRA1 | GDNF family receptor alpha 1(GFRA1) |

| | |
|---|---|
| GHR | growth hormone receptor(GHR) |
| GNAS-AS1 | GNAS antisense RNA 1(GNAS-AS1) |
| H2AC12 | H2A clustered histone 12(H2AC12) |
| HGF | hepatocyte growth factor(HGF) |
| HHIP | hedgehog interacting protein(HHIP) |
| IGF1 | insulin like growth factor 1(IGF1) |
| IGFBP6 | insulin like growth factor binding protein 6(IGFBP6) |
| ITIH1 | inter-alpha-trypsin inhibitor heavy chain 1(ITIH1) |
| JAM2 | junctional adhesion molecule 2(JAM2) |
| KCNS2 | potassium voltage-gated channel modifier subfamily S member 2(KCNS2) |
| KCNT2 | potassium sodium-activated channel subfamily T member 2(KCNT2) |
| KIF25-AS1 | KIF25 antisense RNA 1(KIF25-AS1) |
| LCN10 | lipocalin 10(LCN10) |
| LGI1 | leucine rich glioma inactivated 1(LGI1) |
| LHX2 | LIM homeobox 2(LHX2) |
| LIFR | LIF receptor subunit alpha(LIFR) |
| LRRN1 | leucine rich repeat neuronal 1(LRRN1) |
| MEOX1 | mesenchyme homeobox 1(MEOX1) |
| MGP | matrix Gla protein(MGP) |
| MS4A15 | Membrane-Spanning 4-Domains Subfamily A Member 15(MS4A15) |
| MYOZ3 | myozenin 3(MYOZ3) |
| MYPN | myopalladin(MYPN) |
| NTRK3 | neurotrophic receptor tyrosine kinase 3(NTRK3) |
| OGN | osteoglycin(OGN) |
| PABPC4L | poly(A) binding protein cytoplasmic 4 like(PABPC4L) |
| PALMD | palmdelphin(PALMD) |
| PENK | proenkephalin(PENK) |
| PGM5P3-AS1 | PGM5P3 antisense RNA 1(PGM5P3-AS1) |
| PGM5P4 | phosphoglucomutase 5 pseudogene 4(PGM5P4) |
| PPFIA2 | PTPRF interacting protein alpha 2(PPFIA2) |

| | |
|---|---|
| RAB3C | RAB3C, member RAS oncogene family(RAB3C) |
| RBMS3-AS2 | RBMS3 antisense RNA 2(RBMS3-AS2) |
| RCAN2 | regulator of calcineurin 2(RCAN2) |
| RERG | RAS like estrogen regulated growth inhibitor(RERG) |
| RGS7BP | regulator of G protein signaling 7 binding protein(RGS7BP) |
| RNASE7 | ribonuclease A family member 7(RNASE7) |
| RSPO1 | R-spondin 1(RSPO1) |
| SCARA5 | scavenger receptor class A member 5(SCARA5) |
| SELP | selectin P(SELP) |
| SLCO2A1 | solute carrier organic anion transporter family member 2A1(SLCO2A1) |
| SNTG2 | syntrophin gamma 2(SNTG2) |
| ST8SIA6 | ST8 alpha-N-acetyl-neuraminide alpha-2,8-sialyltransferase 6(ST8SIA6) |
| TACR1 | tachykinin receptor 1(TACR1) |
| TCF23 | transcription factor 23(TCF23) |
| TMEM132C | transmembrane protein 132C(TMEM132C) |
| TMSB15A | thymosin beta 15A(TMSB15A) |
| TNFRSF11B | TNF receptor superfamily member 11b(TNFRSF11B) |
| WIF1 | Wnt Inhibitory Factor 1(WIF1) |
| ZBTB16 | zinc finger and BTB domain containing 16(ZBTB16) |
| ZFPM2-AS1 | ZFPM2 antisense RNA 1(ZFPM2-AS1) |
| ZNF711 | zinc finger protein 711(ZNF711) |
| **non_coding_RNAs** | |
| AC012409.5 | - |
| AC022239.1 | - |
| AC024337.2 | - |
| AC027288.3 | - |
| AC069360.1 | - |
| AC092384.1 | - |
| AC092652.1 | - |

| | |
|---|---|
| AC092919.2 | - |
| AC104260.2 | - |
| AL035670.1 | - |
| AL132655.2 | - |
| AL353147.1 | - |
| AL354861.3 | - |
| AL499602.1 | - |
| AP000224.1 | - |
| AP001528.2 | - |
| AP003119.1 | - |
| MIR210 | microRNA 210(MIR210) |
| LINC01197 | - |
| LINC02057 | long intergenic non-protein coding RNA 2057(LINC02057) |
| LINC02884 | long intergenic non-protein coding RNA 2884(LINC02884) |

| **IG_V_gene** | |
|---|---|
| IGHA2 | immunoglobulin heavy constant alpha 2 (A2m marker)(IGHA2) |
| IGHV1-69-2 | immunoglobulin heavy variable 1-69-2(IGHV1-69-2) |
| IGHV2-5 | immunoglobulin heavy variable 2-5(IGHV2-5) |
| IGHV3-6 | immunoglobulin heavy variable 3-6 (pseudogene)(IGHV3-6) |
| IGHV3-71 | immunoglobulin heavy variable 3-71 (pseudogene)(IGHV3-71) |
| IGHV3OR16-13 | immunoglobulin heavy variable 3/OR16-13 (non-functional)(IGHV3OR16-13) |
| IGKV2-28 | immunoglobulin kappa variable 2-28(IGKV2-28) |
| IGKV2D-24 | immunoglobulin kappa variable 2D-24 (non-functional)(IGKV2D-24) |
| IGKV2D-28 | immunoglobulin kappa variable 2D-28(IGKV2D-28) |
| IGLV7-46 | immunoglobulin lambda variable 7-46(IGLV7-46) |
