## Supplementary table_6 for "Direct Pan-Cancer Multi-Omic Analysis of Radiotherapy Response Reveals Coding and Regulatory Genomic Variation, and Extracellular Adaptive Programs Underlying Radioresistance"

RSI trends across cancer types

| cancer | n_RS | n_RR | median_RS | median_RR | wilcoxon_p | direction | delta_RSI | FDR |
|---|---|---|---|---|---|---|---|---|
| THCA | 178 | 12 | 0.739567 | 0.789063 | 0.10313 | RR > RS | 0.049496 | 0.766017 |
| BRCA | 176 | 12 | 0.534052 | 0.655012 | 0.153203 | RR > RS | 0.12096 | 0.766017 |
| HNSC | 146 | 16 | 0.347393 | 0.400906 | 0.912828 | RR > RS | 0.053513 | 0.97803 |
| LGG | 42 | 120 | 0.964453 | 0.977217 | 0.833505 | RR > RS | 0.012764 | 0.97803 |
| CESC | 85 | 17 | 0.289985 | 0.285672 | 0.60878 | RS > RR | -0.00431 | 0.913169 |
| PRAD | 52 | 16 | 0.52856 | 0.540164 | 0.712369 | RR > RS | 0.011604 | 0.971413 |
| STAD | 38 | 14 | 0.564381 | 0.651373 | 0.265263 | RR > RS | 0.086991 | 0.793717 |
| UCEC | 48 | 7 | 0.410286 | 0.393584 | 0.604665 | RS > RR | -0.0167 | 0.913169 |
| ESCA | 35 | 14 | 0.673183 | 0.464664 | 0.005862 | RS > RR | -0.20852 | 0.087925 |
| LUAD | 17 | 29 | 0.401977 | 0.436012 | 0.399779 | RR > RS | 0.034034 | 0.793717 |
| SARC | 42 | 17 | 0.903713 | 0.940424 | 0.388725 | RR > RS | 0.03671 | 0.793717 |
| PAAD | 14 | 13 | 0.477905 | 0.650556 | 0.423316 | RR > RS | 0.172651 | 0.793717 |
| BLCA | 16 | 12 | 0.290371 | 0.342937 | 0.390428 | RR > RS | 0.052566 | 0.793717 |
| LUSC | 14 | 13 | 0.362643 | 0.359258 | 0.903441 | RS > RR | -0.00338 | 0.97803 |
| SKCM | 23 | 22 | 0.544489 | 0.634975 | 0.981884 | RR > RS | 0.090486 | 0.981884 |

**Overall survival statistics of RR and RS patients**

| treatment_best_response | N | Deaths | Censored | Median_OS_years | Median_lower_95CI | Median_upper_95CI | OS_5year | OS_5year_lower95 | OS_5year_upper95 | OS_5year_percent | OS_5year_lower95_percent | OS_5year_upper95_percent | Overall_logrank_p |
|---|---|---|---|---|---|---|---|---|---|---|---|---|---|
| **Complete response** | 956 | 112 | 844 | NA | 12.81314168 | NA | 0.828162 | 0.792672 | 0.865241 | 82.81621 | 79.26721 | 86.5241 | 3.64E-86 |
| **Partial response** | 93 | 23 | 70 | 7.216974675 | 4.947296372 | NA | 0.617082 | 0.477202 | 0.797966 | 61.70824 | 47.72016 | 79.79662 | 3.64E-86 |
| **Stable disease** | 167 | 48 | 119 | 5.292265572 | 4.320328542 | NA | 0.560414 | 0.445674 | 0.704694 | 56.04141 | 44.56744 | 70.46938 | 3.64E-86 |
| **Radiographic progressive disease** | 224 | 153 | 71 | 1.77412731 | 1.538672142 | 2.242299795 | 0.224849 | 0.166245 | 0.304111 | 22.48491 | 16.62454 | 30.41115 | 3.64E-86 |
| **response_category** | **N** | **Deaths** | **Censored** | **Median_OS_years** | **Median_lower_95CI** | **Median_upper_95CI** | **OS_5year** | **OS_5year_lower95** | **OS_5year_upper95** | **OS_5year_percent** | **OS_5year_lower95_percent** | **OS_5year_upper95_percent** | |
| **Radiosensitive** | 1049 | 135 | 914 | NA | 12.81314168 | NA | 0.808141 | 0.77229047 | 0.845655355 | 80.81408 | 77.22905 | 84.56554 | |
| **Radioresistant** | 391 | 201 | 190 | 2.855578371 | 2.425735797 | 3.679671458 | 0.350609 | 0.28954568 | 0.424549613 | 35.06088 | 28.95457 | 42.45496 | |

immune_effect_matrix_purity-adjusted

| cell_type | BLCA | CESC | HNSC | LUAD | STAD | UCEC | BRCA | ESCA | THCA | SARC | LGG | PRAD | PAAD | LUSC |
|---|---|---|---|---|---|---|---|---|---|---|---|---|---|---|
| T.cells.CD8 | -0.01499 | -0.02915 | 0.044138 | -0.01047 | -0.00826 | -0.02012 | -0.02371 | 0.007702 | -0.03604 | 0.010957 | -0.00321 | 0.001645 | 0.014322 | -0.02128 |
| NK.cells.activated | 0.025618 | 0.013129 | 0.016872 | -0.00068 | -0.01367 | 0.015579 | -0.00458 | -0.01588 | 0.00771 | 0.002037 | 0.005809 | -0.01514 | -0.00758 | -0.01178 |
| NK.cells.resting | -0.00804 | 0.000921 | -0.01113 | 0.000973 | 0.00257 | -0.00849 | 0.003818 | 0.007932 | -0.01007 | -0.00054 | 0.005992 | 0.000772 | 0.011654 | -0.00543 |
| T.cells.CD4.memory.activated | -0.00756 | -0.0083 | -0.00042 | 0.001951 | -0.00572 | -0.00085 | -0.00133 | 0.006754 | -0.00033 | -0.00256 | 0.000112 | -0.00018 | -0.00013 | -0.00211 |
| T.cells.CD4.memory.resting | 0.007471 | 0.00455 | -0.01709 | 0.031121 | -0.00381 | -0.04111 | -0.00653 | -0.00133 | 0.024537 | 0.003101 | 0.015236 | -0.00127 | 0.018228 | -0.02083 |
| T.cells.CD4.naive | 0.007587 | -0.01077 | -0.00298 | 0 | -7.62E-05 | -0.00163 | -0.00055 | -0.00457 | -0.00289 | -0.00341 | -0.00458 | -0.00069 | -0.00031 | 0.003448 |
| T.cells.follicular.helper | 0.00168 | -0.01429 | 0.007575 | -0.01272 | -0.00909 | -0.00125 | -0.01196 | 0.025096 | -0.00055 | 0.014054 | -0.00471 | -0.01775 | -0.02154 | -0.00381 |
| T.cells.regulatory..Tregs. | -0.01764 | -0.00525 | -0.01253 | -0.00532 | 0.005046 | 0.001735 | -0.00908 | -0.00823 | 0.015961 | -0.00015 | -0.0008 | 0.008435 | 0.001432 | 0.00342 |
| T.cells.gamma.delta | 0.006334 | -0.00087 | -0.00016 | -0.00157 | 0 | -0.00133 | -0.00034 | -0.00078 | -0.00114 | 0.001028 | 0 | -0.0002 | 0 | 0.002442 |
| B.cells.memory | -0.03878 | -0.0039 | -0.00766 | -0.00574 | 0.012752 | -0.00746 | -0.00874 | 0.012656 | -0.02902 | -0.00169 | -0.007 | 0.004297 | -0.03085 | -0.01162 |
| B.cells.naive | -0.01731 | -0.00222 | -0.00958 | 0.009456 | 0.019106 | -0.0028 | 0.000465 | 0.033536 | 0.058301 | 0.006172 | -0.00598 | 0.013117 | 0.000434 | 0.026352 |
| Plasma.cells | -0.01002 | 0.024252 | 0.003717 | 0.026094 | 0.010535 | -0.00569 | -0.00152 | 0.005516 | 0.042508 | 0.002455 | -0.00296 | -0.00989 | 0.000709 | -0.00421 |
| Monocytes | -0.0035 | -0.00456 | -0.00124 | -0.00349 | 0.001045 | -0.00531 | 0.007929 | 0.00408 | -0.00589 | -0.00604 | 0.014321 | -9.46E-05 | 0.059647 | -0.00238 |
| Macrophages.M0 | 0.00872 | 0.025146 | -0.0155 | 0.010607 | -0.00912 | 0.048601 | 0.01456 | -0.05654 | 0.035519 | 0.00176 | -0.01085 | -0.00359 | -0.07756 | 0.071877 |
| Macrophages.M1 | -0.00032 | 0.007876 | 0.014504 | 0.007861 | 0.007619 | 0.013851 | -0.00402 | 0.058843 | -0.00068 | -0.00589 | -0.00327 | -0.00742 | -0.01342 | -0.01002 |
| Macrophages.M2 | 0.057019 | -0.00774 | -0.00416 | 0.012733 | 0.00309 | 0.037751 | 0.044819 | -0.08894 | -0.05961 | 0.006948 | -0.007 | 0.043177 | 0.086604 | 0.029464 |
| Dendritic.cells.activated | -0.01708 | -0.02615 | 0.000904 | -0.0196 | 0.001579 | -0.01259 | -0.00392 | 0.019415 | -0.00746 | 0.002107 | 0.001524 | -0.00205 | -0.01015 | -0.01827 |
| Dendritic.cells.resting | 0.011213 | -0.00389 | -0.00718 | -0.01516 | 0.008727 | 0.000834 | 0.000811 | -0.01052 | 0.008727 | -0.00019 | -0.00015 | -0.00422 | -0.02246 | -0.01391 |
| Neutrophils | -0.00045 | 0.010909 | 0.002908 | -0.00172 | -0.00428 | -0.00178 | -0.00345 | 0.008716 | -0.00013 | -0.00118 | 0.005536 | -0.00123 | 0.001612 | -0.01163 |
| Eosinophils | -0.0011 | -0.00168 | 0.000114 | 0 | -0.0023 | 0.001677 | -0.00016 | -0.0009 | -0.00216 | 2.10E-08 | 0.000538 | -0.00065 | 0 | 0 |
| Mast.cells.activated | 0.021246 | 0.032676 | 0.001174 | -0.00409 | -0.03229 | -0.0116 | -0.00223 | 0.014865 | -0.00317 | 0.00058 | 0.012528 | 0.007528 | 0.001755 | 0.003292 |
| Mast.cells.resting | -0.01011 | -0.0007 | -0.00227 | -0.02025 | 0.016561 | 0.001959 | 0.009716 | -0.01741 | -0.03414 | -0.02955 | -0.0111 | -0.01461 | -0.0124 | -0.00303 |
